# Device-measured vigorous intermittent lifestyle physical activity and major cardiovascular events

**DOI:** 10.1101/2023.10.23.23297430

**Authors:** Emmanuel Stamatakis, Matthew N. Ahmadi, Raaj Kishore Biswas, Borja del Pozo Cruz, Cecilie Thøgersen-Ntoumani, Marie H Murphy, Angelo Sabag, Scott A. Lear, Jason MR Gill, Clara K Chow, Mark Hamer

**Affiliations:** Mackenzie Wearables Research Hub, Charles Perkins Centre, The University of Sydney, Sydney, Australia; School of Health Sciences, Faculty of Medicine and Health, The University of Sydney, NSW, Australia; Faculty of Education, University of Cádiz, Cádiz, Spain; Biomedical Research and Innovation Institute of Cádiz (INiBICA) Research Unit, Puerta del Mar University Hospital, University of Cádiz, Spain; Department of Sports Science and Clinical Biomechanics, University of Southern Denmark, Odense, Denmark; Danish Centre for Motivation and Behaviour Science, Department of Sports Science and Clinical Biomechanics, University of Southern Denmark, Denmark; Physical Activity for Health Research Centre, Moray House School of Education and Sport, University of Edinburgh; Faculty of Health Sciences, Simon Fraser University, Vancouver, Canada; School of Cardiovascular and Metabolic Health, University of Glasgow, Glasgow, United Kingdom; Westmead Applied Research Centre, University of Sydney and Department of Cardiology, Westmead Hospital, Sydney, Australia; Institute Sport Exercise Health, Division Surgery Interventional Science, University College London, U.K

**Keywords:** wearables, physical activity, exercise, mortality, cohort studies, epidemiology, machine learning

## Abstract

**Importance:** Vigorous physical activity is a time-efficient and potent preventive intervention for major adverse cardiovascular events (MACE), although longer traditional exercise sessions are unappealing or inaccessible to most adults.

**Objective:** We examined the dose-response associations of device-measured vigorous intermittent lifestyle physical activity (VILPA, brief sporadic bouts of higher intensity occurring during daily living) with MACE and its sub-types in women and men. We also undertook analogous analyses in a sample of exercisers.

**Design, Setting, and Participants:** Prospective cohort analysis of 13,018 women and 9,350 men non-exercisers from the UK Biobank accelerometry sub-study; the contextual analyses involved 34,364 female/24,284 male exercisers from the same sub-study.

**Exposures:** Wrist accelerometer assessed daily VILPA duration of bouts lasting up to 1 and up to 2 minutes.

**Outcomes and Measures:** Overall and sex-specific dose-response associations of daily VILPA with MACE and its subtypes (incident myocardial infarction, heart failure and stroke).

**Results:** Among female/male non-exercisers there were 331/488 all-MACE events (129/250 myocardial infarction, 96/119 heart failure,106/119 stroke events) over a mean 7.9-year follow-up. Daily VILPA duration exhibited a near-linear dose-response association with all MACE, myocardial infarction, and heart failure in women but not in men. Compared to women with no VILPA, the median daily VILPA duration of 3.4 minutes per day was associated with HRs of 0.55 (0.41, 0.75) for all MACE; and 0.33 (0.18, 0.59) for heart failure. Women’s minimum doses (the dose associated with 50% of the optimal risk reduction) of 1.2-1.6 minutes of VILPA per day were associated with HRs of 0.70 (0.58, 0.86) for all-MACE, 0.67 (0.50, 0.91) for myocardial infarction and 0.60 (0.45, 0.81) for heart failure, respectively. The equivalent analyses in exercisers in the UK Biobank showed comparable beneficial associations of vigorous intensity activity with all MACE, myocardial infarction and heart failure in both sex groups.

**Conclusions and Relevance:** Amongst non-exercisers, small amounts of VILPA were associated with substantially lower risk of myocardial infarction and heart failure in women but not in men. No such sex differences were evident among exercisers. VILPA may be a promising physical activity target for CVD prevention in women not willing or able to exercise.

## Key Points

**Question:** What are the dose-response associations between vigorous intermittent lifestyle physical activity (VILPA) and major adverse cardiovascular events (cardiovascular disease related death or incidence of myocardial infarction, heart failure, or stroke) in women and men?

**Findings:** VILPA exhibited a near-linear dose-response association with most outcomes in women but not in men. The median daily VILPA duration of 3.4 minutes per day was associated with 45% (25%-59%) lower risk of a major adverse cardiovascular event in women.

**Meaning:** Amongst non-exercisers, small amounts of VILPA were associated with substantially lower risk of myocardial infarction and heart failure in women but not in men. VILPA may be a promising physical activity target for CVD prevention in women not willing or able to exercise.

## INTRODUCTION

Cardiovascular disease (CVD) is the leading cause of death in both men and women globally^1^. Major adverse cardiovascular events (MACE), defined as non-fatal stroke/myocardial infarction/heart failure or cardiovascular death^2^ are a commonly used composite of main CVD outcomes in trials and observational studies. Clinical and public health practice have traditionally focused on the cardio-protective properties of longer bouts of physical activity done during structured exercise sessions. The dose-response associations of shorter bouts of vigorous intensity physical activity and cardiovascular outcomes is less clear. Compared with lower intensities, vigorous physical activity (≥6 absolute metabolic equivalents of task) elicits more pronounced cardiovascular effects^3–6^ in a shorter period of time^7^. Despite these advantages, vigorous intensity exercise is not feasible or appealing to most middle-aged adults^8^.

Vigorous intermittent lifestyle physical activity (VILPA)^4,9^ refers to brief and sporadic (e.g. up to one minute long)^4,10^ bouts that are done during daily living. Since short bouts of physical activity cannot be captured by questionnaires, wearable trackers are essential for measuring VILPA^4,10^. A recent study in non-exercisers^4^ (i.e., individuals reporting no leisure-time exercise) found a beneficial association of daily VILPA with cardiovascular mortality, although the relatively low number of events precluded a detailed examination of dose-response, or examining sex-specific associations with disease sub-types.

Sex differences in pathophysiology may moderate the influence of risk factors (including physical activity) on heart failure, myocardial infarction, and stroke^11^. Women have lower cardiorespiratory fitness than men^12^, making the level of physical effort for a given physical task (and hence the physiological stimulus for adaptation) higher for women. Despite the established sex differences in fitness, and in vascular, muscular, and respiratory responses to physical activity of higher intensity^13^, there is no evidence on whether sex differences exist in the long-term cardiovascular health effects of vigorous physical activity. Such evidence is critical to inform appropriate sex-specific clinical and public health guidelines for CVD prevention^14^.

We examined the overall and sex-specific dose-response associations of daily VILPA duration and frequency with MACE and its sub-types, and estimated minimal VILPA amounts for MACE risk reduction. To understand the role of physical activity context, we also examined the analogous dose-response associations of (exercise or non-exercise) vigorous physical activity with the same MACE outcomes among exercisers in the same cohort.

## METHODS

### Sample & design

The UK Biobank Study is a prospective cohort study of adults aged between 40-69 years at baseline (2006-10). Participants provided informed consent and ethical approval was provided by the UK’s National Health Service, National Research Ethics Service (Ref80 11/NW/0382).

Between 2013 and 2015, 103,684 UK Biobank participants wore a wrist-worn accelerometer for 7 days^15,16^. We defined a valid monitoring day as wear time greater than 16 hours. To be included in the analysis, participants were required to have at least three valid monitoring days, including at least one weekend day^4,5,10,17^. We excluded participants with insufficient valid wear days, those who had missing covariate data and participants who reported an inability to walk. **eFigure 1** shows the derivation of the core analytic samples of non-exercisers.

As previously described^4,10^, we examined VILPA by separating accelerometry sub-study participants who self-reported no leisure time exercise participation and no more than one recreational walk per week^4,10^. To provide a comparison between the sex-specific effects of VILPA and (context-agnostic) vigorous physical activity, we repeated the main analyses among the UK Biobank accelerometry sub-study participants who self-reported participation in leisure-time exercise^4^ (**eFigure 2)**.

### Physical activity assessment and exposure variables

We have described the physical activity intensity classification method in detail elsewhere^4,5,10^ and in **eText 1**. In brief, physical activity intensity was classified into light, moderate, and vigorous using a validated^4,5^ 2-stage machine learning-based Random Forest activity classifier. In the 88 non-exercisers from the Australian validation sample, the correct classification of predicted VILPA against ground truth was >94.0% for females and males (**eText 1)**. We focused on short bouts lasting <2 minutes, based on recent data^4^ showing the mean time required to reach vigorous intensity during typical VILPA activities is 73.5 (*SD*=26.2) seconds. Considering 89.1% of all VILPA bouts lasted up to 1 minute and only 3.8% of bouts lasted 1-2 minutes, we present detailed data only for the former with indicative results presented for the latter length. As previously described^4^, for daily VILPA frequency analyses we length-standardized bouts to one minute to allow a more concrete interpretation of the corresponding effect sizes (**eText 2**). We also present VILPA frequency analyses with raw (unstandardised) bouts.

We have previously^4,10^ described in detail the non-exercisers sample selection. We used information on leisure-time exercise and recreational walking participation available in the baseline of the UK Biobank study. Among the 6,095 UK Biobank accelerometry sample participants who reported no exercise at baseline and had a re-examination on average 1.5 (*SD*=1.4) years prior to the accelerometry measurements, 88% maintained their non-exercise status over time.

### Mortality and MACE ascertainment

Participants were followed up through November 30^th^, 2022, with deaths obtained via linkage with the National Health Service (NHS) Digital of England and Wales or the NHS Central Register and National Records of Scotland. Inpatient hospitalisation data were provided by either the Hospital Episode Statistics for England, the Patient Episode Database for Wales, or the Scottish Morbidity Record for Scotland. MACE was defined as CVD death or incidence of ST-elevated or non-ST elevated myocardial infarction (International Classification of Diseases Version 10: I21, I23, I24, I25, I26, I30, I31, I33, I34, I35, I38, I42, I45, I46, I48), stroke (I60, I61, I63, I64, I67), and heart failure (I11, II13, I50, I51).

### Statistical analyses

To reduce the risk of reverse causation through prodromal/undiagnosed disease, we excluded those with an event within the first 2 years of follow up and those with prevalent CVD at the accelerometry baseline. The upper range of VILPA/VPA values were truncated at the 97.5 percentile to minimize the effect of sparse data or outliers^4,5,10^.

Using Fine-Gray sub-distribution hazards to account for competing risks from non-CVD deaths^18^, we examined dose-response of average daily duration, and length standardised and raw frequency of VILPA bouts, as well as the corresponding (context-agnostic) VPA variables in exercisers. Since the distribution of primary exposures (VILPA and VPA) were highly skewed, knots were placed on the higher density data areas^19^ at equally distributed frequencies (1st, 33^rd^, 67th percentiles). Departure from linearity was assessed by a Wald test. Proportional hazards assumptions were tested using Schoenfeld Residuals in the models with all three outcomes and no violations were observed (all p>0.05). Core analyses were adjusted for age, sex (except for sex-specific models), average daily duration of light and moderate physical activity, average duration of VILPA/VPA coming from bouts lasting over 1 or 2 minutes, smoking history, alcohol consumption, accelerometer estimated sleep duration, diet, education, ethnicity, self-reported parental history of CVD, prevalent cancer, and self-reported medication use (cholesterol, blood pressure, and diabetes) (**eTable 9)**. To prevent multicollinearity, raw frequency of VILPA was adjusted for the residual of VILPA duration. For sex-specific models, interaction effects were assessed and if it was found to be associated with the outcome (p-value<0.05), an interaction model was plotted. Otherwise, independent models were plotted stratifying the sample by sex. The reference data point for all main models were zero minutes of VILPA/VPA per day^4^.

To provide conservative point estimates we calculated the “minimal dose”, defined as VILPA/VPA, volume/frequency associated with 50% of the optimal risk reduction^4,5,20,21^. We also present point estimates (hazard ratios and 95% confidence intervals) associated with the median volume/frequency VILPA/VPA values. We calculated E-values to estimate the plausibility of bias from unmeasured confounding. To provide a broader physical activity context to sex differences, we also examined the dose response curves of light and moderate intensity physical activity against MACE outcomes.

We conducted sensitivity analyses of VILPA with additional adjustment for potential mediators, i.e., glycated haemoglobin (HbA1c), low-density lipoprotein (LDL), high-density lipoprotein (HDL), triglycerides, systolic blood pressure, diastolic blood pressure, and body mass index (BMI). To further reduced reverse causation bias we also excluded participants who had poor self-rated health or a BMI below 18.5 kg/m^2^ or current smokers^4^ or a frailty index ≥3 (on a 0 to 5 scale)^22^. To assess the influence of variations of the reference data point on estimates, we repeated sex-specific analyses of VILPA duration using the 15^th^ percentile of the VILPA duration distribution as referent (0.63 minutes of VILPA per day). To assist the interpretation of our findings we calculated physical activity energy expenditure during VILPA bouts and relative physical activity intensity (%VO_2_max) during VILPA bouts in the subsample of 2,043 female and 1,588 male non-exercisers with valid accelerometry and fitness test data (**eText 3**).

We performed all analysis using R statistical software (version 4.2.3) with the RMS (version 6.3.0) and survival packages (version 3.5.5). We reported this study according to Strengthening the Reporting of Observational Studies in Epidemiology (STROBE) guidelines (**eTable 12)**.

## RESULTS

### Sample

**eFigure 1** shows the sample derivation process which resulted in 22,368 (13,018 women/9,350 men) UK Biobank participants being included in the analyses, corresponding to 819 MACE events (331 female/488 male). Slightly smaller sample sizes were entered into the analysis of myocardial infarction (n=21,928; 379 events (129 female / 250 male)), heart failure (n = 21,764; 215 events (96 female /119 male)), and stroke (n = 21,774; 225 events (106 female /119 male)). We also considered analyses for stroke sub-types, but these were less feasible due to the very low number of events for hemorrhagic stroke (n = 21,596; 47 events (30 female/17 male)) vs ischemic (n = 21,774; 175 events (76 female/99 male)).

**Table 1** presents the characteristics of the sample by sex and daily VILPA duration. The mean age of participants was 61.9 (7.6) years and mean follow-up was 7.9 (1.0) years corresponding to 176,678 person-years. To understand the role of how confounding by indication may have influenced sex-specific, **eText 4** provides a comparison between the referent groups of females and males. **eFigure 2, eTable 1**, and **eText 4** describe the characteristics of the exercisers sample. **eTable 2** and **eText 4** presents descriptives of bout length of VILPA (non-exercisers) and VPA (exercisers).

**Table 1:**
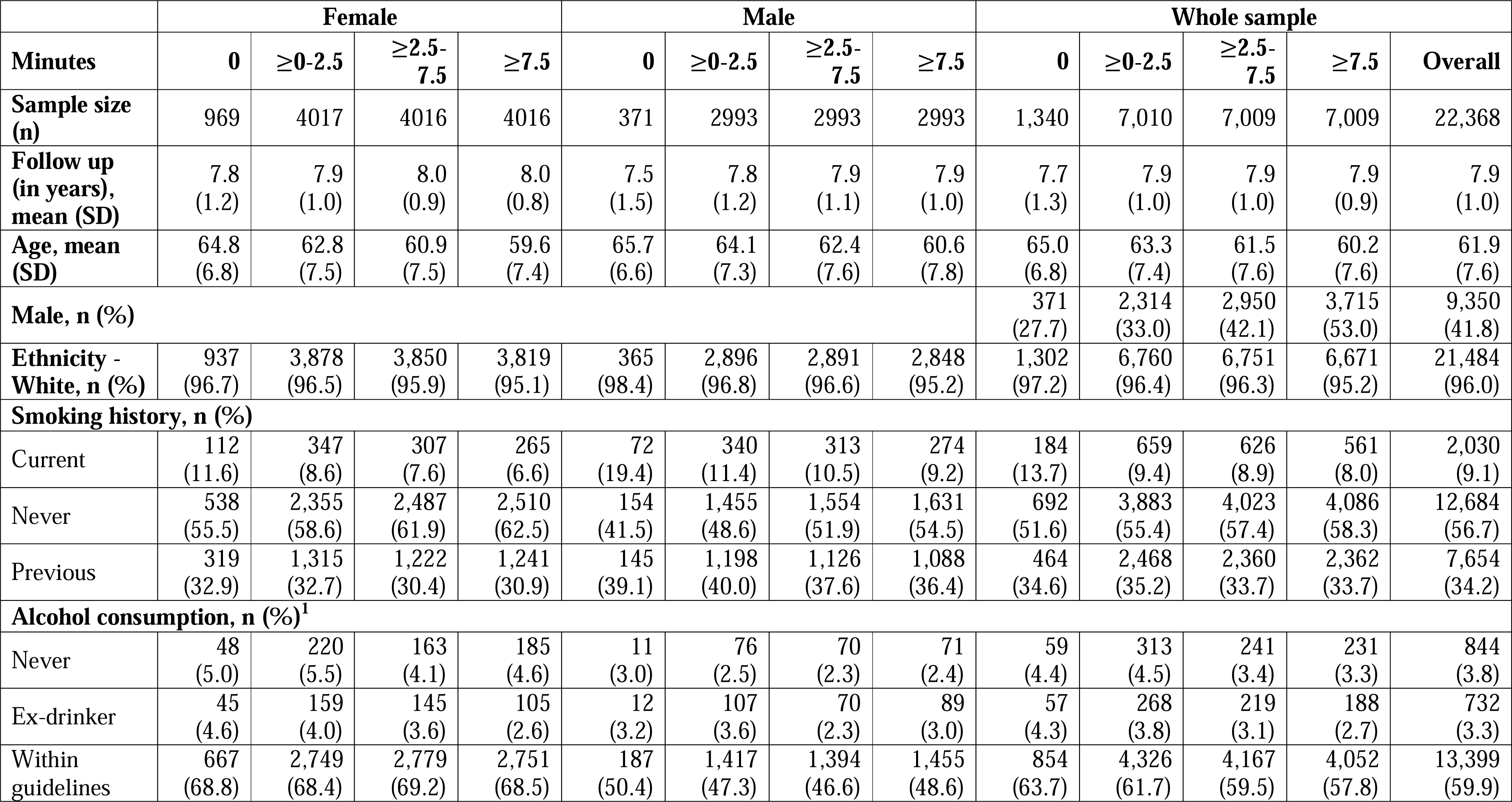
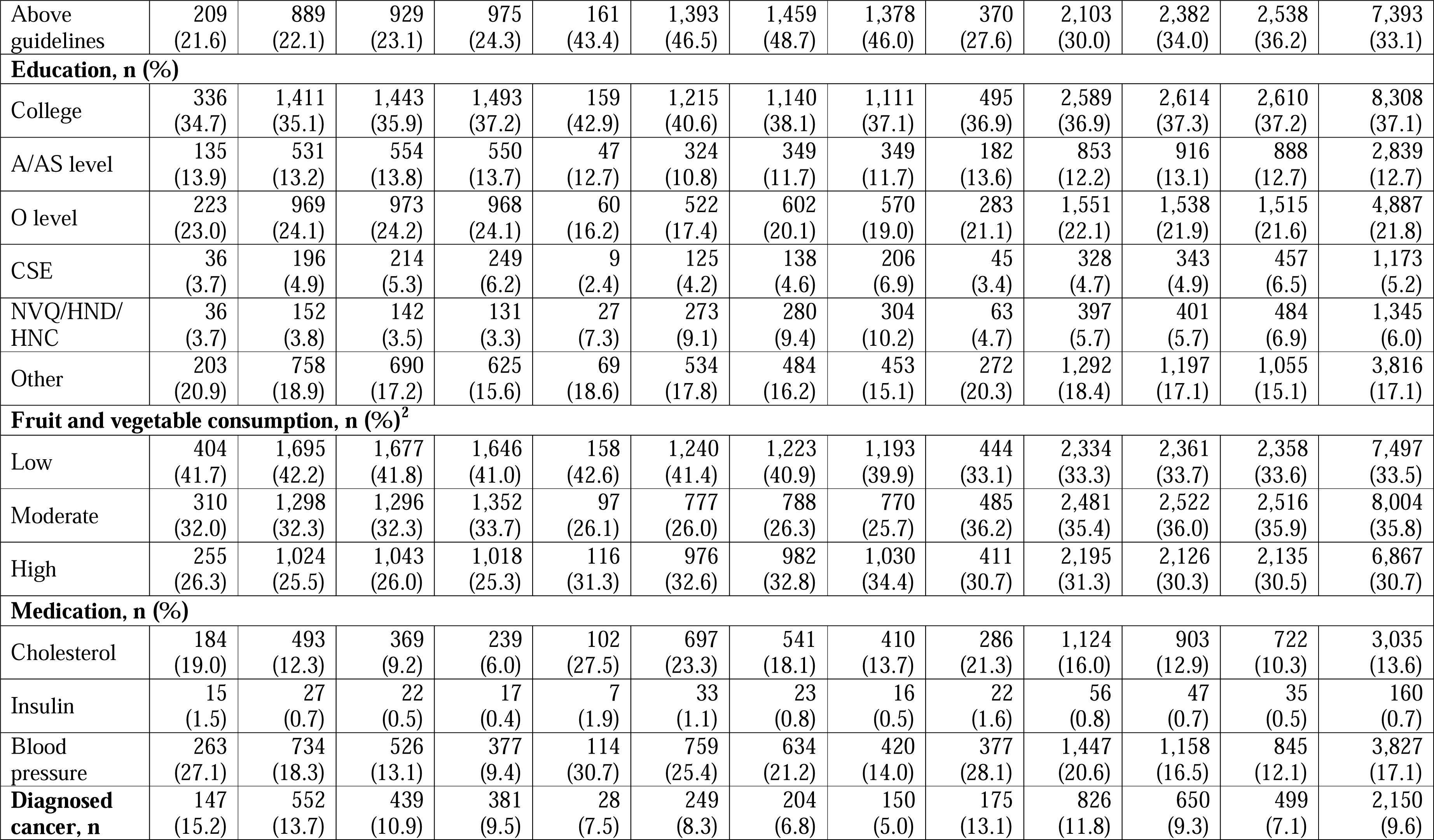
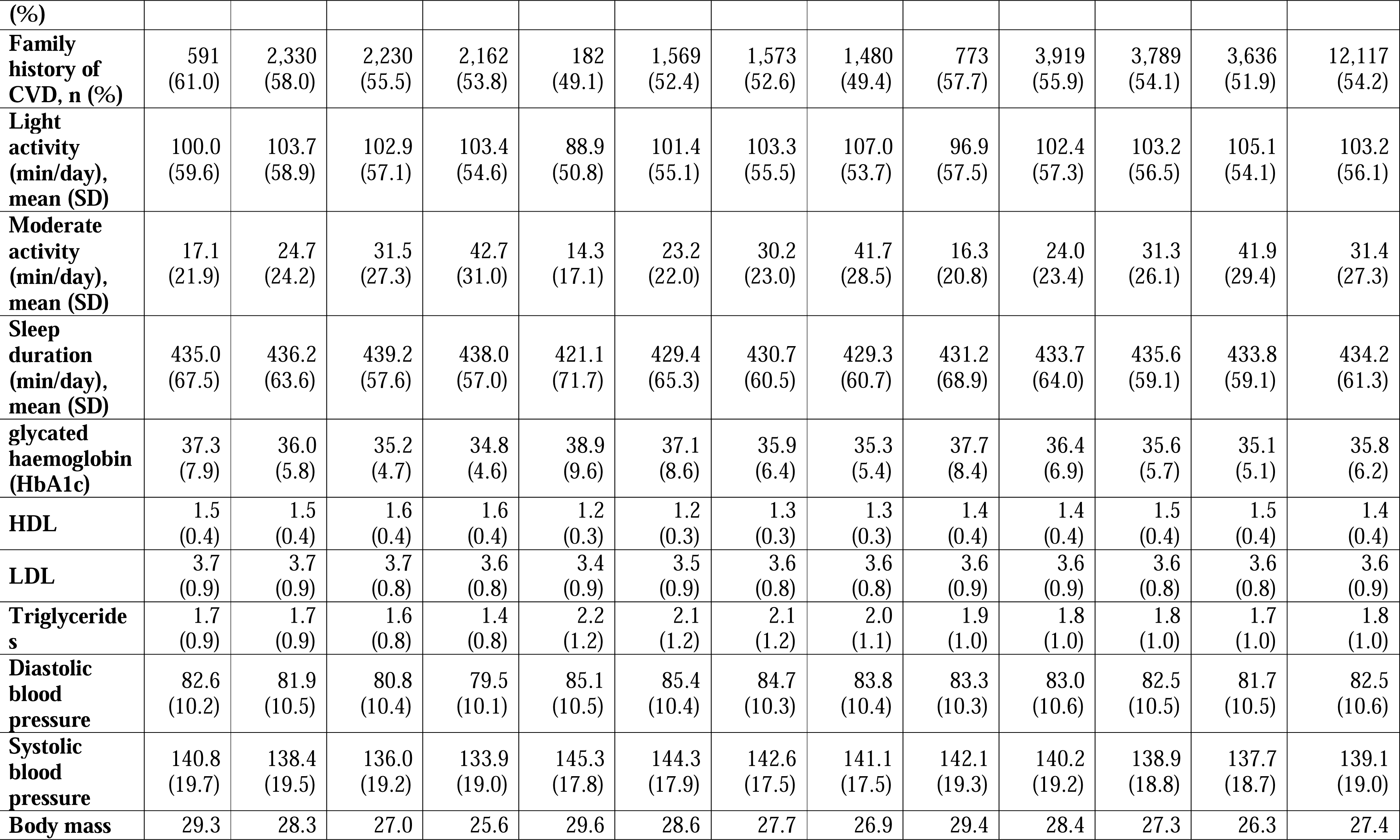
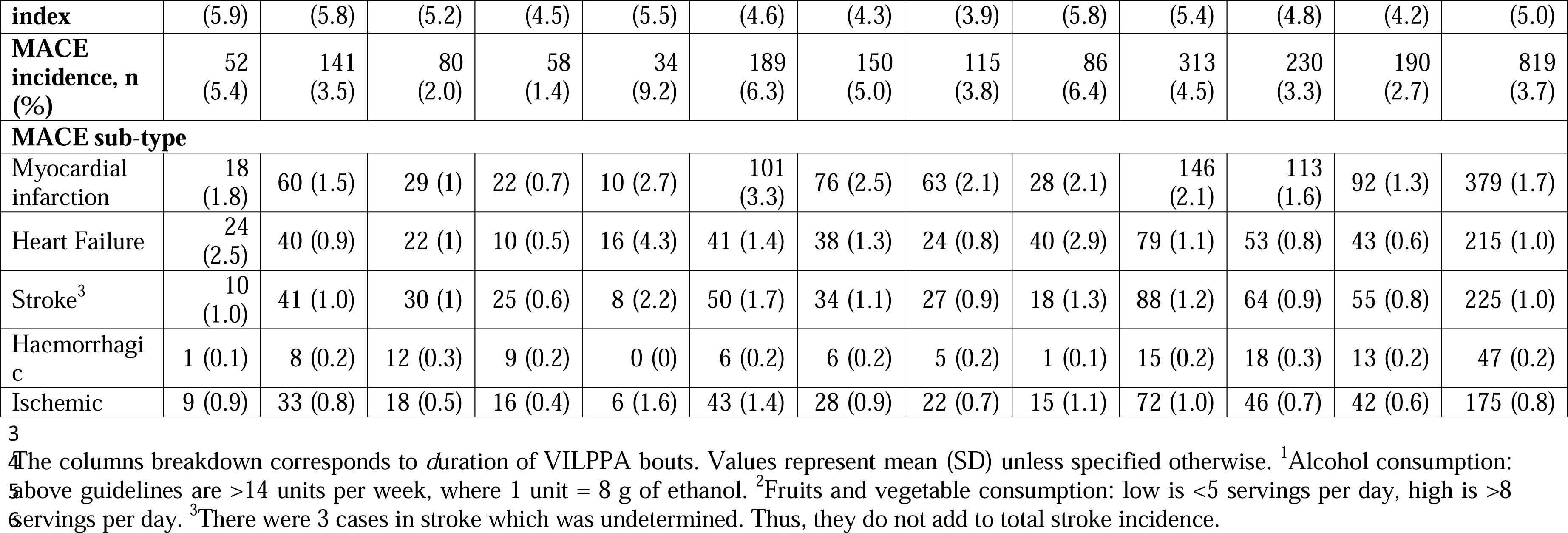
Participant baseline characteristics by VILPA duration and sex (non-exercisers, n= 22,368).

|  | Female |  |  |  | Male |  |  |  | Whole sample |  |  |  |  |
| --- | --- | --- | --- | --- | --- | --- | --- | --- | --- | --- | --- | --- | --- |
| Minutes | 0 | ≥0-2.5 | ≥2.5-7.5 | ≥7.5 | 0 | ≥0-2.5 | ≥2.5-7.5 | ≥7.5 | 0 | ≥0-2.5 | ≥2.5-7.5 | ≥7.5 | Overall |
| Sample size (n) | 969 | 4017 | 4016 | 4016 | 371 | 2993 | 2993 | 2993 | 1,340 | 7,010 | 7,009 | 7,009 | 22,368 |
| Follow up (in years), mean (SD) | 7.8<br>(1.2) | 7.9<br>(1.0) | 8.0<br>(0.9) | 8.0<br>(0.8) | 7.5<br>(1.5) | 7.8<br>(1.2) | 7.9<br>(1.1) | 7.9<br>(1.0) | 7.7<br>(1.3) | 7.9<br>(1.0) | 7.9<br>(1.0) | 7.9<br>(0.9) | 7.9<br>(1.0) |
| Age, mean (SD) | 64.8<br>(6.8) | 62.8<br>(7.5) | 60.9<br>(7.5) | 59.6<br>(7.4) | 65.7<br>(6.6) | 64.1<br>(7.3) | 62.4<br>(7.6) | 60.6<br>(7.8) | 65.0<br>(6.8) | 63.3<br>(7.4) | 61.5<br>(7.6) | 60.2<br>(7.6) | 61.9<br>(7.6) |
| Male, n (%) |  |  |  |  |  |  |  |  | 371<br>(27.7) | 2,314<br>(33.0) | 2,950<br>(42.1) | 3,715<br>(53.0) | 9,350<br>(41.8) |
| Ethnicity - White, n (%) | 937<br>(96.7) | 3,878<br>(96.5) | 3,850<br>(95.9) | 3,819<br>(95.1) | 365<br>(98.4) | 2,896<br>(96.8) | 2,891<br>(96.6) | 2,848<br>(95.2) | 1,302<br>(97.2) | 6,760<br>(96.4) | 6,751<br>(96.3) | 6,671<br>(95.2) | 21,484<br>(96.0) |
| Smoking history, n (%) |  |  |  |  |  |  |  |  |  |  |  |  |  |
| Current | 112<br>(11.6) | 347<br>(8.6) | 307<br>(7.6) | 265<br>(6.6) | 72<br>(19.4) | 340<br>(11.4) | 313<br>(10.5) | 274<br>(9.2) | 184<br>(13.7) | 659<br>(9.4) | 626<br>(8.9) | 561<br>(8.0) | 2,030<br>(9.1) |
| Never | 538<br>(55.5) | 2,355<br>(58.6) | 2,487<br>(61.9) | 2,510<br>(62.5) | 154<br>(41.5) | 1,455<br>(48.6) | 1,554<br>(51.9) | 1,631<br>(54.5) | 692<br>(51.6) | 3,883<br>(55.4) | 4,023<br>(57.4) | 4,086<br>(58.3) | 12,684<br>(56.7) |
| Previous | 319<br>(32.9) | 1,315<br>(32.7) | 1,222<br>(30.4) | 1,241<br>(30.9) | 145<br>(39.1) | 1,198<br>(40.0) | 1,126<br>(37.6) | 1,088<br>(36.4) | 464<br>(34.6) | 2,468<br>(35.2) | 2,360<br>(33.7) | 2,362<br>(33.7) | 7,654<br>(34.2) |
| Alcohol consumption, n (%)<sup>1</sup> |  |  |  |  |  |  |  |  |  |  |  |  |  |
| Never | 48<br>(5.0) | 220<br>(5.5) | 163<br>(4.1) | 185<br>(4.6) | 11<br>(3.0) | 76<br>(2.5) | 70<br>(2.3) | 71<br>(2.4) | 59<br>(4.4) | 313<br>(4.5) | 241<br>(3.4) | 231<br>(3.3) | 844<br>(3.8) |
| Ex-drinker | 45<br>(4.6) | 159<br>(4.0) | 145<br>(3.6) | 105<br>(2.6) | 12<br>(3.2) | 107<br>(3.6) | 70<br>(2.3) | 89<br>(3.0) | 57<br>(4.3) | 268<br>(3.8) | 219<br>(3.1) | 188<br>(2.7) | 732<br>(3.3) |
| Within guidelines | 667<br>(68.8) | 2,749<br>(68.4) | 2,779<br>(69.2) | 2,751<br>(68.5) | 187<br>(50.4) | 1,417<br>(47.3) | 1,394<br>(46.6) | 1,455<br>(48.6) | 854<br>(63.7) | 4,326<br>(61.7) | 4,167<br>(59.5) | 4,052<br>(57.8) | 13,399<br>(59.9) |
| Above guidelines | 209<br>(21.6) | 889<br>(22.1) | 929<br>(23.1) | 975<br>(24.3) | 161<br>(43.4) | 1,393<br>(46.5) | 1,459<br>(48.7) | 1,378<br>(46.0) | 370<br>(27.6) | 2,103<br>(30.0) | 2,382<br>(34.0) | 2,538<br>(36.2) | 7,393<br>(33.1) |
| **Education, n (%)** |  |  |  |  |  |  |  |  |  |  |  |  |  |
| College | 336<br>(34.7) | 1,411<br>(35.1) | 1,443<br>(35.9) | 1,493<br>(37.2) | 159<br>(42.9) | 1,215<br>(40.6) | 1,140<br>(38.1) | 1,111<br>(37.1) | 495<br>(36.9) | 2,589<br>(36.9) | 2,614<br>(37.3) | 2,610<br>(37.2) | 8,308<br>(37.1) |
| A/AS level | 135<br>(13.9) | 531<br>(13.2) | 554<br>(13.8) | 550<br>(13.7) | 47<br>(12.7) | 324<br>(10.8) | 349<br>(11.7) | 349<br>(11.7) | 182<br>(13.6) | 853<br>(12.2) | 916<br>(13.1) | 888<br>(12.7) | 2,839<br>(12.7) |
| O level | 223<br>(23.0) | 969<br>(24.1) | 973<br>(24.2) | 968<br>(24.1) | 60<br>(16.2) | 522<br>(17.4) | 602<br>(20.1) | 570<br>(19.0) | 283<br>(21.1) | 1,551<br>(22.1) | 1,538<br>(21.9) | 1,515<br>(21.6) | 4,887<br>(21.8) |
| CSE | 36<br>(3.7) | 196<br>(4.9) | 214<br>(5.3) | 249<br>(6.2) | 9<br>(2.4) | 125<br>(4.2) | 138<br>(4.6) | 206<br>(6.9) | 45<br>(3.4) | 328<br>(4.7) | 343<br>(4.9) | 457<br>(6.5) | 1,173<br>(5.2) |
| NVQ/HND/<br>HNC | 36<br>(3.7) | 152<br>(3.8) | 142<br>(3.5) | 131<br>(3.3) | 27<br>(7.3) | 273<br>(9.1) | 280<br>(9.4) | 304<br>(10.2) | 63<br>(4.7) | 397<br>(5.7) | 401<br>(5.7) | 484<br>(6.9) | 1,345<br>(6.0) |
| Other | 203<br>(20.9) | 758<br>(18.9) | 690<br>(17.2) | 625<br>(15.6) | 69<br>(18.6) | 534<br>(17.8) | 484<br>(16.2) | 453<br>(15.1) | 272<br>(20.3) | 1,292<br>(18.4) | 1,197<br>(17.1) | 1,055<br>(15.1) | 3,816<br>(17.1) |
| **Fruit and vegetable consumption, n (%)<sup>2</sup>** |  |  |  |  |  |  |  |  |  |  |  |  |  |
| Low | 404<br>(41.7) | 1,695<br>(42.2) | 1,677<br>(41.8) | 1,646<br>(41.0) | 158<br>(42.6) | 1,240<br>(41.4) | 1,223<br>(40.9) | 1,193<br>(39.9) | 444<br>(33.1) | 2,334<br>(33.3) | 2,361<br>(33.7) | 2,358<br>(33.6) | 7,497<br>(33.5) |
| Moderate | 310<br>(32.0) | 1,298<br>(32.3) | 1,296<br>(32.3) | 1,352<br>(33.7) | 97<br>(26.1) | 777<br>(26.0) | 788<br>(26.3) | 770<br>(25.7) | 485<br>(36.2) | 2,481<br>(35.4) | 2,522<br>(36.0) | 2,516<br>(35.9) | 8,004<br>(35.8) |
| High | 255<br>(26.3) | 1,024<br>(25.5) | 1,043<br>(26.0) | 1,018<br>(25.3) | 116<br>(31.3) | 976<br>(32.6) | 982<br>(32.8) | 1,030<br>(34.4) | 411<br>(30.7) | 2,195<br>(31.3) | 2,126<br>(30.3) | 2,135<br>(30.5) | 6,867<br>(30.7) |
| **Medication, n (%)** |  |  |  |  |  |  |  |  |  |  |  |  |  |
| Cholesterol | 184<br>(19.0) | 493<br>(12.3) | 369<br>(9.2) | 239<br>(6.0) | 102<br>(27.5) | 697<br>(23.3) | 541<br>(18.1) | 410<br>(13.7) | 286<br>(21.3) | 1,124<br>(16.0) | 903<br>(12.9) | 722<br>(10.3) | 3,035<br>(13.6) |
| Insulin | 15<br>(1.5) | 27<br>(0.7) | 22<br>(0.5) | 17<br>(0.4) | 7<br>(1.9) | 33<br>(1.1) | 23<br>(0.8) | 16<br>(0.5) | 22<br>(1.6) | 56<br>(0.8) | 47<br>(0.7) | 35<br>(0.5) | 160<br>(0.7) |
| Blood pressure | 263<br>(27.1) | 734<br>(18.3) | 526<br>(13.1) | 377<br>(9.4) | 114<br>(30.7) | 759<br>(25.4) | 634<br>(21.2) | 420<br>(14.0) | 377<br>(28.1) | 1,447<br>(20.6) | 1,158<br>(16.5) | 845<br>(12.1) | 3,827<br>(17.1) |
| **Diagnosed cancer, n** | 147<br>(15.2) | 552<br>(13.7) | 439<br>(10.9) | 381<br>(9.5) | 28<br>(7.5) | 249<br>(8.3) | 204<br>(6.8) | 150<br>(5.0) | 175<br>(13.1) | 826<br>(11.8) | 650<br>(9.3) | 499<br>(7.1) | 2,150<br>(9.6) |
| (%) |  |  |  |  |  |  |  |  |  |  |  |  |  |
| **Family history of CVD, n (%)** | 591<br>(61.0) | 2,330<br>(58.0) | 2,230<br>(55.5) | 2,162<br>(53.8) | 182<br>(49.1) | 1,569<br>(52.4) | 1,573<br>(52.6) | 1,480<br>(49.4) | 773<br>(57.7) | 3,919<br>(55.9) | 3,789<br>(54.1) | 3,636<br>(51.9) | 12,117<br>(54.2) |
| **Light activity (min/day), mean (SD)** | 100.0<br>(59.6) | 103.7<br>(58.9) | 102.9<br>(57.1) | 103.4<br>(54.6) | 88.9<br>(50.8) | 101.4<br>(55.1) | 103.3<br>(55.5) | 107.0<br>(53.7) | 96.9<br>(57.5) | 102.4<br>(57.3) | 103.2<br>(56.5) | 105.1<br>(54.1) | 103.2<br>(56.1) |
| **Moderate activity (min/day), mean (SD)** | 17.1<br>(21.9) | 24.7<br>(24.2) | 31.5<br>(27.3) | 42.7<br>(31.0) | 14.3<br>(17.1) | 23.2<br>(22.0) | 30.2<br>(23.0) | 41.7<br>(28.5) | 16.3<br>(20.8) | 24.0<br>(23.4) | 31.3<br>(26.1) | 41.9<br>(29.4) | 31.4<br>(27.3) |
| **Sleep duration (min/day), mean (SD)** | 435.0<br>(67.5) | 436.2<br>(63.6) | 439.2<br>(57.6) | 438.0<br>(57.0) | 421.1<br>(71.7) | 429.4<br>(65.3) | 430.7<br>(60.5) | 429.3<br>(60.7) | 431.2<br>(68.9) | 433.7<br>(64.0) | 435.6<br>(59.1) | 433.8<br>(59.1) | 434.2<br>(61.3) |
| **glycated haemoglobin (HbA1c)** | 37.3<br>(7.9) | 36.0<br>(5.8) | 35.2<br>(4.7) | 34.8<br>(4.6) | 38.9<br>(9.6) | 37.1<br>(8.6) | 35.9<br>(6.4) | 35.3<br>(5.4) | 37.7<br>(8.4) | 36.4<br>(6.9) | 35.6<br>(5.7) | 35.1<br>(5.1) | 35.8<br>(6.2) |
| **HDL** | 1.5<br>(0.4) | 1.5<br>(0.4) | 1.6<br>(0.4) | 1.6<br>(0.4) | 1.2<br>(0.3) | 1.2<br>(0.3) | 1.3<br>(0.3) | 1.3<br>(0.3) | 1.4<br>(0.4) | 1.4<br>(0.4) | 1.5<br>(0.4) | 1.5<br>(0.4) | 1.4<br>(0.4) |
| **LDL** | 3.7<br>(0.9) | 3.7<br>(0.9) | 3.7<br>(0.8) | 3.6<br>(0.8) | 3.4<br>(0.9) | 3.5<br>(0.9) | 3.6<br>(0.8) | 3.6<br>(0.8) | 3.6<br>(0.9) | 3.6<br>(0.9) | 3.6<br>(0.8) | 3.6<br>(0.8) | 3.6<br>(0.9) |
| **Triglyceride s** | 1.7<br>(0.9) | 1.7<br>(0.9) | 1.6<br>(0.8) | 1.4<br>(0.8) | 2.2<br>(1.2) | 2.1<br>(1.2) | 2.1<br>(1.2) | 2.0<br>(1.1) | 1.9<br>(1.0) | 1.8<br>(1.0) | 1.8<br>(1.0) | 1.7<br>(1.0) | 1.8<br>(1.0) |
| **Diastolic blood pressure** | 82.6<br>(10.2) | 81.9<br>(10.5) | 80.8<br>(10.4) | 79.5<br>(10.1) | 85.1<br>(10.5) | 85.4<br>(10.4) | 84.7<br>(10.3) | 83.8<br>(10.4) | 83.3<br>(10.3) | 83.0<br>(10.6) | 82.5<br>(10.5) | 81.7<br>(10.5) | 82.5<br>(10.6) |
| **Systolic blood pressure** | 140.8<br>(19.7) | 138.4<br>(19.5) | 136.0<br>(19.2) | 133.9<br>(19.0) | 145.3<br>(17.8) | 144.3<br>(17.9) | 142.6<br>(17.5) | 141.1<br>(17.5) | 142.1<br>(19.3) | 140.2<br>(19.2) | 138.9<br>(18.8) | 137.7<br>(18.7) | 139.1<br>(19.0) |
| **Body mass** | 29.3 | 28.3 | 27.0 | 25.6 | 29.6 | 28.6 | 27.7 | 26.9 | 29.4 | 28.4 | 27.3 | 26.3 | 27.4 |
| **index** | (5.9) | (5.8) | (5.2) | (4.5) | (5.5) | (4.6) | (4.3) | (3.9) | (5.8) | (5.4) | (4.8) | (4.2) | (5.0) |
| **MACE incidence, n (%)** | 52<br>(5.4) | 141<br>(3.5) | 80<br>(2.0) | 58<br>(1.4) | 34<br>(9.2) | 189<br>(6.3) | 150<br>(5.0) | 115<br>(3.8) | 86<br>(6.4) | 313<br>(4.5) | 230<br>(3.3) | 190<br>(2.7) | 819<br>(3.7) |
| **MACE sub-type** |  |  |  |  |  |  |  |  |  |  |  |  |  |
| Myocardial infarction | 18<br>(1.8) | 60 (1.5) | 29 (1) | 22 (0.7) | 10 (2.7) | 101<br>(3.3) | 76 (2.5) | 63 (2.1) | 28 (2.1) | 146<br>(2.1) | 113<br>(1.6) | 92 (1.3) | 379 (1.7) |
| Heart Failure | 24<br>(2.5) | 40 (0.9) | 22 (1) | 10 (0.5) | 16 (4.3) | 41 (1.4) | 38 (1.3) | 24 (0.8) | 40 (2.9) | 79 (1.1) | 53 (0.8) | 43 (0.6) | 215 (1.0) |
| Stroke<sup>3</sup> | 10<br>(1.0) | 41 (1.0) | 30 (1) | 25 (0.6) | 8 (2.2) | 50 (1.7) | 34 (1.1) | 27 (0.9) | 18 (1.3) | 88 (1.2) | 64 (0.9) | 55 (0.8) | 225 (1.0) |
| Haemorrhagic | 1 (0.1) | 8 (0.2) | 12 (0.3) | 9 (0.2) | 0 (0) | 6 (0.2) | 6 (0.2) | 5 (0.2) | 1 (0.1) | 15 (0.2) | 18 (0.3) | 13 (0.2) | 47 (0.2) |
| Ischemic | 9 (0.9) | 33 (0.8) | 18 (0.5) | 16 (0.4) | 6 (1.6) | 43 (1.4) | 28 (0.9) | 22 (0.7) | 15 (1.1) | 72 (1.0) | 46 (0.7) | 42 (0.6) | 175 (0.8) |

The columns breakdown corresponds to duration of VILPPA bouts. Values represent mean (SD) unless specified otherwise. <sup>1</sup>Alcohol consumption: above guidelines are >14 units per week, where 1 unit = 8 g of ethanol. <sup>2</sup>Fruits and vegetable consumption: low is <5 servings per day, high is >8 servings per day. <sup>3</sup>There were 3 cases in stroke which was undetermined. Thus, they do not add to total stroke incidence.

### Dose-response associations of VILPA with MACE and its subtypes

Absolute risk dose-response curves of VILPA for MACE and subtypes by sex are presented in **eFigure 3** (unadjusted) and **eFigure 4** (multivariable-adjusted). In analyses of female and male participants combined, daily VILPA duration from bouts lasting up to one minute (**Figure 1**) exhibited a near-linear dose-response association with MACE and myocardial infarction events, a steep L-shaped association with heart failure, and no association with stroke. However, there were substantial sex differences as the dose-response associations were evident only in women for MACE, myocardial infarction, and heart failure (**Figure 2**). For example, for MACE the median daily VILPA duration doses (3.4/5.6 minutes for females/males) were associated with HRs of 0.55 (0.41, 0.75) in females and 0.84 (0.63, 1.12) in males; for heart failure the HRs were 0.33 (0.18, 0.59) in females and 0.61 (0.35, 1.06) in males **(eTable 3)**. We found statistically significant sex*VILPA interactions MACE, myocardial infarction and heart failure, but not for stroke in either sex groups (**eTable 11**).

**Figure 1:**
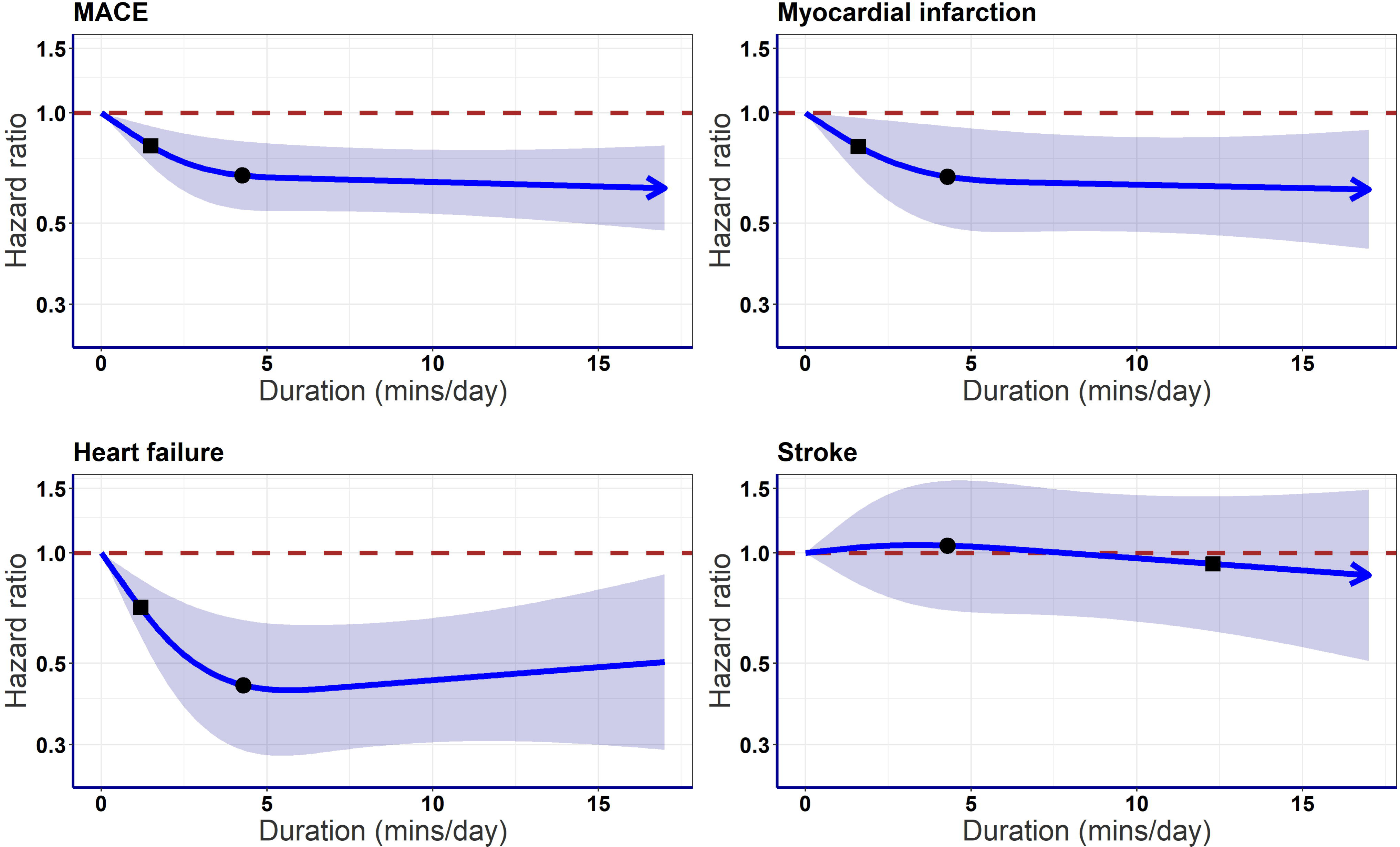
Adjusted dose response curves of daily VILPA duration for MACE and its subtypes, bouts lasting up to 1 minute (minutes/day). Adjusted for sex, age, light intensity, moderate intensity, VILPA bouts over 1-minute, smoking history, alcohol consumption, accelerometer estimated sleep duration, diet, education, ethnicity, self-reported parental history of CVD, previous incidence of cancer, and self-reported medication use (cholesterol, blood pressure, and diabetes). **Panel A:** all MACE: n = 22,368; events: 819, **Panel B:** myocardial infarction: n = 21,928; events = 379. **Panel C:** heart failure: n = 21,764; events = 215. **Panel D:** stroke: n = 21,774; events = 225. Diamond, minimal dose, as indicated by the ED_50_ statistic which estimates the daily duration of VILPA associated with 50% of optimal risk reduction. Circle, HR associated with the median VILPA value (see **eTable 3** for the list of values).

**Figure 2:**
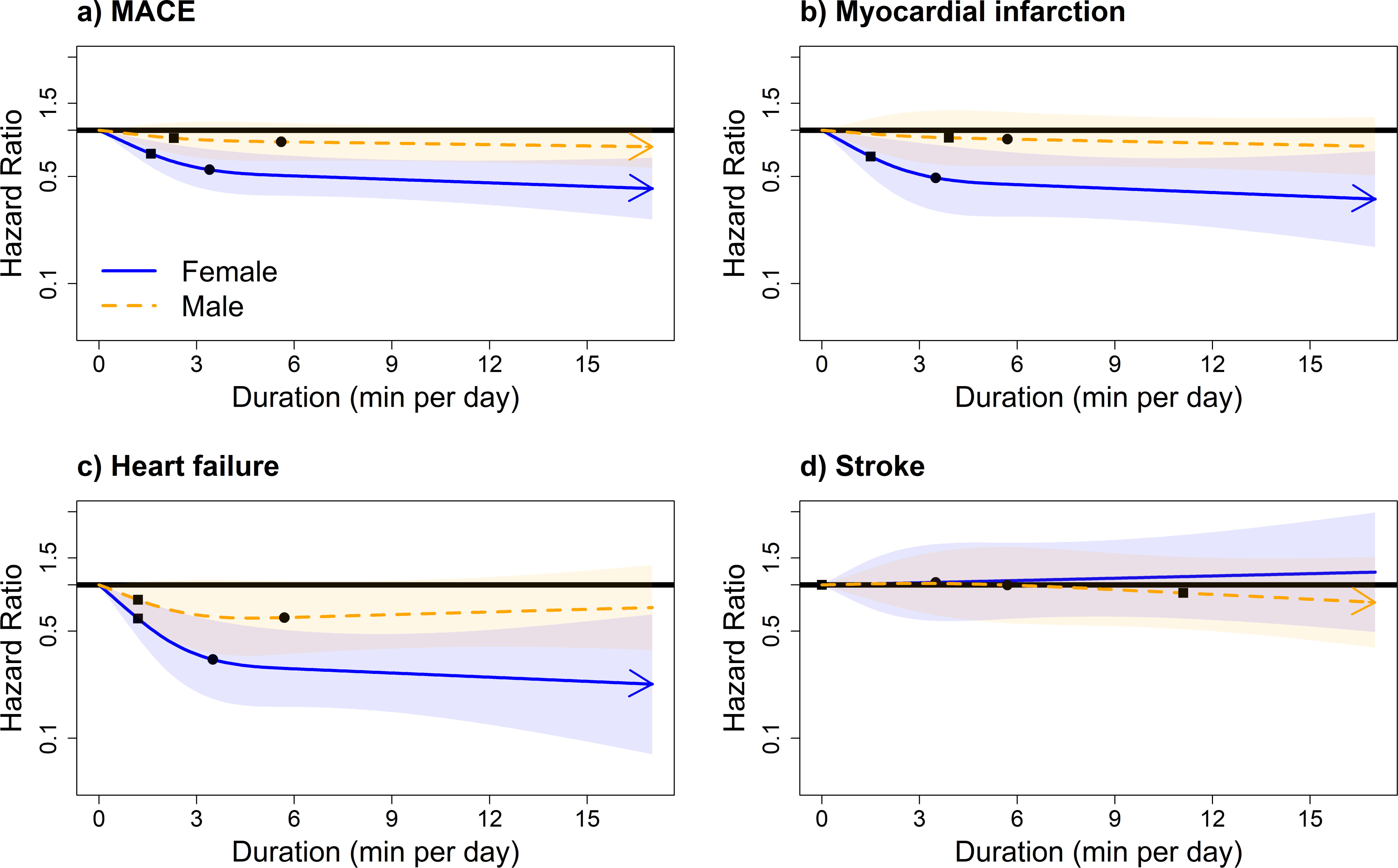
Sex-specific adjusted dose response curves of daily VILPA duration for MACE and its sub-types, bouts lasting up to 1 minute (minutes/day). Adjusted for age, light intensity, moderate intensity, VILPA bouts over 1-minute, smoking history, alcohol consumption, accelerometer estimated sleep duration, diet, education, ethnicity, self-reported parental history of CVD, previous incidence of cancer, and self-reported medication use (cholesterol, blood pressure, and diabetes). **Panel A:** all MACE: n = 22,368; events: 819 (female/male = 331/488), **Panel B:** myocardial infarction: n = 21,928; events = 379 (female/male = 129/250). **Panel C:** heart failure: n = 21,764; events = 215 (female/male = 96/119). **Panel D:** stroke: n = 21,774; events = 225 (female/male = 106/119). Diamond, minimal dose, as indicated by the ED_50_ statistic which estimates the daily duration of VILPA associated with 50% of optimal risk reduction. Circle, HR associated with the median VILPA value (see **eTable 4** for the list of values).

Length-standardised and raw daily VILPA bouts frequency showed similar dose-response patterns with VILPA duration across all outcomes **(eFigures 5-7**, **Figure 3)**. The median VILPA length-standardised bouts frequency (1.4/2.2 bouts per day in females/males) was associated with HRs of 0.56 (0.42, 0.75) in females and 0.83 (0.60, 1.10) in males for MACE; and HRs of 0.31 (0.18, 0.54) in females and 0.68 (0.40, 1.16) in males for heart failure. For MACE in females, the median raw frequency dose of 9.3 bouts per day was associated with an HR of 0.63 (95%CI: 0.46, 0.87). For MACE in males, the median raw frequency dose of 11.4 raw bouts per day was associated with an HR of 0.76 (95%CI: 0.56, 1.02) (**eTable 4)**. Dose-response analyses of daily duration for VILPA bouts lasting up to 2 minutes elicited very similar results to bouts lasting up to 1 minute (**eFigures 8-9**).

**Figure 3:**
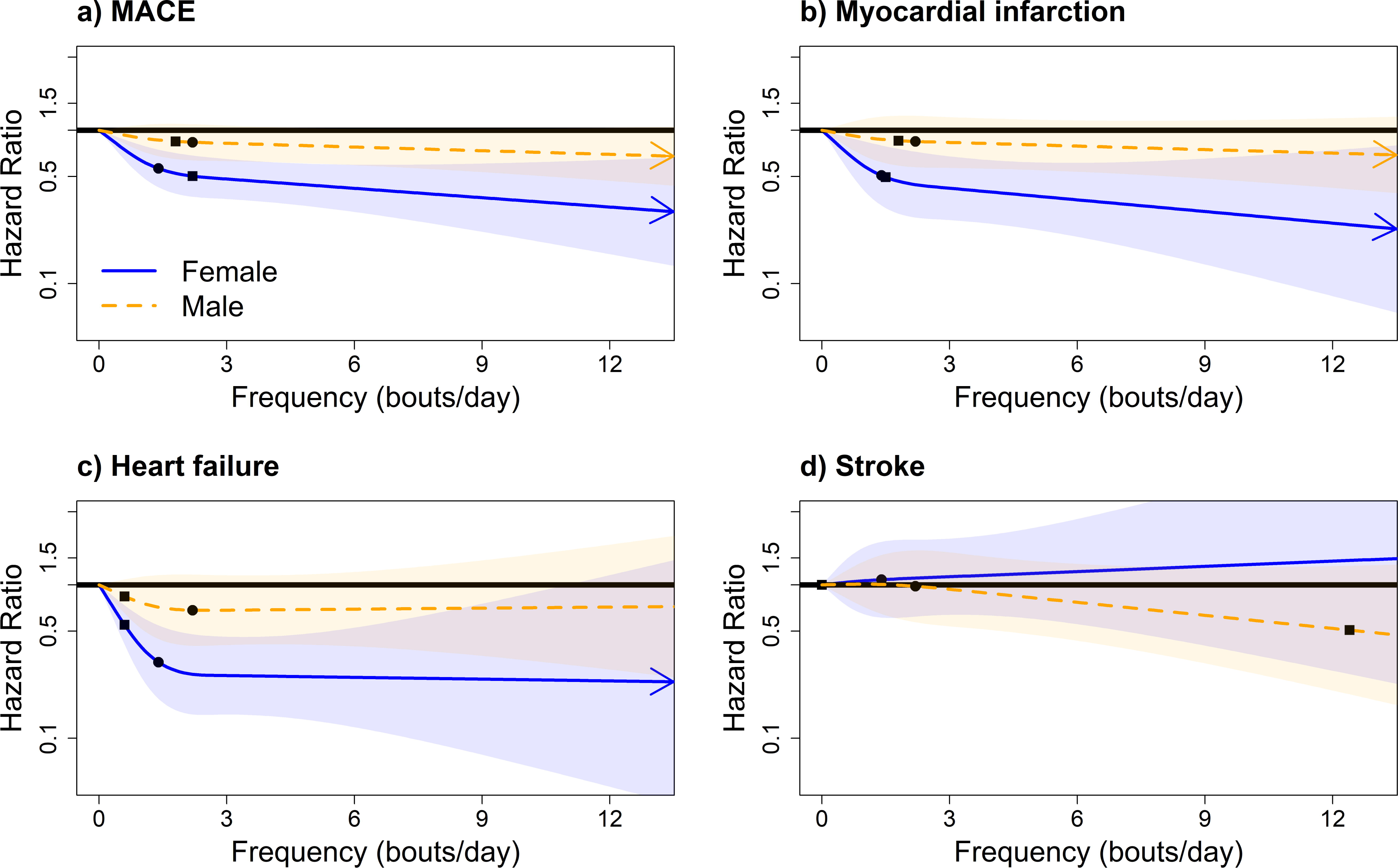
Sex-specific adjusted dose response curves for MACE and its subtypes by length-standardized VILPA frequency; bouts lasting up to 1Lmin (bouts/day). Adjusted for age, light intensity, moderate intensity, VILPA bouts over 1-minute, smoking history, alcohol consumption, accelerometer estimated sleep duration, diet, education, ethnicity, self-reported parental history of CVD, previous incidence of cancer, and self-reported medication use (cholesterol, blood pressure, and diabetes). The range was capped at the 97.5 percentile to minimize the influence of sparse data. **Panel A:** all MACE: n = 22,368; events: 819, **Panel B:** myocardial infarction: n = 21,928; events = 379. **Panel C:** heart failure: n = 21,764; events = 215. **Panel D:** stroke: n = 21,774; events = 225. Diamond, minimal dose, as indicated by the ED_50_ statistic which estimates the daily duration of VILPA associated with 50% of optimal risk reduction. Circle, HR associated with the median VILPA value (see **eTable 4** for the list of values).

### Minimum daily doses

**eTables 3** and **4** presents the HR and 95%CI associated with the minimum dose (eliciting 50% of the total effect)^4,5,20,21^, for VILPA bouts lasting up to 1 minute. For MACE in females and males, the minimum duration dose was 1.6 and 2.3 minutes per day, corresponding to HRs of 0.70 (0.58, 0.86) and 0.89 (0.70, 1.12), respectively. Findings were analogous for the minimum doses of myocardial infarction (1.5/3.9 minutes per day for female/male) and heart failure (1.2 minutes per day in both sex groups) which were statistically significant for females only (HRs of 0.67 (0.50, 0.91) and 0.60 (0.45, 0.81), respectively **(eTable 4).**

For MACE in females, the minimum frequency dose was 2.2 length standardised bouts and 9.6 raw bouts per day corresponding to a HR of 0.50 (0.37, 0.68) and 0.63 (0.46, 0.86), respectively. For MACE in males, the minimum frequency dose was 1.7 length standardised and 4.4 raw bouts per day corresponding to HRs of 0.85 (0.65, 1.10) and 0.86 (0.71, 1.03), respectively (**eTable 4).**

### Comparisons of dose-response associations in exercisers and non-exercisers

The range of average daily VPA in exercisers was considerably wider than that of VILPA in non-exercisers (0-45 vs 0 – 17 minutes per day), as well as the bout length (**eTable 2**). The associations of VPA with MACE, myocardial infarction and stroke in male and female exercisers combined were similar in shape and magnitude to the equivalent VILPA findings in non-exercisers (**Figure 1)**, while dose-response curves with heart failure was steeper in exercisers (**eFigure 10**). Among exercisers there were no major sex differences in the dose response associations of VPA with overall MACE, myocardial infarction or heart failure while there was evidence of a dose response association with stroke only in males (**Figure 4)**. Male exercisers’ median daily VPA duration value of 8.3 minutes was associated with an HR of 0.64 (0.44, 0.93) (**eTable 5 and 6)**. The results of the length-standardised **(eFigure 11**) and raw (**eFigure 12**) daily VPA frequency in exercisers were consistent with the equivalent VPA duration findings.

**Figure 4:**
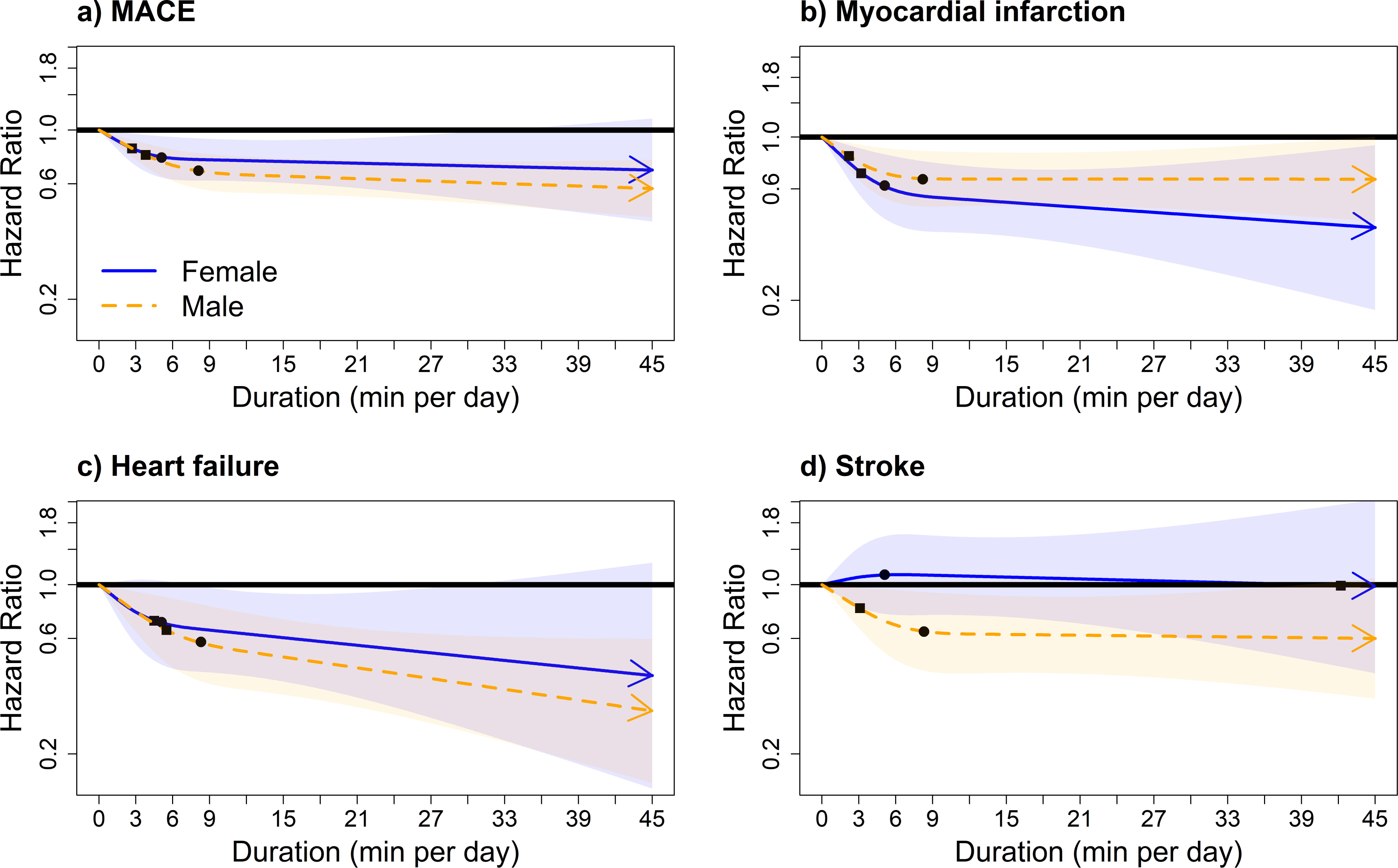
Adjusted sex-specific dose response curves of vigorous physical activity (VPA) in exercisers for MACE and its subtypes, bouts lasting up to 1 minute (minutes/day). Adjusted for sex, age, light intensity, moderate intensity, VILPA bouts over 1-minute, smoking history, alcohol consumption, accelerometer estimated sleep duration, diet, education, ethnicity, self-reported parental history of CVD, previous incidence of cancer, and self-reported medication use (cholesterol, blood pressure, and diabetes). The range was capped at the 97.5 percentile to minimize the influence of sparse data. **Panel A:** MACE: n = 58,648; events: 1854 (female/male = 749/1105), **Panel B:** myocardial infarction: n = 57,622; events = 828 (female/male = 287/541). **Panel C:** heart failure: n = 57,289; events = 495 (female/male = 210/285). **Panel D:** stroke: n = 57,325; events = 531 (female/male = 252/279). Diamond, minimal dose, as indicated by the ED_50_ statistic which estimates the daily duration of VILPA associated with 50% of optimal risk reduction. Circle, HR associated with the median VILPA value (see **eTable 6** for the list of values).

### Sensitivity analyses

All sensitivity analyses produced results consistent with the main findings **(eFigures 13-16 and eText 4). Additional analyses**

In the older age group (> 62.4 years, *n*= 11,673; 618 MACE events), the dose response associations of VILPA with MACE, myocardial infarction, and heart failure closely mirrored the sex-specific patterns in the whole sample of non-exercisers (**eFigure 17)**. In the younger age group (*n* = 10,695), associations were less clear, possibly due to the low number of events, The age*VILPA interaction tests were not statistically significant (**eTable 11**), possibly due to the same reason. We noted no major sex differences in the dose-response of daily light and moderate intensity physical activity with MACE and its subtypes **(eFigures 18-21)**. E-values indicated that for our estimates in females to be null, the association of an unmeasured confounder with VILPA duration exposures and outcomes should be a HR (lower 95% CI) of 2.21 (1.60) – 3.04 (2.00) for MACE; 2.34 (1.42) - 3.49 (1.80) for myocardial infarction; or 2.72 (1.77) – 5.51 (2.78) for heart failure. In males, for our estimates to be null, the association of an unmeasured confounder should be a HR (lower 95% CI) of 1.81 (1.00) – 2.66 (1.00) for heart failure **(eTable 7)**. In the subsample of 2,043 female and 1,588 male non-exercisers with valid accelerometry and fitness data (**eText 3**), the average absolute VO_2_ during VILPA bouts was 6.04 (1.02) MET for females and 6.21 (1.52) MET for males, corresponding to a relative intensity of 83.2 (18.2)% of VO_2_max for females and 70.5 (22.1)% of VO_2_max for males (**eTable 14).**

## DISCUSSION

Current clinical and public health guidelines assume similar cardiovascular responses to physical activity between sexes and offer no guidance on what quantity of incidental (non-exercise) activity has benefit. Our study, which uniquely examined sex-differences in the dose-response of incidental physical activity quantified by wearable trackers with major cardiovascular events, demonstrated a significant and linear dose-response association of VILPA with all MACE, myocardial infarction, and heart failure among women, but not men. The daily median duration of incidental physical activity of 3.4 minutes was associated with 45% lower hazards for MACE: 51% for myocardial infarction, and 67% for heart failure in women. Minimum doses of around 1.5 VILPA minutes per day (range 1.2-1.6 minutes) were associated with 30%, 33%, and 40% lower hazards for all-MACE, myocardial infarction and heart failure, respectively for women.

High-intensity interval training (HIIT)^23^ and proof-of-concept studies of intermittent stair climbing^7^ have shown bursts of VPA as brief as 20s to a few minutes in length, performed three to five times a day, can result in substantial improvements in cardiorespiratory fitness in previously inactive adults, providing a potential physiological basis^24^ for our findings. However, current literature does not shed light on the sex differences we observed since women are largely under-represented in HIIT trials^25^. Considering accelerometers record absolute intensity it is likely women’s VILPA bouts reflect higher relative loads compared to males, leading to more pronounced physiological adaptations in the long term (and the steeper dose-response associations in women. The metabolic, contractile and haemodynamic properties of skeletal muscle differ between males and females, which may moderate the response to the same absolute dose of vigorous exercise activities like VILPA^13^. These explanations are directly supported by our additional analyses (**eText 3**) where despite the very similar absolute energy expenditure of VILPA bouts between females and males (6.04-6.21 MET), the relative intensity during VILPA was substantially higher in females (83.2% (females) vs. 70.5% (males)) (**eTable 14).** Such relative intensity would categorise females’ but not males’ VILPA exertion as high intensity, as per current HIIT protocols^26,27^. The absence of sex differences in the dose response of light and moderate intensity physical activity (**eTables 16-19**) supports this interpretation as the overall level of exertion in these intensities is relatively modest and unlikely to elicit substantially different physiological responses between sex groups.

The sex-specific effects we observed were restricted to non-exercisers, suggesting a likely moderating role of the context in which vigorous physical activity is performed and bout characteristics. For example, vigorous bouts of any length (**eTable 2**) were approximately 30% longer for exercisers than non-exercisers, a pattern that likely reflects the voluntary and sustained effort involved in leisure-time exercise activities. One possible explanation of the more consistent dose-response in female and male exercisers is their vigorous bouts were longer and more likely to occur during activities designed for recreation and fitness. On the other hand, VILPA is more likely to be functional, opportunistic, and less voluntary (e.g., occupation, housework, or transportation). In our sub-group analyses (**eText 3**) women had 26% lower VO_2_max than men, confirming previous literature^24^. The higher relative effort required by women for a given incidental activity in daily living may explain, at least in part, the sex differences in VILPA dose-response.

### Strengths and limitations

We used device-based physical activity measurement and a validated^4–6^ two-stage machine learning-based intensity classifier. While we cannot rule out reverse causation bias and residual confounding, our results were robust to comprehensive sensitivity analyses and the E-values indicated that unmeasured confounding is unlikely to explain the observed associations. Some VILPA activities may not be fully captured by accelerometers (e.g., carrying a backpack), although such measurement error likely leads to underestimation of the “true” associations with MACE due to regression dilution bias^28^. There was a median lag of 5.5 years between the UK Biobank baseline when covariates and leisure-time physical activity measurements were taken and the accelerometry study. However, covariates have been shown to be stable over time^29^ and non-exerciser status a stable over time (82-88% stability)^4^. Accelerometry-measured physical activity is generally stable over time in adults (e.g., >90% of classification accuracy within 1 quartile over a period of 2-3 years)^30^. Although the UK Biobank had a low response rate (5.5%), recent empirical work has shown that cohort representativeness does not materially influence the associations between physical activity and cardiovascular outcomes in the UK Biobank^31^.

## Conclusions

Non-exercise vigorous incidental physical activity showed a beneficial dose response with MACE outcomes only in females, among whom very small amounts of VILPA (e.g., approximately 1.5 to 4 minutes per day) were associated with substantially lower risks of myocardial infarction and heart failure. While these findings are observational in nature, they suggest VILPA as a physical activity target for CVD prevention among non-exercising women. However, this pattern was not found in men among whom a recommendation of exercise-based vigorous intensity physical activity may still be needed. Our results support sex-specific physical activity guidelines for CVD prevention^14^. Our approach shows that wearable devices combined with machine learning based methods can reveal novel physical activity targets for CVD prevention and important sex differences.

## Funding

This study is funded by an Australian National Health and Medical Research Council (NHMRC) Investigator Grant (APP 1194510) and Ideas Grant (APP1180812). The funder had no specific role in any of the following study aspects: the design and conduct of the study; collection, management, analysis, and interpretation of the data; preparation, review, or approval of the manuscript; and decision to submit the manuscript for publication.

## Acknowledgements

This research has been conducted using the UK Biobank Resource under Application Number 25813. The authors would like to thank all the participants and professionals contributing to the UK Biobank. All information and materials in the manuscript are original and have not need submitted for publication elsewhere.

## Competing interests

The authors do not have any competing interests to declare.

## Data availability statement

The data that support the findings of this study are available from the UK Biobank, but restrictions apply to the availability of these data, which were used under license for the current study, and so are not publicly available. Data are however available from the authors upon reasonable request and with permission of the UK Biobank. ES and MA had full access to all the data in the study and takes responsibility for the integrity of the data and the accuracy of the data analysis.

## Code availability statement

The statistical code used in the analyses of this manuscript is available upon reasonable request.

## Abbreviations

MACE: major adverse cardiovascular events
CVD: cardiovascular disease
VILPA: vigorous intermittent lifestyle physical activity

## Notes

### Competing Interest Statement

The authors have declared no competing interest.

### Author Declarations

The UK National Research Ethics Service gave ethical approval for this work.

