## Supplement for "Device-measured vigorous intermittent lifestyle physical activity and major cardiovascular events"

Supplemental material

Dose-response of device-measured vigorous intermittent lifestyle physical activity and major adverse cardiovascular events in UK women and men

**Supplemental Material – ToC**

**Item**

**eTable 1:** Exercisers’ baseline characteristics by duration of VPA bouts and sex (n= 58,648).

**eTable 2:** mean and median bout length of VILPA (non-exercisers, n= 22,368) and VPA (exercisers, n= 58,648).

**eTable 3:** Hazard ratios associated with the minimum dose and median VILPA values among non-exercisers for bouts lasting up to 1 minute (n= 22,368).

**eTable 4:** Sex-specific hazard ratios associated with the minimum dose and median VPA values among non-exercisers for bouts lasting up to 1 minute (females n = 13018; males n= 9350).

**eTable 5**: Hazard ratios associated with the minimum dose and median VPA values among exercisers for bouts lasting up to 1 minute.

**eTable 6:** Sex-specific hazard ratios associated with the minimum dose and median VPA values among exercisers for bouts lasting up to 1 minute.

**eTable 7:** Sex-specific e-values for minimum dose and median VILPA values for MACE and its subtypes among non-exercisers for bouts lasting up to 1 minute.

**eTable 8:** Sex-specific e-values for minimum dose and median VPA values for MACE and its subtypes among exercisers for bouts lasting up to 1 minute.

**eTable 9:** Covariate definitions.

**eTable 10:** Interaction test between age and daily VILPA duration bouts lasting up to 1 minute (minutes/day) for MACE and its subtypes.

**eTable 11:** Interaction test between sex and daily VILPA duration bouts lasting up to 1 minute (minutes/day) for MACE and its subtypes.

**eTable 12:** Bivariate associations between VILPA duration and frailty index between men and women.

**eTable 13:** STROBE Statement.

**eTable 14:** Estimated maximal oxygen consumption, and average oxygen consumption and relative intensity of VILPA bouts (up to 1 minute) among 1,588 males non-exercisers UK Biobank participants with valid accelerometry and ergometer data (**eText 3).**

**eFigure 1:** Flow diagram of non-exerciser participants.

**eFigure 2:** Flow diagram of exercisers participants.

**eFigure 3:** Unadjusted absolute risk estimates for MACE and its subtypes by daily VILPA duration, bouts lasting up to 1 minute (minutes/day).

**eFigure 4:** Sex-specific adjusted absolute risk estimates of daily VILPA duration for MACE and its subtypes, bouts lasting up to 1 minute (minutes/day).

**eFigure 5**: Adjusted dose response curves of **length-standardised frequenc**y for MACE and its sub-types, VILPA bouts equivalent to 1 minute (bouts/day).

**eFigure 6**: Adjusted dose response curves of **raw VILPA frequency** for MACE and its sub-types, VILPA bouts lasting up to 1 minutes (bouts/day).

**eFigure 7**: Adjusted sex-specific dose response curves of **raw VILPA frequency** for MACE and its sub-types, VILPA bouts lasting up to 1 minutes (bouts/day).

**eFigure 8**: Adjusted dose response curves of daily VILPA duration for MACE and its sub-types, bouts lasting up to 2 minutes (minutes/day).

**eFigure 9**: Adjusted sex-specific dose response curves of daily VILPA duration for MACE and its sub-types, bouts lasting up to 2 minutes (minutes/day).

**eFigure 10**: Adjusted dose response curves of vigorous physical activity (VPA) in exercisers for MACE and its subtypes for bouts lasting up to 1 minute (minutes/day).

**eFigure 11:** Sex-specific adjusted dose response curves of **length-standardised frequency** of vigorous physical activity (VPA) in exercisers for MACE and its subtypes for bouts lasting up to 1 minute (minutes/day).

**eFigure 12:** Sex-specific adjusted dose response curves of **raw frequency** of vigorous physical activity (VPA) in exercisers for MACE and its subtypes for bouts lasting up to 1 minute (minutes/day).

**eFigure 13**: Adjusted Sex-specific dose response curves of daily VILPA duration for MACE and its subtypes, with additional adjustment for glycated haemoglobin (HbA1c), low-density lipoprotein (LDL), high-density lipoprotein (HDL), triglycerides, systolic blood pressure, diastolic blood pressure, and body mass index; bouts lasting up to 1 minute (minutes/day).

**eFigure 14**: Adjusted Sex-specific dose response curves of daily VILPA duration for MACE and its subtypes, after excluding participants with poor health or a BMI below 18.5 kg/m2 or current smokers.

**eFigure 15:** Adjusted sex-specific dose response curves of VILPA duration for MACE and its sub-types, VILPA bouts lasting up to 1 minutes (bouts/day), with reference as 15^th^ percentile for VILPA duration (0.625 mins/day).

**eFigure 16:** Adjusted sex-specific dose response curves of VILPA duration for MACE and its sub-types, VILPA bouts lasting up to 1 minutes (bouts/day), excluding frail participants (n = 1,924).

**eFigure 17:** Age-stratified and sex-specific dose response curves of daily VILPA duration for MACE and its subtypes for bouts lasting up to 1 minute (minutes/day).

**eFigure 18:** Adjusted Sex-specific dose response curves of light intensity physical activity for MACE and its subtypes for non-exercisers.

**eFigure 19:** Adjusted Sex-specific dose response curves of moderate intensity physical activity for MACE and its subtypes for non-exercisers.

**eFigure 20:** Adjusted Sex-specific dose response curves of light intensity physical activity for MACE and its subtypes for exercisers.

**eFigure 21:** Adjusted Sex-specific dose response curves of moderate intensity physical activity for MACE and its subtypes for exercisers.

**eText 1:** Physical activity classification.

**eText 2:** Bout length standardisation.

**eText 3:** Physical activity energy expenditure, cardiorespiratory fitness, and relative physical activity intensity.

**eText 4:** Unabridged sections of summarised results in main manuscript.

**eTable 1:** Exercisers’ baseline characteristics by VILPA duration and sex (n= 58,648)

|  | **Duration of VPA** | | | | | | | | | | | | |
| --- | --- | --- | --- | --- | --- | --- | --- | --- | --- | --- | --- | --- | --- |
|  | **Female** | | | | **Male** | | | | **Whole sample** | | | | |
| **Minutes** | **0** | $\boldsymbol{\geq}$**0-2.5** | $\boldsymbol{\geq}$**2.5-7.5** | $\boldsymbol{\geq}$**7.5** | **0** | $\boldsymbol{\geq}$**0-2.5** | $\boldsymbol{\geq}$**2.5-7.5** | $\boldsymbol{\geq}$**7.5** | **0** | $\boldsymbol{\geq}$**0-2.5** | $\boldsymbol{\geq}$**2.5-7.5** | $\boldsymbol{\geq}$**7.5** | **Overall** |
| **Sample size (n)** | 1,628 | 10,912 | 10,912 | 10,912 | 523 | 7,921 | 7,920 | 7,920 | 2,151 | 18,833 | 18,832 | 18,832 | 58,648 |
| **Follow up (in years), mean (SD)** | 7.9 (1.1) | 8.0 (0.9) | 8.0 (0.8) | 8.0 (0.8) | 7.7 (1.3) | 7.8 (1.2) | 7.9 (1.0) | 8.0 (1.0) | 7.8 (1.1) | 7.9 (1.0) | 7.9 (0.9) | 8.0 (0.9) | 7.9 (1.0) |
| **Age, mean (SD)** | 65.3 (7.1) | 62.9 (7.5) | 60.7 (7.6) | 58.5 (7.5) | 66.4 (6.8) | 63.7 (7.7) | 61.5 (8.0) | 59.4 (7.9) | 65.6 (7.0) | 63.1 (7.6) | 61.0 (7.8) | 59.1 (7.7) | 61.2 (7.9) |
| **Male, n (%)** | | | | | | | | | 523 (24.3) | 6,050 (32.1) | 7,915 (42.0) | 9,796 (52.0) | 24,284 (41.4) |
| **Ethnicity - White, n (%)** | 1,577 (96.9) | 10,614 (97.3) | 10,597 (97.1) | 10,496 (96.2) | 513 (98.1) | 7,716 (97.4) | 7,673 (96.9) | 7,622 (96.2) | 2,090 (97.2) | 18,320 (97.3) | 18,272 (97.0) | 18,126 (96.3) | 56,808 (96.9) |
| **Smoking history, n (%)** | | | | | | | | | | | | | |
| Current | 96 (5.9) | 595 (5.5) | 533 (4.9) | 474 (4.4) | 73 (14.1) | 614 (7.8) | 541 (6.8) | 473 (6.0) | 169 (7.9) | 1,177 (6.3) | 1,071 (5.7) | 982 (5.2) | 3,399 (5.8) |
| Never | 990 (61.0) | 6,597 (60.6) | 6,672 (61.3) | 6,880 (63.2) | 244 (47.0) | 4,058 (51.3) | 4,376 (55.4) | 4,668 (59.0) | 1,234 (57.6) | 10,805 (57.5) | 10,987 (58.5) | 11,459 (60.9) | 34,485 (58.9) |
| Previous | 537 (33.1) | 3,694 (33.9) | 3,688 (33.9) | 3,540 (32.5) | 202 (38.9) | 3,231 (40.9) | 2,985 (37.8) | 2,767 (35.0) | 739 (34.5) | 6,809 (36.2) | 6,735 (35.8) | 6,361 (33.8) | 20,644 (35.3) |
| **Alcohol consumption, n (%)^1^** | | | | | | | | | | | | | |
| Never | 73 (4.5) | 379 (3.5) | 292 (2.7) | 333 (3.1) | 12 (2.3) | 123 (1.6) | 120 (1.5) | 107 (1.4) | 85 (4.0) | 534 (2.9) | 395 (2.1) | 425 (2.3) | 1,439 (2.5) |
| Ex-drinker | 60 (3.7) | 288 (2.7) | 246 (2.3) | 214 (2.0) | 13 (2.5) | 198 (2.5) | 180 (2.3) | 167 (2.1) | 73 (3.4) | 490 (2.6) | 413 (2.2) | 390 (2.1) | 1,366 (2.3) |
| Within guidelines | 1,129 (69.6) | 7,305 (67.4) | 7,281 (67.0) | 7,269 (66.9) | 235 (45.3) | 3,305 (41.9) | 3,229 (40.9) | 3,352 (42.5) | 1,364 (63.7) | 11,042 (59.0) | 10,576 (56.4) | 10,123 (54.0) | 33,105 (56.7) |
| Above guidelines | 361 (22.2) | 2,874 (26.5) | 3,044 (28.0) | 3,048 (28.1) | 259 (49.9) | 4,259 (54.0) | 4,360 (55.3) | 4,268 (54.1) | 620 (28.9) | 6,665 (35.6) | 7,370 (39.3) | 7,818 (41.7) | 22,473 (38.5) |
| **Education, n (%)** | | | | | | | | | | | | | |
| College | 657 (40.4) | 4,764 (43.7) | 4,920 (45.1) | 5,247 (48.1) | 218 (41.7) | 3,834 (48.4) | 3,815 (48.2) | 3,962 (50.0) | 875 (40.7) | 8,537 (45.3) | 8,779 (46.6) | 9,226 (49.0) | 27,417 (46.7) |
| A/AS level | 191 (11.7) | 1,541 (14.1) | 1,621 (14.9) | 1,581 (14.5) | 79 (15.1) | 917 (11.6) | 999 (12.6) | 997 (12.6) | 270 (12.6) | 2,515 (13.4) | 2,597 (13.8) | 2,544 (13.5) | 7,926 (13.5) |
| O level | 381 (23.4) | 2,408 (22.1) | 2,321 (21.3) | 2,264 (20.7) | 87 (16.6) | 1,329 (16.8) | 1,365 (17.2) | 1,360 (17.2) | 468 (21.8) | 3,842 (20.4) | 3,609 (19.2) | 3,596 (19.1) | 11,515 (19.6) |
| CSE | 40 (2.5) | 342 (3.1) | 400 (3.7) | 420 (3.8) | 13 (2.5) | 227 (2.9) | 285 (3.6) | 338 (4.3) | 53 (2.5) | 557 (3.0) | 699 (3.7) | 756 (4.0) | 2,065 (3.5) |
| NVQ/HND/HNC | 64 (3.9) | 331 (3.0) | 340 (3.1) | 289 (2.6) | 41 (7.8) | 595 (7.5) | 566 (7.1) | 555 (7.0) | 105 (4.9) | 843 (4.5) | 908 (4.8) | 925 (4.9) | 2,781 (4.7) |
| Other | 295 (18.1) | 1,526 (14.0) | 1,310 (12.0) | 1,111 (10.2) | 85 (16.3) | 1,019 (12.9) | 890 (11.2) | 708 (8.9) | 380 (17.7) | 2,539 (13.5) | 2,240 (11.9) | 1,785 (9.5) | 6,944 (11.8) |
| **Fruit and vegetable consumption, n (%)^2^** | | | | | | | | | | | | | |
| Low | 696 (43.4) | 4,686 (43.4) | 4,721 (43.7) | 4,592 (42.5) | 254 (49.5) | 3,464 (44.4) | 3,337 (42.8) | 3,254 (41.5) | 725 (34.2) | 6,357 (34.1) | 6,708 (36.0) | 6,620 (35.5) | 20,410 (35.2) |
| Moderate | 510 (31.8) | 3,384 (31.3) | 3,301 (30.5) | 3,292 (30.5) | 121 (23.6) | 1,851 (23.7) | 1,934 (24.8) | 1,866 (23.8) | 753 (35.6) | 6,486 (34.8) | 6,223 (33.4) | 6,225 (33.4) | 19,687 (33.9) |
| High | 398 (24.8) | 2,729 (25.3) | 2,793 (25.8) | 2,927 (27.1) | 138 (26.9) | 2,495 (31.9) | 2,533 (32.5) | 2,723 (34.7) | 639 (30.2) | 5,772 (31.0) | 5,695 (30.6) | 5,796 (31.1) | 17,902 (30.9) |
| **Medication, n (%)** | | | | | | | | | | | | | |
| Cholesterol | 222 (13.6) | 1,111 (10.2) | 716 (6.6) | 440 (4.0) | 139 (26.6) | 1,557 (19.7) | 1,088 (13.7) | 757 (9.6) | 361 (16.8) | 2,500 (13.3) | 1,821 (9.7) | 1,348 (7.2) | 6,030 (10.3) |
| Insulin | 8 (0.5) | 59 (0.5) | 28 (0.3) | 42 (0.4) | 5 (1.0) | 70 (0.9) | 47 (0.6) | 35 (0.4) | 13 (0.6) | 120 (0.6) | 87 (0.5) | 74 (0.4) | 294 (0.5) |
| Blood pressure | 353 (21.7) | 1,601 (14.7) | 1,060 (9.7) | 696 (6.4) | 164 (31.4) | 1,704 (21.5) | 1,165 (14.7) | 767 (9.7) | 517 (24.0) | 3,208 (17.0) | 2,214 (11.8) | 1,571 (8.3) | 7,510 (12.8) |
| **Diagnosed cancer, n (%)** | 243 (14.9) | 1,352 (12.4) | 1,148 (10.5) | 994 (9.1) | 52 (9.9) | 603 (7.6) | 478 (6.0) | 356 (4.5) | 295 (13.7) | 2,025 (10.8) | 1,636 (8.7) | 1,270 (6.7) | 5,226 (8.9) |
| **Family history of CVD, n (%)** | 999 (61.4) | 6,403 (58.7) | 5,976 (54.8) | 5,665 (51.9) | 286 (54.7) | 4,258 (53.8) | 4,101 (51.8) | 3,869 (48.9) | 1,285 (59.7) | 10,744 (57.0) | 10,002 (53.1) | 9,526 (50.6) | 31,557 (53.8) |
| **Light activity (min/day), mean (SD)** | 102.2 (59.3) | 106.5 (58.5) | 106.5 (57.5) | 109.4 (55.2) | 95.3 (52.9) | 103.0 (53.8) | 105.1 (53.4) | 110.3 (52.3) | 100.6 (57.9) | 105.4 (56.9) | 105.5 (55.6) | 109.8 (53.8) | 106.7 (55.6) |
| **Moderate activity (min/day),**  **mean (SD)** | 18.7 (21.7) | 27.3 (24.7) | 36.6 (28.0) | 47.4 (31.8) | 16.5 (18.6) | 27.0 (22.2) | 35.7 (24.8) | 44.9 (28.5) | 18.2 (21.0) | 27.4 (24.3) | 36.2 (26.4) | 46.1 (30.4) | 35.9 (28.2) |
| **Sleep (min/day),**  **mean (SD)** | 433.4 (64.3) | 437.9 (59.1) | 440.2 (56.2) | 438.8 (52.9) | 432.8 (65.2) | 431.0 (62.6) | 430.9 (59.2) | 431.6 (54.9) | 433.2 (64.5) | 436.0 (60.4) | 435.9 (57.8) | 435.2 (54.1) | 435.6 (57.8) |
| **glycated haemoglobin (HbA1c)** | 35.9 (5.1) | 35.3 (4.8) | 34.6 (4.2) | 34.3 (4.0) | 37.4 (8.4) | 35.7 (6.3) | 34.9 (5.3) | 34.4 (4.4) | 36.3 (6.1) | 35.5 (5.3) | 34.8 (4.8) | 34.4 (4.2) | 34.9 (4.9) |
| **HDL** | 1.6 (0.4) | 1.6 (0.4) | 1.7 (0.4) | 1.7 (0.4) | 1.3 (0.3) | 1.3 (0.3) | 1.3 (0.3) | 1.4 (0.3) | 1.5 (0.4) | 1.5 (0.4) | 1.5 (0.4) | 1.5 (0.4) | 1.5 (0.4) |
| **LDL** | 3.7 (0.9) | 3.7 (0.9) | 3.6 (0.8) | 3.5 (0.8) | 3.5 (0.9) | 3.5 (0.8) | 3.6 (0.8) | 3.6 (0.8) | 3.7 (0.9) | 3.6 (0.9) | 3.6 (0.8) | 3.5 (0.8) | 3.6 (0.8) |
| **Triglycerides** | 1.6 (0.9) | 1.5 (0.8) | 1.4 (0.8) | 1.3 (0.7) | 2.1 (1.1) | 2.0 (1.1) | 1.9 (1.1) | 1.7 (1.0) | 1.7 (1.0) | 1.7 (0.9) | 1.6 (0.9) | 1.5 (0.9) | 1.6 (0.9) |
| **Diastolic blood pressure** | 82.0 (10.3) | 80.8 (10.3) | 79.5 (10.4) | 78.5 (10.3) | 85.1 (10.6) | 84.3 (10.2) | 83.6 (10.2) | 82.5 (10.2) | 82.7 (10.5) | 81.9 (10.5) | 81.2 (10.6) | 80.6 (10.4) | 81.3 (10.5) |
| **Systolic blood pressure** | 141.2 (20.5) | 137.5 (19.9) | 134.5 (19.6) | 132.1 (18.9) | 146.1 (18.9) | 143.4 (18.2) | 141.6 (17.7) | 139.7 (17.5) | 142.4 (20.2) | 139.4 (19.6) | 137.3 (19.1) | 136.1 (18.7) | 137.8 (19.2) |
| **Body mass index** | 28.1 (5.3) | 26.8 (4.7) | 25.6 (4.2) | 24.5 (3.7) | 29.1 (4.4) | 27.6 (4.0) | 26.8 (3.5) | 25.9 (3.2) | 28.4 (5.1) | 27.0 (4.5) | 26.0 (4.0) | 25.2 (3.6) | 26.2 (4.2) |
| **MACE incidence, n (%)** | 66 (4.1) | 330 (3.0) | 198 (1.8) | 155 (1.4) | 47 (9.0) | 511 (6.5) | 304 (3.8) | 243 (3.1) | 113 (5.3) | 762 (4.0) | 550 (2.9) | 429 (2.3) | 1,854 (3.2) |
| Myocardial infarction | 27 (1.7) | 133 (1.0) | 69 (0.6) | 58 (0.5) | 23 (4.4) | 243 (3.1) | 153 (1.9) | 122 (1.5) | 50 (2.3) | 332 (1.8) | 249 (1.3) | 197 (1.0) | 828 (1.4) |
| Heart Failure | 19 (1.2) | 100 (0.9) | 56 (0.5) | 35 (0.3) | 14 (2.7) | 149 (1.9) | 74 (0.9) | 48 (0.6) | 33 (1.5) | 223 (1.1) | 146 (0.8) | 93 (0.5) | 495 (0.9) |
| Stroke^3^ | 20 (1.2) | 97 (0.9) | 75 (0.7) | 60 (0.5) | 10 (1.9) | 128 (1.6) | 71 (0.9) | 70 (0.9) | 30 (1.4) | 207 (1.1) | 155 (0.8) | 139 (0.7) | 531 (0.9) |
| Haemorrhagic | 6 (0.4) | 31 (0.3) | 16 (0.1) | 15 (0.1) | 1 (0.2) | 31 (0.4) | 14 (0.2) | 21 (0.3) | 7 (0.3) | 57 (0.3) | 32 (0.2) | 39 (0.2) | 135 (0.2) |
| Ischemic | 13 (0.8) | 66 (0.6) | 62 (0.6) | 44 (0.4) | 9 (1.7) | 98 (1.2) | 57 (0.6) | 51 (0.6) | 22 (1.0) | 149 (0.8) | 122 (0.6) | 100 (0.5) | 393 (0.7) |

The columns breakdown corresponds to *d*uration of VILPPA bouts. Values represent mean (SD) unless specified otherwise. ^1^Alcohol consumption: above guidelines is >14 units per week, where 1 unit = 8 g of ethanol. ^2^Fruits and vegetable consumption: low is <5 servings per day, high is >8 servings per day. ^3^There were 3 cases in stroke which was undetermined. Thus, they do not add to total stroke incidence.

**eTable 2:** mean and median bout length of VILPA (non-exercisers, n= 22,368) and VPA (exercisers, n= 58,648)

|  | Median | Mean | SD |
| --- | --- | --- | --- |
| VILPA |  |  |  |
| Up to 1 min | 20 | 27.6 | 15.3 |
| Up to 2 min | 30 | 34.9 | 24.7 |
| Over 2 min | 190 | 222.2 | 307.1 |
| VPA |  |  |  |
| Up to 1 min | 30 | 39.1 | 15.8 |
| Up to 2 min | 30 | 49.6 | 27.7 |
| Over 2 min | 230 | 332.4 | 447.1 |

1) Males; values represent seconds:

|  | Median | Mean | SD |
| --- | --- | --- | --- |
| VILPA |  |  |  |
| Up to 1 min | 30 | 38.8 | 15.7 |
| Up to 2 min | 30 | 47.2 | 25.7 |
| Over 2 min | 190 | 225.5 | 319.6 |
| VPA |  |  |  |
| Up to 1 min | 30 | 40.1 | 16.1 |
| Up to 2 min | 30 | 51.7 | 28.3 |
| Over 2 min | 240 | 362.8 | 492.5 |

2) Females; values represent seconds:

|  | Median | Mean | SD |
| --- | --- | --- | --- |
| VILPA |  |  |  |
| Up to 1 min | 20 | 26.6 | 15.0 |
| Up to 2 min | 30 | 32.8 | 23.7 |
| Over 2 min | 190 | 218.7 | 293.1 |
| VPA |  |  |  |
| Up to 1 min | 30 | 38.3 | 15.6 |
| Up to 2 min | 30 | 47.6 | 26.9 |
| Over 2 min | 220 | 291.7 | 376.7 |

**eTable 3:** Hazard ratios associated with the minimum dose and median VILPA values among non-exercisers for bouts lasting up to 1 minute (n= 22,368).

A. All MACE

| **VILPA duration (minutes/day)** | Dose | HR | Lower 95 CI | Upper 95 CI |
| --- | --- | --- | --- | --- |
| Minimum dose | 1.5 | 0.81 | 0.72 | 0.92 |
| VILPA median | 4.3 | 0.67 | 0.55 | 0.84 |
| **Length standardised frequency (1-minute equivalent bouts/day)** |  |  |  |  |
| Minimum dose | 0.9 | 0.77 | 0.67 | 0.89 |
| VILPA median | 1.7 | 0.68 | 0.56 | 0.83 |
| **Raw Frequency (bouts/day)** |  |  |  |  |
| Minimum dose | 5.6 | 0.77 | 0.65 | 0.90 |
| VILPA median | 10.1 | 0.68 | 0.54 | 0.85 |

Minimal dose (ED50 value): defined as the duration/frequency of VILPA associated with 50% of the optimal risk reduction. The VILPA duration and frequency median values were calculated in the sample excluding participants with zero VILPA. Analyses adjusted for age, light intensity, moderate intensity, VILPA bouts over 1-minute, smoking history, alcohol consumption, accelerometry-derived sleep duration, diet, education, self-reported parental history of CVD, previous incidence of cancer, and self-reported medication use (cholesterol, blood pressure, and diabetes). Raw frequency bouts were additionally adjusted for residual of VILPA bouts of 1 minutes.

B. Myocardial infarction

| **VILPA duration (minutes/day)** | Dose | HR | Lower 95 CI | Upper 95 CI |
| --- | --- | --- | --- | --- |
| Minimum dose | 1.6 | 0.81 | 0.68 | 0.97 |
| VILPA median | 4.3 | 0.67 | 0.49 | 0.92 |
| **Length standardised frequency (1-minute equivalent bouts/day)** |  |  |  |  |
| Minimum dose | 0.9 | 0.77 | 0.63 | 0.94 |
| VILPA median | 1.7 | 0.67 | 0.50 | 0.90 |
| **Raw Frequency (bouts/day)** |  |  |  |  |
| Minimum dose | 11.1 | 0.76 | 0.54 | 1.07 |
| VILPA median | 10.1 | 0.77 | 0.55 | 1.09 |

Minimal dose (ED50 value): defined as the duration/frequency of VILPA associated with 50% of the optimal risk reduction. The VILPA duration and frequency median values were calculated in the sample excluding participants with zero VILPA. Analyses adjusted for age, light intensity, moderate intensity, VILPA bouts over 1-minute, smoking history, alcohol consumption, accelerometry-derived sleep duration, diet, education, self-reported parental history of CVD, previous incidence of cancer, and self-reported medication use (cholesterol, blood pressure, and diabetes). Raw frequency bouts were additionally adjusted for residual of VILPA bouts of 1 minutes.

C. Heart failure

| **VILPA duration (minutes/day)** | Dose | HR | Lower 95 CI | Upper 95 CI |
| --- | --- | --- | --- | --- |
| Minimum dose | 1.2 | 0.71 | 0.59 | 0.85 |
| VILPA median | 4.3 | 0.45 | 0.29 | 0.66 |
| **Length standardised frequency (1-minute equivalent bouts/day)** |  |  |  |  |
| Minimum dose | 0.5 | 0.73 | 0.62 | 0.86 |
| VILPA median | 1.7 | 0.46 | 0.31 | 0.67 |
| **Raw Frequency (bouts/day)** |  |  |  |  |
| Minimum dose | 2.9 | 0.67 | 0.57 | 0.79 |
| VILPA median | 10.1 | 0.36 | 0.24 | 0.54 |

Minimal dose (ED50 value): defined as the duration/frequency of VILPA associated with 50% of the optimal risk reduction. The VILPA duration and frequency median values were calculated in the sample excluding participants with zero VILPA. Analyses adjusted for age, light intensity, moderate intensity, VILPA bouts over 1-minute, smoking history, alcohol consumption, accelerometry-derived sleep duration, diet, education, self-reported parental history of CVD, previous incidence of cancer, and self-reported medication use (cholesterol, blood pressure, and diabetes). Raw frequency bouts were additionally adjusted for residual of VILPA bouts of 1 minutes.

**eTable 4:** Sex-specific hazard ratios associated with the minimum dose and median VPA values among non-exercisers for bouts lasting up to 1 minute (females n = 13018; males n= 9350).

A. All MACE

| **Female** | | | | |
| --- | --- | --- | --- | --- |
| **VILPA duration (minutes/day)** | Dose | HR | Lower 95 CI | Upper 95 CI |
| Minimum dose | 1.6 | 0.70 | 0.58 | 0.86 |
| VILPA median | 3.4 | 0.55 | 0.41 | 0.75 |
| **Length standardised frequency (1-minute equivalent bouts/day)** |  |  |  |  |
| Minimum dose | 2.2 | 0.50 | 0.37 | 0.68 |
| VILPA median | 1.4 | 0.56 | 0.42 | 0.75 |
| **Raw Frequency (bouts/day)** |  |  |  |  |
| Minimum dose | 9.6 | 0.63 | 0.46 | 0.86 |
| VILPA median | 9.3 | 0.63 | 0.46 | 0.87 |
| **Male** | |  |  |  |
| **VILPA duration (minutes/day)** |  |  |  |  |
| Minimum dose | 2.3 | 0.89 | 0.70 | 1.12 |
| VILPA median | 5.6 | 0.84 | 0.63 | 1.12 |
| **Length standardised frequency (1-minute equivalent bouts/day)** |  |  |  |  |
| Minimum dose | 1.7 | 0.85 | 0.65 | 1.10 |
| VILPA median | 2.2 | 0.83 | 0.60 | 1.10 |
| **Raw Frequency (bouts/day)** |  |  |  |  |
| Minimum dose | 4.4 | 0.86 | 0.71 | 1.03 |
| VILPA median | 11.4 | 0.76 | 0.56 | 1.02 |

B. Myocardial infarction

| **Female** | | | | |
| --- | --- | --- | --- | --- |
| **VILPA duration (minutes/day)** | Dose | HR | Lower 95 CI | Upper 95 CI |
| Minimum dose | 1.5 | 0.67 | 0.50 | 0.91 |
| VILPA median | 3.4 | 0.49 | 0.30 | 0.80 |
| **Length standardised frequency (1-minute equivalent bouts/day)** |  |  |  |  |
| Minimum dose | 1.5 | 0.50 | 0.31 | 0.79 |
| VILPA median | 1.4 | 0.51 | 0.32 | 0.80 |
| **Raw Frequency (bouts/day)** |  |  |  |  |
| Minimum dose | 12.4 | 0.59 | 0.36 | 0.99 |
| VILPA median | 9.3 | 0.67 | 0.40 | 1.11 |
| **Male** | | | | |
| **VILPA duration (minutes/day)** |  |  |  |  |
| Minimum dose | 3.9 | 0.89 | 0.59 | 1.40 |
| VILPA median | 5.6 | 0.87 | 0.58 | 1.31 |
| **Length standardised frequency (1-minute equivalent bouts/day)** |  |  |  |  |
| Minimum dose | 1.8 | 0.85 | 0.59 | 1.24 |
| VILPA median | 2.2 | 0.84 | 0.57 | 1.24 |
| **Raw Frequency (bouts/day)** |  |  |  |  |
| Minimum dose | 19.6 | 0.87 | 0.57 | 1.33 |
| VILPA median | 11.4 | 0.92 | 0.59 | 1.43 |

C. Heart failure

| **Female** | | | | |
| --- | --- | --- | --- | --- |
| **VILPA duration (minutes/day)** | Dose | HR | Lower 95 CI | Upper 95 CI |
| Minimum dose | 1.2 | 0.60 | 0.45 | 0.81 |
| VILPA median | 3.4 | 0.33 | 0.18 | 0.59 |
| **Length standardised frequency (1-minute equivalent bouts/day)** |  |  |  |  |
| Minimum dose | 0.6 | 0.55 | 0.41 | 0.74 |
| VILPA median | 1.4 | 0.31 | 0.18 | 0.54 |
| **Raw Frequency (bouts/day)** |  |  |  |  |
| Minimum dose | 3.1 | 0.59 | 0.46 | 0.76 |
| VILPA median | 9.3 | 0.28 | 0.16 | 0.49 |
| **Male** | | | | |
| **VILPA duration (minutes/day)** |  |  |  |  |
| Minimum dose | 1.2 | 0.80 | 0.61 | 1.04 |
| VILPA median | 5.6 | 0.61 | 0.35 | 1.06 |
| **Length standardised frequency (1-minute equivalent bouts/day)** |  |  |  |  |
| Minimum dose | 0.6 | 0.84 | 0.65 | 1.08 |
| VILPA median | 2.2 | 0.68 | 0.40 | 1.16 |
| **Raw Frequency (bouts/day)** |  |  |  |  |
| Minimum dose | 3.1 | 0.74 | 0.58 | 0.95 |
| VILPA median | 11.4 | 0.49 | 0.28 | 0.87 |

Minimal dose (ED_50_ value): defined as the duration/frequency of VILPA associated with 50% of the optimal risk reduction. The VILPA duration and frequency median values were calculated in the sample excluding participants with zero VILPA. Analyses adjusted for age, light intensity, moderate intensity, VILPA bouts over 1-minute, smoking history, alcohol consumption, accelerometry-derived sleep duration, diet, education, self-reported parental history of CVD, previous incidence of cancer, and self-reported medication use (cholesterol, blood pressure, and diabetes). Raw frequency bouts were additionally adjusted for residual of VILPA bouts of 1 minutes.

**eTable 5**: Hazard ratios associated with the minimum dose and median VPA values among exercisers for bouts lasting up to 1 minute.

A. All MACE

| **VILPA duration** | Dose | HR | Lower 95 CI | Upper 95 CI |
| --- | --- | --- | --- | --- |
| **VILPA duration (minutes/day)** | 3.7 | 0.79 | 0.73 | 0.86 |
| Minimum dose | 6.1 | 0.73 | 0.66 | 0.81 |
| VILPA median |  |  |  |  |
| **Length standardised frequency (1-minute equivalent bouts/day)** | 1.4 | 0.80 | 0.74 | 0.86 |
| Minimum dose | 2.4 | 0.73 | 0.64 | 0.81 |
| VILPA median |  |  |  |  |
| **Raw Frequency (bouts/day)** |  |  |  |  |
| Minimum dose | 6.5 | 0.77 | 0.72 | 0.83 |
| VILPA median | 12.7 | 0.65 | 0.58 | 0.73 |

Minimal dose (ED50 value): defined as the duration/frequency of VILPA associated with 50% of the optimal risk reduction. The VILPA duration and frequency median values were calculated in the sample excluding participants with zero VILPA. Analyses adjusted for age, light intensity, moderate intensity, VILPA bouts over 1-minute, smoking history, alcohol consumption, accelerometry-derived sleep duration, diet, education, self-reported parental history of CVD, previous incidence of cancer, and self-reported medication use (cholesterol, blood pressure, and diabetes). Raw frequency bouts were additionally adjusted for residual of VILPA bouts of 1 minutes.

B. Myocardial infarction

| **VILPA duration (minutes/day)** | Dose | HR | Lower 95 CI | Upper 95 CI |
| --- | --- | --- | --- | --- |
| Minimum dose | 2.2 | 0.85 | 0.79 | 0.91 |
| VILPA median | 6.1 | 0.70 | 0.60 | 0.82 |
| **Length standardised frequency (1-minute equivalent bouts/day)** |  |  |  |  |
| Minimum dose | 0.9 | 0.85 | 0.79 | 0.91 |
| VILPA median | 2.4 | 0.70 | 0.60 | 0.82 |
| **Raw Frequency (bouts/day)** |  |  |  |  |
| Minimum dose | 5.1 | 0.80 | 0.75 | 0.87 |
| VILPA median | 12.9 | 0.61 | 0.52 | 0.73 |

Minimal dose (ED50 value): defined as the duration/frequency of VILPA associated with 50% of the optimal risk reduction. The VILPA duration and frequency median values were calculated in the sample excluding participants with zero VILPA. Analyses adjusted for age, light intensity, moderate intensity, VILPA bouts over 1-minute, smoking history, alcohol consumption, accelerometry-derived sleep duration, diet, education, self-reported parental history of CVD, previous incidence of cancer, and self-reported medication use (cholesterol, blood pressure, and diabetes). Raw frequency bouts were additionally adjusted for residual of VILPA bouts of 1 minutes.

C. Heart failure

| **VILPA duration (minutes/day)** | Dose | HR | Lower 95 CI | Upper 95 CI |
| --- | --- | --- | --- | --- |
| Minimum dose | 5.1 | 0.64 | 0.53 | 0.77 |
| VILPA median | 6.1 | 0.61 | 0.50 | 0.75 |
| **Length standardised frequency (1-minute equivalent bouts/day)** |  |  |  |  |
| Minimum dose | 1.9 | 0.64 | 0.54 | 0.77 |
| VILPA median | 2.4 | 0.60 | 0.50 | 0.73 |
| **Raw Frequency (bouts/day)** |  |  |  |  |
| Minimum dose | 7.9 | 0.62 | 0.53 | 0.74 |
| VILPA median | 12.9 | 0.52 | 0.41 | 0.63 |

Minimal dose (ED50 value): defined as the duration/frequency of VILPA associated with 50% of the optimal risk reduction. The VILPA duration and frequency median values were calculated in the sample excluding participants with zero VILPA. Analyses adjusted for age, light intensity, moderate intensity, VILPA bouts over 1-minute, smoking history, alcohol consumption, accelerometry-derived sleep duration, diet, education, self-reported parental history of CVD, previous incidence of cancer, and self-reported medication use (cholesterol, blood pressure, and diabetes). Raw frequency bouts were additionally adjusted for residual of VILPA bouts of 1 minutes.

**eTable 6:** Sex-specific hazard ratios associated with the minimum dose and median VPA values among exercisers for bouts lasting up to 1 minute.

A. All MACE

| **Female** | | | | |
| --- | --- | --- | --- | --- |
| **VILPA duration (minutes/day)** | Dose | HR | Lower 95 CI | Upper 95 CI |
| Minimum dose | 2.7 | 0.84 | 0.73 | 0.97 |
| VILPA median | 5.1 | 0.77 | 0.64 | 0.94 |
| **Length standardised frequency (1-minute equivalent bouts/day)** |  |  |  |  |
| Minimum dose | 1.3 | 0.82 | 0.70 | 0.97 |
| VILPA median | 5.1 | 0.74 | 0.61 | 0.90 |
| **Raw Frequency (bouts/day)** |  |  |  |  |
| Minimum dose | 6.3 | 0.80 | 0.68 | 0.96 |
| VILPA median | 5.1 | 0.84 | 0.72 | 0.96 |
| **Male** | |  |  |  |
| **VILPA duration (minutes/day)** |  |  |  |  |
| Minimum dose | 3.8 | 0.79 | 0.71 | 0.88 |
| VILPA median | 8.1 | 0.68 | 0.57 | 0.80 |
| **Length standardised frequency (1-minute equivalent bouts/day)** |  |  |  |  |
| Minimum dose | 1.5 | 0.78 | 0.70 | 0.88 |
| VILPA median | 8.1 | 0.63 | 0.53 | 0.75 |
| **Raw Frequency (bouts/day)** |  |  |  |  |
| Minimum dose | 6.1 | 0.74 | 0.66 | 0.83 |
| VILPA median | 8.1 | 0.67 | 0.58 | 0.79 |

B. Myocardial infarction

| **Female** | | | | |
| --- | --- | --- | --- | --- |
| **VILPA duration (minutes/day)** | Dose | HR | Lower 95 CI | Upper 95 CI |
| Minimum dose | 3.5 | 0.69 | 0.53 | 0.89 |
| VILPA median | 5.1 | 0.62 | 0.45 | 0.84 |
| **Length standardised frequency (1-minute equivalent bouts/day)** |  |  |  |  |
| Minimum dose | 1.4 | 0.69 | 0.53 | 0.89 |
| VILPA median | 2.1 | 0.61 | 0.44 | 0.85 |
| **Raw Frequency (bouts/day)** |  |  |  |  |
| Minimum dose | 5.8 | 0.65 | 0.51 | 0.82 |
| VILPA median | 11.9 | 0.45 | 0.31 | 0.66 |
| **Male** | | | | |
| **VILPA duration (minutes/day)** |  |  |  |  |
| Minimum dose | 2.2 | 0.83 | 0.73 | 0.95 |
| VILPA median | 8.3 | 0.66 | 0.50 | 0.87 |
| **Length standardised frequency (1-minute equivalent bouts/day)** |  |  |  |  |
| Minimum dose | 0.8 | 0.83 | 0.74 | 0.95 |
| VILPA median | 3.2 | 0.66 | 0.50 | 0.87 |
| **Raw Frequency (bouts/day)** |  |  |  |  |
| Minimum dose | 4.1 | 0.74 | 0.65 | 0.85 |
| VILPA median | 15 | 0.49 | 0.35 | 0.68 |

C. Heart failure

| **Female** | | | | |
| --- | --- | --- | --- | --- |
| **VILPA duration (minutes/day)** | Dose | HR | Lower 95 CI | Upper 95 CI |
| Minimum dose | 4.5 | 0.71 | 0.49 | 1.04 |
| VILPA median | 5.1 | 0.70 | 0.47 | 1.03 |
| **Length standardised frequency (1-minute equivalent bouts/day)** |  |  |  |  |
| Minimum dose | 1.8 | 0.67 | 0.47 | 0.96 |
| VILPA median | 2.1 | 0.65 | 0.44 | 0.95 |
| **Raw Frequency (bouts/day)** |  |  |  |  |
| Minimum dose | 10.6 | 0.73 | 0.47 | 1.13 |
| VILPA median | 5.1 | 0.84 | 0.65 | 1.10 |
| **Male** | | | | |
| **VILPA duration (minutes/day)** |  |  |  |  |
| Minimum dose | 5.5 | 0.65 | 0.47 | 0.89 |
| VILPA median | 8.3 | 0.58 | 0.40 | 0.83 |
| **Length standardised frequency (1-minute equivalent bouts/day)** |  |  |  |  |
| Minimum dose | 2.5 | 0.65 | 0.45 | 0.94 |
| VILPA median | 3.2 | 0.62 | 0.43 | 0.90 |
| **Raw Frequency (bouts/day)** |  |  |  |  |
| Minimum dose | 8.6 | 0.65 | 0.47 | 0.90 |
| VILPA median | 8.3 | 0.66 | 0.48 | 0.91 |

Minimal dose (ED_50_ value): defined as the duration/frequency of VILPA associated with 50% of the optimal risk reduction. The VILPA duration and frequency median values were calculated in the sample excluding participants with zero VILPA. Analyses adjusted for age, light intensity, moderate intensity, VILPA bouts over 1-minute, smoking history, alcohol consumption, accelerometry-derived sleep duration, diet, education, self-reported parental history of CVD, previous incidence of cancer, and self-reported medication use (cholesterol, blood pressure, and diabetes). Raw frequency bouts were additionally adjusted for residual of VILPA bouts of 1 minutes.

**eTable 7:** Sex-specific e-values for minimum dose and median VILPA values for MACE and its subtypes among non-exercisers for bouts lasting up to 1 minute.

D. All MACE

| **Female** | |
| --- | --- |
| **VILPA duration (minutes/day)** | E-value |
| Minimum dose | 2.21 (1.60) |
| VILPA median | 3.04 (2.00) |
| **Length standardised frequency (1-minute equivalent bouts/day)** |  |
| Minimum dose | 3.41 (2.30) |
| VILPA median | 2.97 (2.00) |
| **Raw Frequency (bouts/day)** |  |
| Minimum dose | 2.55 (1.60) |
| VILPA median | 2.55 (1.56) |
| **Male** | |
| **VILPA duration (minutes/day)** |  |
| Minimum dose | 1.49 (1.00) |
| VILPA median | 1.67 (1.00) |
| **Length standardised frequency (1-minute equivalent bouts/day)** |  |
| Minimum dose | 1.63 (1.00) |
| VILPA median | 1.70 (1.00) |
| **Raw Frequency (bouts/day)** |  |
| Minimum dose | 1.60 (1.00) |
| VILPA median | 1.96 (1.00) |

E. Myocardial infarction

| **Female** | |
| --- | --- |
| **VILPA duration (minutes/day)** | E-value |
| Minimum dose | 2.34 (1.42) |
| VILPA median | 3.49 (1.80) |
| **Length standardised frequency (1-minute equivalent bouts/day)** |  |
| Minimum dose | 3.41 (1.85) |
| VILPA median | 3.33 (1.81) |
| **Raw Frequency (bouts/day)** |  |
| Minimum dose | 2.78 (0.99) |
| VILPA median | 2.34 (1.00) |
| **Male** | |
| **VILPA duration (minutes/day)** |  |
| Minimum dose | 1.50 (1.00) |
| VILPA median | 1.56 (1.00) |
| **Length standardised frequency (1-minute equivalent bouts/day)** |  |
| Minimum dose | 1.63 (1.00) |
| VILPA median | 1.67 (1.00) |
| **Raw Frequency (bouts/day)** |  |
| Minimum dose | 1.56 (1.00) |
| VILPA median | 1.39 (1.00) |

F. Heart failure

| **Female** | |
| --- | --- |
| **VILPA duration (minutes/day)** | E-value |
| Minimum dose | 2.72 (1.77) |
| VILPA median | 5.51 (2.78) |
| **Length standardised frequency (1-minute equivalent bouts/day)** |  |
| Minimum dose | 3.04 (2.04) |
| VILPA median | 5.91 (3.12) |
| **Raw Frequency (bouts/day)** |  |
| Minimum dose | 2.78 (1.96) |
| VILPA median | 6.60 (3.50) |
| **Male** | |
| **VILPA duration (minutes/day)** |  |
| Minimum dose | 1.81 (1.00) |
| VILPA median | 2.66 (1.00) |
| **Length standardised frequency (1-minute equivalent bouts/day)** |  |
| Minimum dose | 1.67 (1.00) |
| VILPA median | 2.30 (1.00) |
| **Raw Frequency (bouts/day)** |  |
| Minimum dose | 2.04 (1.29) |
| VILPA median | 3.49 (1.56) |

**eTable 8:** Sex-specific e-values for minimum dose and median VPA values for MACE and its subtypes among exercisers for bouts lasting up to 1 minute.

A. All MACE

| **Female** | |
| --- | --- |
| **VILPA duration (minutes/day)** | E-value |
| Minimum dose | 1.67 (1.20) |
| VILPA median | 1.92 (1.32) |
| **Length standardised frequency (1-minute equivalent bouts/day)** |  |
| Minimum dose | 1.73 (1.21) |
| VILPA median | 2.04 (1.46) |
| **Raw Frequency (bouts/day)** |  |
| Minimum dose | 1.81 (1.25) |
| VILPA median | 1.67 (1.25) |
| **Male** | |
| **VILPA duration (minutes/day)** |  |
| Minimum dose | 1.85 (1.53) |
| VILPA median | 2.30 (1.81) |
| **Length standardised frequency (1-minute equivalent bouts/day)** |  |
| Minimum dose | 1.88 (1.53) |
| VILPA median | 2.34 (1.78) |
| **Raw Frequency (bouts/day)** |  |
| Minimum dose | 2.04 (1.70) |
| VILPA median | 2.35 (1.85) |

B. Myocardial infarction

| **Female** | |
| --- | --- |
| **VILPA duration (minutes/day)** | E-value |
| Minimum dose | 2.60 (1.51) |
| VILPA median | 2.61 (1.67) |
| **Length standardised frequency (1-minute equivalent bouts/day)** |  |
| Minimum dose | 2.26 (1.50) |
| VILPA median | 2.66 (1.63) |
| **Raw Frequency (bouts/day)** |  |
| Minimum dose | 2.44 (1.74) |
| VILPA median | 3.87 (2.40) |
| **Male** | |
| **VILPA duration (minutes/day)** |  |
| Minimum dose | 1.70 (1.29) |
| VILPA median | 2.40 (1.56) |
| **Length standardised frequency (1-minute equivalent bouts/day)** |  |
| Minimum dose | 1.70 (1.29) |
| VILPA median | 2.40 (1.56) |
| **Raw Frequency (bouts/day)** |  |
| Minimum dose | 2.04 (1.63) |
| VILPA median | 3.49 (2.30) |

C. Heart failure

| **Female** | |
| --- | --- |
| **VILPA duration (minutes/day)** | E-value |
| Minimum dose | 2.17 (1.00) |
| VILPA median | 2.21 (1.00) |
| **Length standardised frequency (1-minute equivalent bouts/day)** |  |
| Minimum dose | 2.35 (1.25) |
| VILPA median | 2.44 (1.29) |
| **Raw Frequency (bouts/day)** |  |
| Minimum dose | 2.08 (1.00) |
| VILPA median | 1.67 (1.00) |
| **Male** | |
| **VILPA duration (minutes/day)** |  |
| Minimum dose | 2.45 (1.50) |
| VILPA median | 2.84 (1.70) |
| **Length standardised frequency (1-minute equivalent bouts/day)** |  |
| Minimum dose | 2.45 (1.32) |
| VILPA median | 2.61 (1.46) |
| **Raw Frequency (bouts/day)** |  |
| Minimum dose | 2.45 (1.46) |
| VILPA median | 2.40 (1.43) |

**eTable 9:** Covariate definitions

| **Variable** | **Definition** | **UK Biobank field ID (if applicable)** |
| --- | --- | --- |
| Age | Continuous | 34, 52, accelerometer date-timestamp |
| Sex | Female/Male | 31 |
| Ethnicity | White/Others () |  |
| Light intensity physical activity | Standing utilitarian movements, slow walking (<3 METs) | Derived from accelerometer data (see Online Methods) |
| Moderate intensity physical activity | Brisk walking, energetic activities (≥3 to <6 METs) | Derived from accelerometer data (see see Online Methods) |
| Frequency/duration of bouts above the specified bout length (> 1 or > 2 minutes) in each frequency/duration analysis | Number of bouts above the specified bout in the analysis. Eg, for the analysis of bouts lasting up to 2 minutes in duration, this variable contained bouts/duration that were more than 2 minutes in duration | Derived from accelerometer data (see Methods) |
| Smoking status | Never, past, current | 20116 |
| Alcohol consumption | Never, ex-drinker, within guidelines, above guidelines | 20117, 1558 |
| Sleep duration | Hours spent sleeping | Derived from accelerometer data (see Methods) |
| Diet | Fruits and vegetables servings/day, categorised as low (<5 servings/day), moderate (5 to 8 servings/day) and high (>8 servings/day) | 1309, 1319, 1289, 1299 |
| Prevalent cancer | Identified by self-report and cancer registry | 20001, 100092 |
| Education | College/University; A/AS level; O levels; CSE; NVQ/HND/HNC; other | 6138 |
| Parental history of CVD | Self-reported mother or father diagnosed with heart disease or stroke | 20107, 20110 |
| Use of medication (cholesterol, blood pressure and diabetes) | Yes/No | 6177, 6153 |

**eTable 10:** Interaction test between age and daily VILPA duration bouts lasting up to 1 minute (minutes/day) for MACE and its subtypes.

| Outcome | Age*VILPA, p-values | | |
| --- | --- | --- | --- |
|  | Female | Male | Total* |
| MACE | 0.437 | 0.941 | 0.487 |
| Myocardial infarction | 0.811 | 0.713 | 0.480 |
| Heart failure | 0.172 | 0.480 | 0.564 |
| Stroke | 0.962 | 0.771 | 0.756 |

The models were adjusted for sex (in the total sample only)*, light intensity, moderate intensity, VILPA bouts over 1-minute, smoking history, alcohol consumption, accelerometry-derived sleep duration, diet, education, ethnicity, self-reported parental history of CVD, previous incidence of cancer, and self-reported medication use (cholesterol, blood pressure, and diabetes).

**eTable 11:** Interaction test between sex and daily VILPA duration bouts lasting up to 1 minute (minutes/day) for MACE and its subtypes.

| Outcome | Sex*VILPA, p-values |
| --- | --- |
| MACE | 0.006 |
| Myocardial infarction | 0.031 |
| Heart failure | 0.008 |
| Stroke | 0.742 |

The models were adjusted for age, light intensity, moderate intensity, VILPA bouts over 1-minute, smoking history, alcohol consumption, accelerometry-derived sleep duration, diet, education, ethnicity, self-reported parental history of CVD, previous incidence of cancer, and self-reported medication use (cholesterol, blood pressure, and diabetes).
**eTable 12:** STROBE Statement

|  | Item No | Recommendation | | Page  No |
| --- | --- | --- | --- | --- |
| **Title and abstract** | 1 | (*a*) Indicate the study’s design with a commonly used term in the title or the abstract | |  |
|  |  | (*b*) Provide in the abstract an informative and balanced summary of what was done and what was found | |  |
| Introduction | | | | |
| Background/rationale | 2 | Explain the scientific background and rationale for the investigation being reported | |  |
| Objectives | 3 | State specific objectives, including any prespecified hypotheses | |  |
| Methods | | | | |
| Study design | 4 | Present key elements of study design early in the paper | |  |
| Setting | 5 | Describe the setting, locations, and relevant dates, including periods of recruitment, exposure, follow-up, and data collection | |  |
| Participants | 6 | (*a*) *Cohort study*—Give the eligibility criteria, and the sources and methods of selection of participants. Describe methods of follow-up  *Case-control study*—Give the eligibility criteria, and the sources and methods of case ascertainment and control selection. Give the rationale for the choice of cases and controls  *Cross-sectional study*—Give the eligibility criteria, and the sources and methods of selection of participants | |  |
|  |  | (*b*) *Cohort study*—For matched studies, give matching criteria and number of exposed and unexposed  *Case-control study*—For matched studies, give matching criteria and the number of controls per case | |  |
| Variables | 7 | Clearly define all outcomes, exposures, predictors, potential confounders, and effect modifiers. Give diagnostic criteria, if applicable | |  |
| Data sources/ measurement | 8* | For each variable of interest, give sources of data and details of methods of assessment (measurement). Describe comparability of assessment methods if there is more than one group | |  |
| Bias | 9 | Describe any efforts to address potential sources of bias | |  |
| Study size | 10 | Explain how the study size was arrived at | |  |
| Quantitative variables | 11 | Explain how quantitative variables were handled in the analyses. If applicable, describe which groupings were chosen and why | |  |
| Statistical methods | 12 | (*a*) Describe all statistical methods, including those used to control for confounding | |  |
|  |  | (*b*) Describe any methods used to examine subgroups and interactions | |  |
|  |  | (*c*) Explain how missing data were addressed | |  |
|  |  | (*d*) *Cohort study*—If applicable, explain how loss to follow-up was addressed  *Case-control study*—If applicable, explain how matching of cases and controls was addressed  *Cross-sectional study*—If applicable, describe analytical methods taking account of sampling strategy | |  |
|  |  | (*e*) Describe any sensitivity analyses | |  |
| **Results** |  | |  | |
| Participants | 13* | | (a) Report numbers of individuals at each stage of study—eg numbers potentially eligible, examined for eligibility, confirmed eligible, included in the study, completing follow-up, and analysed | |
|  |  | | (b) Give reasons for non-participation at each stage | |
|  |  | | (c) Consider use of a flow diagram | |
| Descriptive data | 14* | | (a) Give characteristics of study participants (eg demographic, clinical, social) and information on exposures and potential confounders | |
|  |  | | (b) Indicate number of participants with missing data for each variable of interest | |
|  |  | | (c) *Cohort study*—Summarise follow-up time (eg, average and total amount) | |
| Outcome data | 15* | | *Cohort study*—Report numbers of outcome events or summary measures over time | |
|  |  | | *Case-control study—*Report numbers in each exposure category, or summary measures of exposure | |
|  |  | | *Cross-sectional study—*Report numbers of outcome events or summary measures | |
| Main results | 16 | | (*a*) Give unadjusted estimates and, if applicable, confounder-adjusted estimates and their precision (eg, 95% confidence interval). Make clear which confounders were adjusted for and why they were included | |
|  |  | | (*b*) Report category boundaries when continuous variables were categorized | |
|  |  | | (*c*) If relevant, consider translating estimates of relative risk into absolute risk for a meaningful time period | |
| Other analyses | 17 | | Report other analyses done—eg analyses of subgroups and interactions, and sensitivity analyses | |
| **Discussion** |  | |  | |
| Key results | 18 | | Summarise key results with reference to study objectives | |
| Limitations | 19 | | Discuss limitations of the study, taking into account sources of potential bias or imprecision. Discuss both direction and magnitude of any potential bias | |
| Interpretation | 20 | | Give a cautious overall interpretation of results considering objectives, limitations, multiplicity of analyses, results from similar studies, and other relevant evidence | |
| Generalisability | 21 | | Discuss the generalisability (external validity) of the study results | |
| **Other information** |  | |  | |
| Funding | 22 | | Give the source of funding and the role of the funders for the present study and, if applicable, for the original study on which the present article is based | |

**eTable 14:** Estimated maximal oxygen consumption, and average oxygen consumption and relative intensity of VILPA bouts (up to 1 minute) among 1,588 males non-exercisers UK Biobank participants with valid accelerometry and ergometer data (**eText 3).**

| **VO_2_ max** |  |
| --- | --- |
| *Females* | 25.56 (6.82) ml/kg/min  7.30 (1.95) METs |
| *Males* | 32.32 (8.78) ml/kg/min  9.24 (2.51) METs |
| **Average** **VO_2_ during VILPA bouts** |  |
| *Females* | 21.05 (3.56) ml/kg/min  6.04 (1.02) METs |
| *Males* | 21.74 (5.32) ml/kg/min  6.21 (1.52) METs |
| **Average relative intensity** **(%****VO2max) during VILPA bouts** |  |
| *Females* | 83.2 (18.2) % |
| *Males* | 70.5 (22.1) % |

**eFigure 1:** Flow diagram of non-exerciser participants

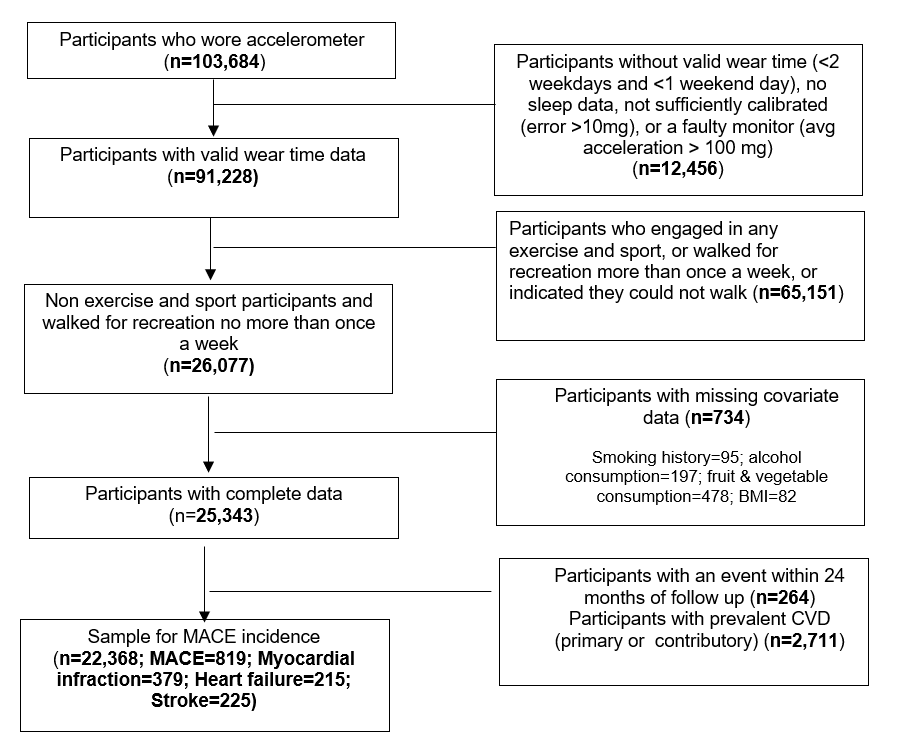

**eFigure 2:** Flow diagram of exercisers participants

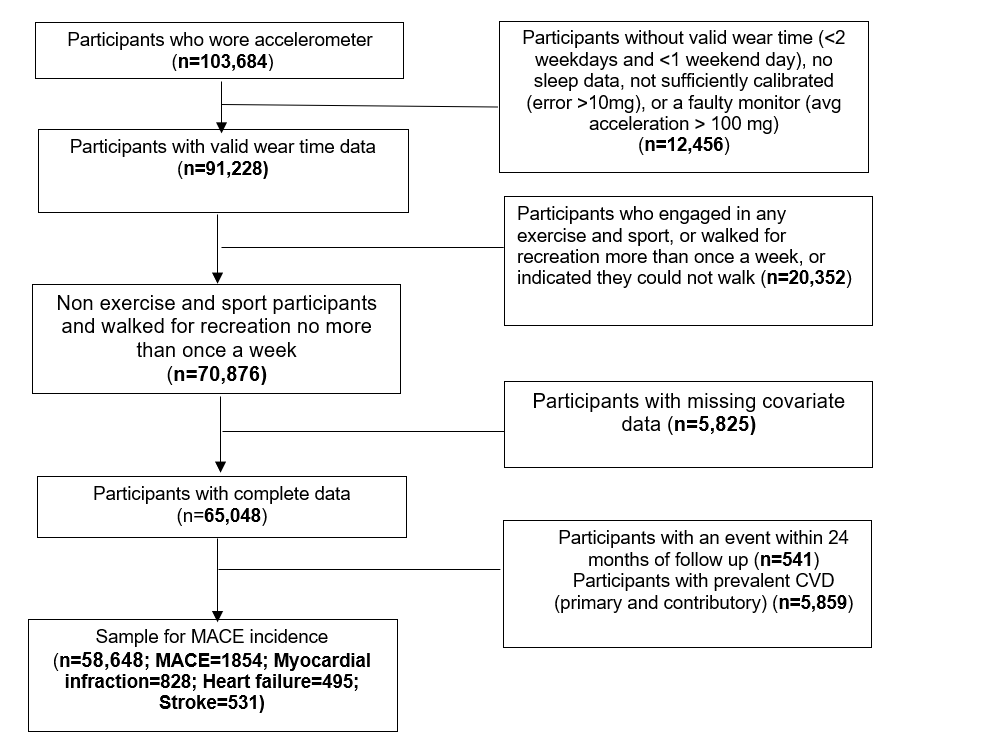

**eFigure 3:** Unadjusted absolute risk estimates for MACE and its subtypes by daily VILPA duration, bouts lasting up to 1 minute (minutes/day).

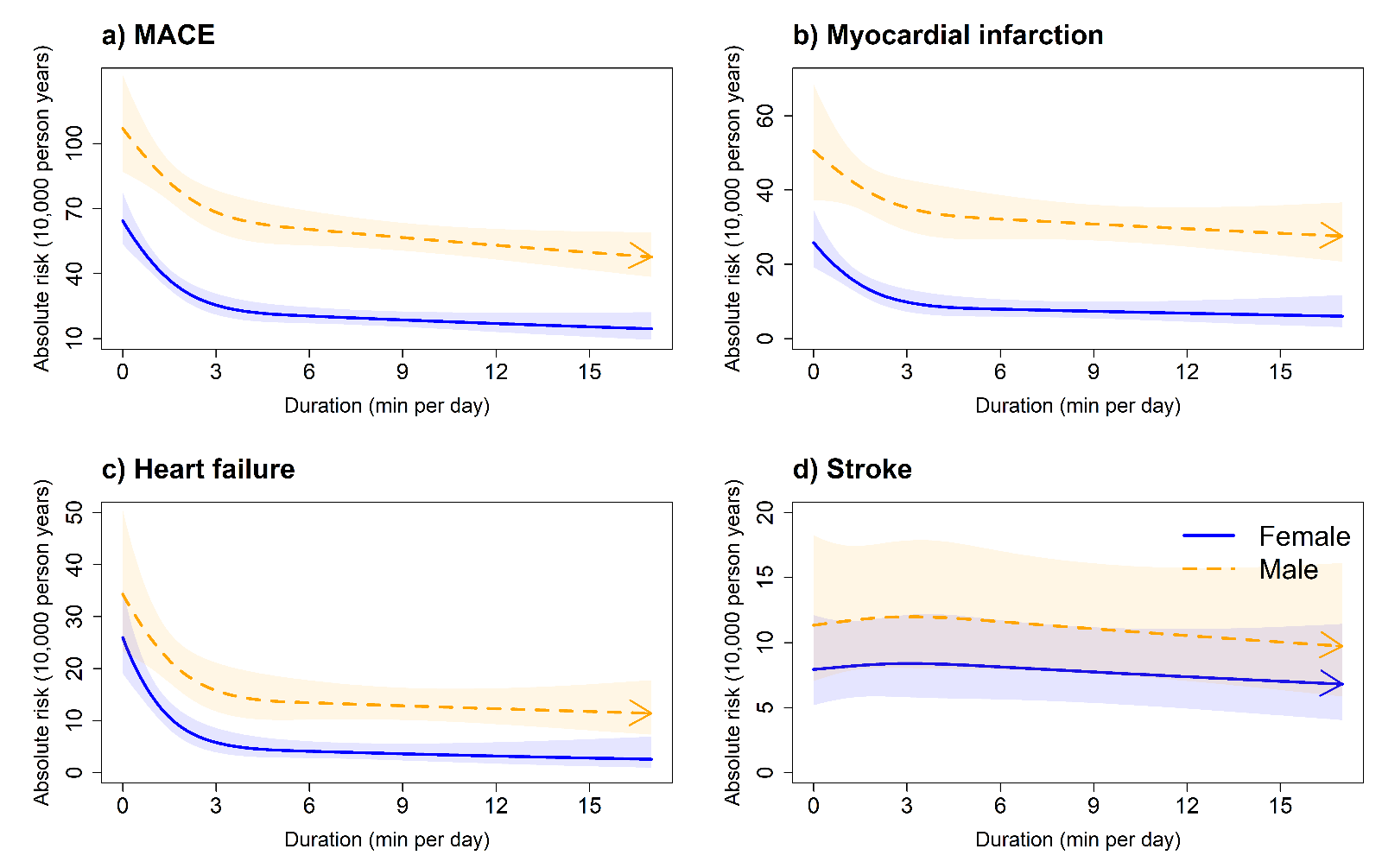

***Legend:*** The models were not adjusted for any covariates. **Panel A:** MACE: n = 22,368; events: all MACE = 819 (female/male = 331/488), **Panel B:** myocardial infarction: n = 21,928; events = 379 (female/male = 129/250). **Panel C:** heart failure: n = 21,764; events = 215 (female/male = 96/119). **Panel D:** stroke: n = 21,774; events = 225 (female/male = 106/119).
**eFigure 4:** Sex-specific adjusted absolute risk estimates of daily VILPA duration for MACE and its subtypes, bouts lasting up to 1 minute (minutes/day).

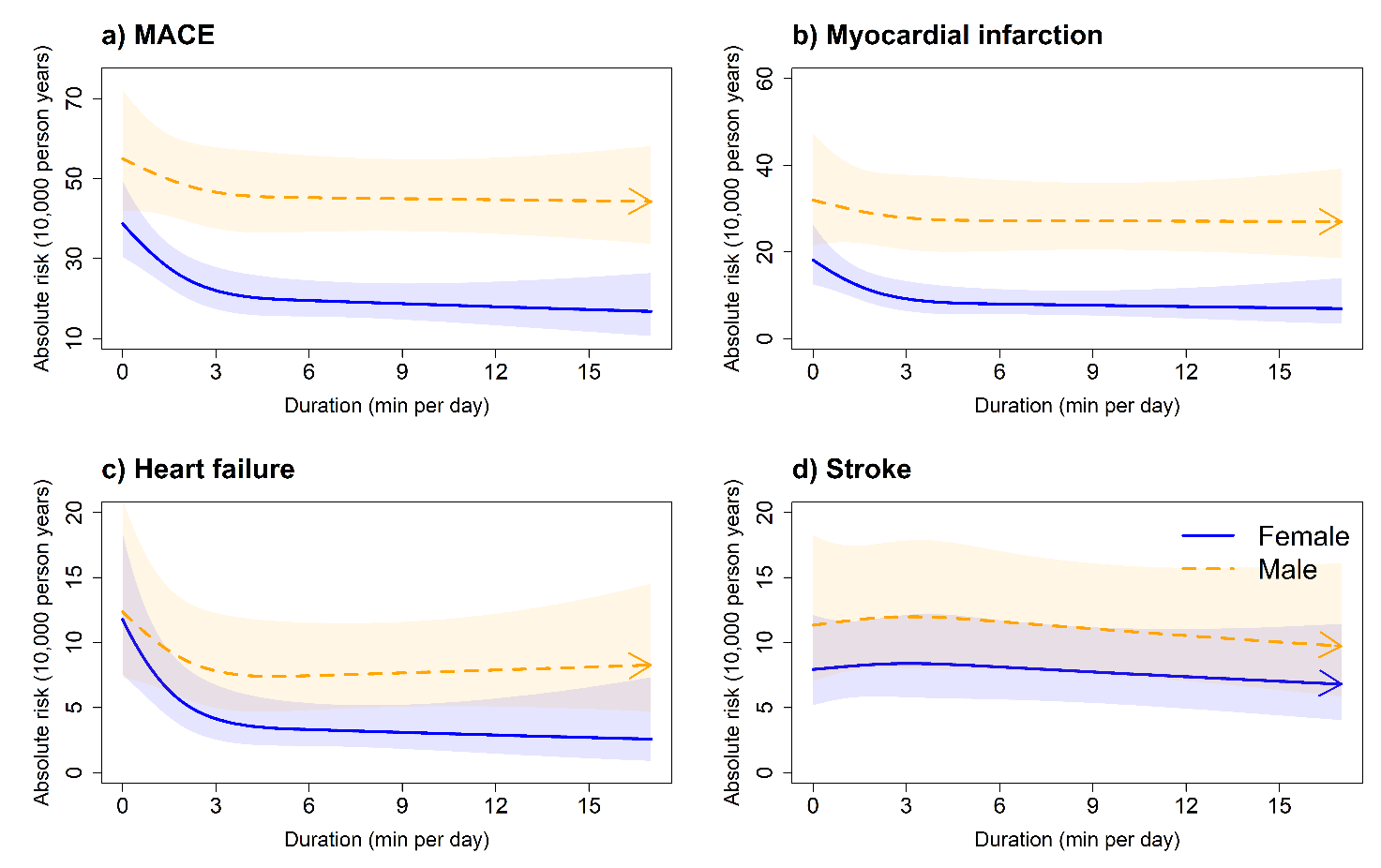

***Legend:*** Adjusted for age, light intensity, moderate intensity, VILPA bouts over 1-minute, smoking history, alcohol consumption, accelerometer estimated sleep duration, diet, education, ethnicity, self-reported parental history of CVD, previous incidence of cancer, and self-reported medication use (cholesterol, blood pressure, and diabetes). **Panel A:** MACE: n = 22,368; events: all MACE = 819 (female/male = 331/488), **Panel B:** myocardial infarction: n = 21,928; events = 379 (female/male = 129/250). **Panel C:** heart failure: n = 21,764; events = 215 (female/male = 96/119). **Panel D:** stroke: n = 21,774; events = 225 (female/male = 106/119).

**eFigure 5:** Adjusted dose response curves of length-standardised frequency for MACE and its sub-types, VILPA bouts equivalent to 1 minute (bouts/day).

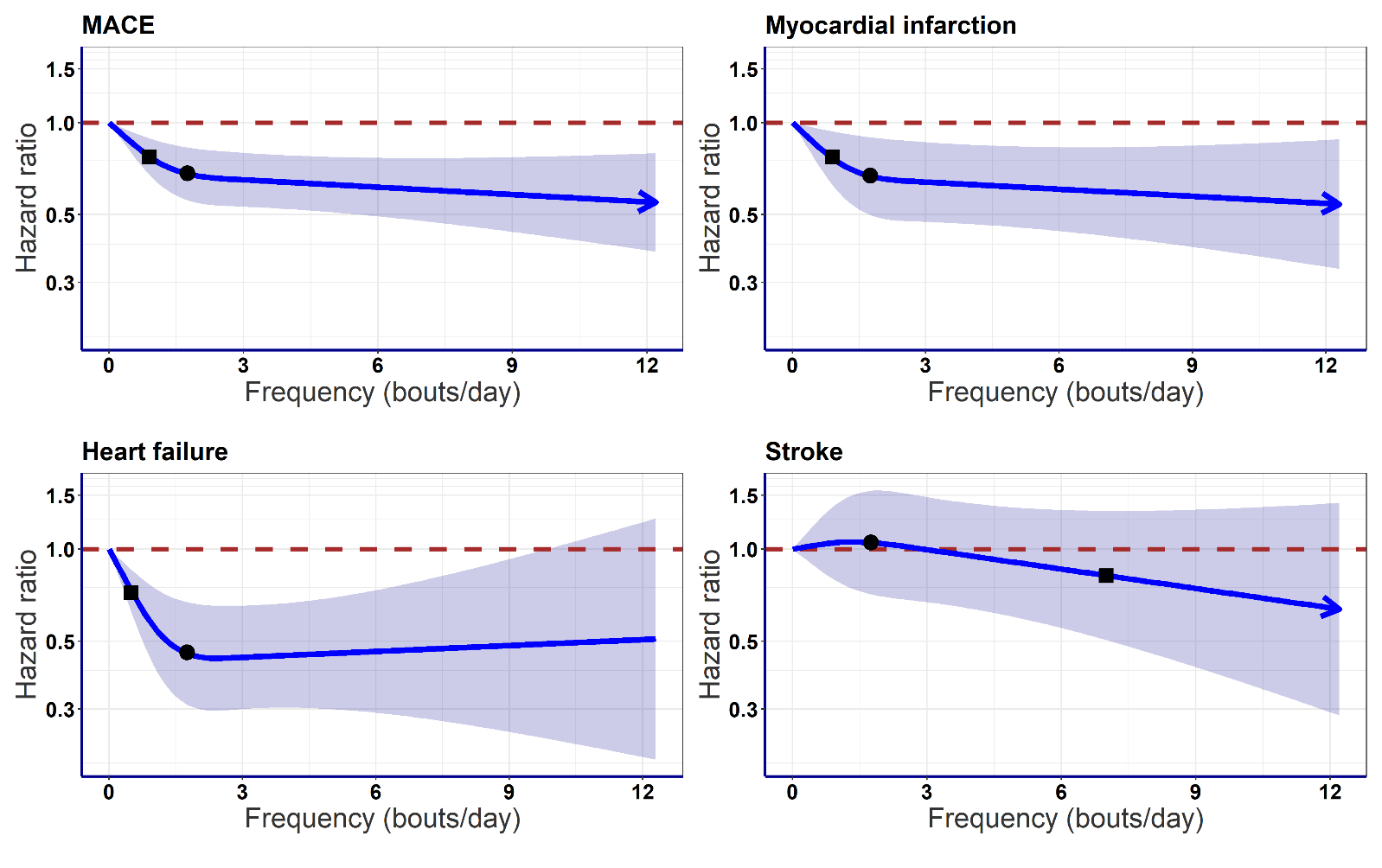

***Legend:*** Adjusted for sex, age, light intensity, moderate intensity, VILPA bouts over 1-minute, smoking history, alcohol consumption, accelerometry-derived sleep duration, diet, education, ethnicity, self-reported parental history of CVD, previous incidence of cancer, and self-reported medication use (cholesterol, blood pressure, and diabetes). **Panel A:** all MACE: n = 22,368; events: 819, **Panel B:** myocardial infarction: n = 21,928; events = 379. **Panel C:** heart failure: n = 21,764; events = 215. **Panel D:** stroke: n = 21,774; events = 225. Diamond, minimal dose, as indicated by the ED_50_ statistic which estimates the daily duration of VILPA associated with 50% of optimal risk reduction. Circle, HR associated with the median VILPA value (see **eTable 3** for the list of values).

**eFigure 6:** Adjusted dose response curves of raw VILPA frequency for MACE and its sub-types, VILPA bouts lasting up to 1 minutes (bouts/day).

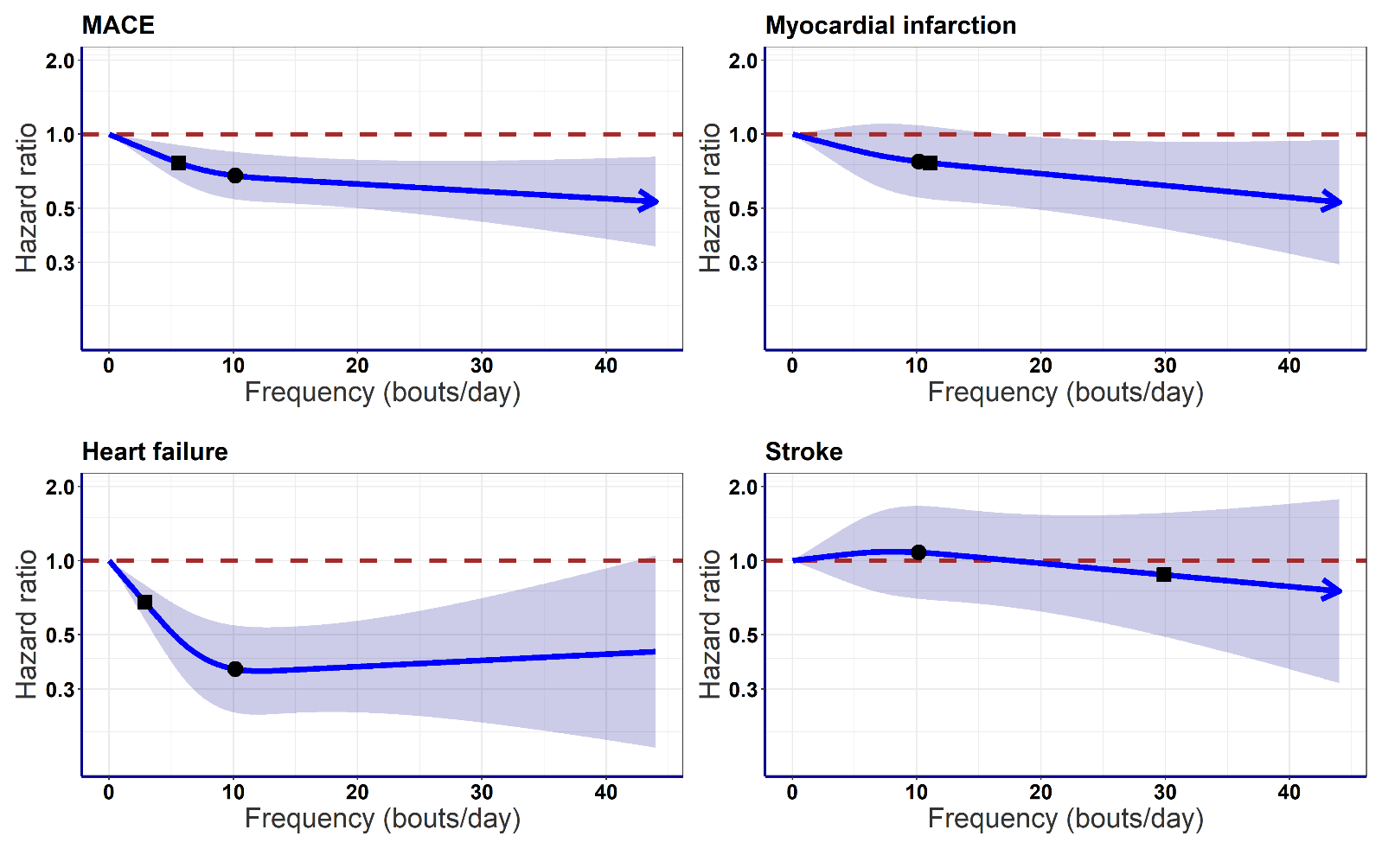

***Legend:*** Adjusted for sex, age, light intensity, moderate intensity, VILPA bouts over 1-minute, smoking history, alcohol consumption, accelerometry-derived sleep duration, diet, education, ethnicity, self-reported parental history of CVD, previous incidence of cancer, self-reported medication use (cholesterol, blood pressure, and diabetes) and residual of VILPA duration of 1-minute bouts. **Panel A:** all MACE: n = 22,368; events: 819, **Panel B:** myocardial infarction: n = 21,928; events = 379. **Panel C:** heart failure: n = 21,764; events = 215. **Panel D:** stroke: n = 21,774; events = 225. Diamond, minimal dose, as indicated by the ED_50_ statistic which estimates the daily duration of VILPA associated with 50% of optimal risk reduction. Circle, HR associated with the median VILPA value (see eTable 3 for the list of values).

**eFigure 7:** Adjusted sex-specific dose response curves of raw VILPA frequency for MACE and its sub-types, VILPA bouts lasting up to 1 minutes (bouts/day).

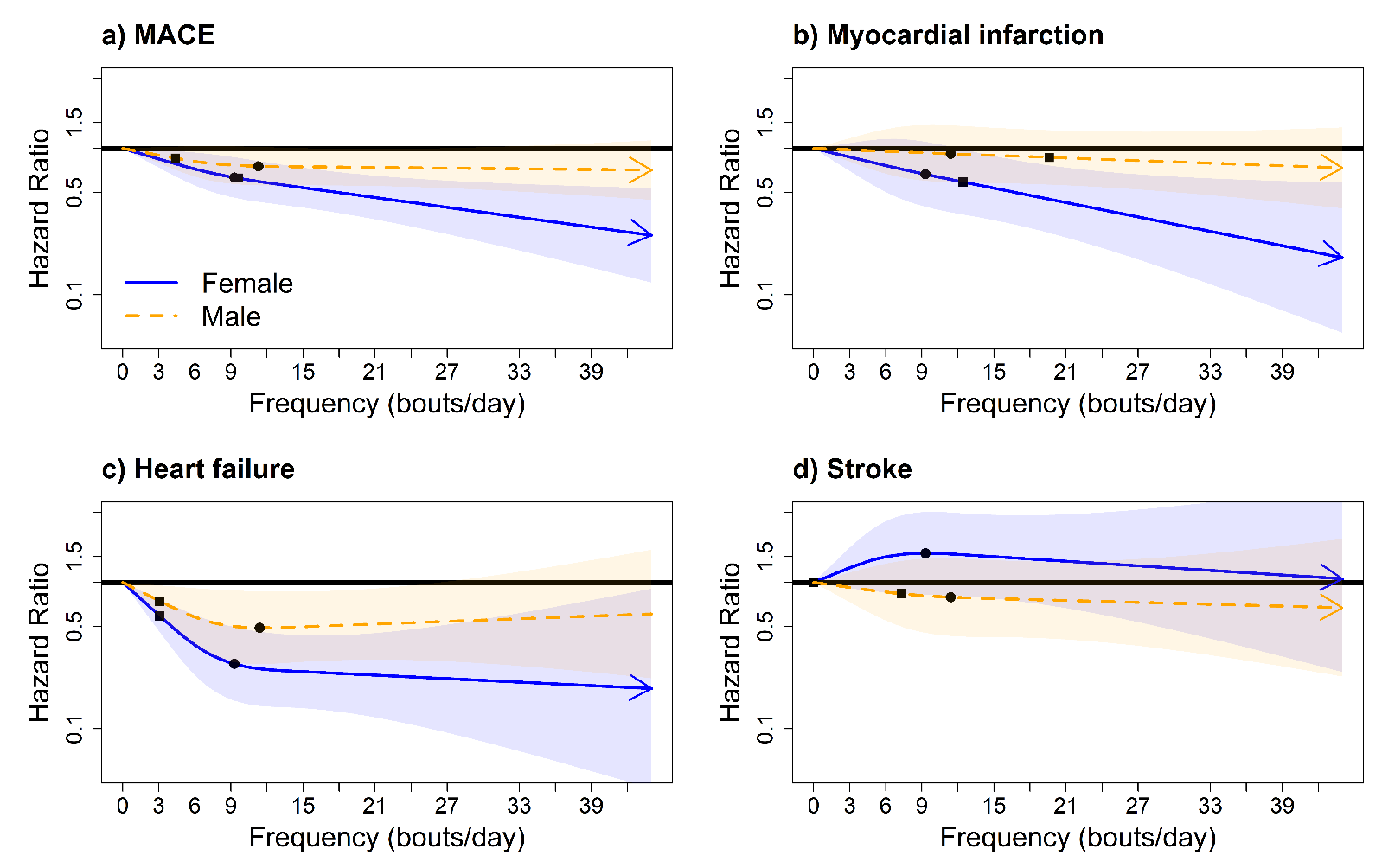

***Legend:*** Adjusted for age, light intensity, moderate intensity, VILPA bouts over 1-minute, smoking history, alcohol consumption, accelerometry-derived sleep duration, diet, education, ethnicity, self-reported parental history of CVD, previous incidence of cancer, self-reported medication use (cholesterol, blood pressure, and diabetes) and residual of VILPA duration of 1-minute bouts. **Panel A:** all MACE: n = 22,368; events: 819 (female/male = 331/488), **Panel B:** myocardial infarction: n = 21,928; events = 379 (female/male = 129/250). **Panel C:** heart failure: n = 21,764; events = 215 (female/male = 96/119). **Panel D:** stroke: n = 21,774; events = 225 (female/male = 106/119). Diamond, minimal dose, as indicated by the ED_50_ statistic which estimates the daily duration of VILPA associated with 50% of optimal risk reduction. Circle, HR associated with the median VILPA value (see eTable 3 for the list of values).

**eFigure 8:** Adjusted dose response curves of daily VILPA duration for MACE and its sub-types, bouts lasting up to 2 minutes (minutes/day).

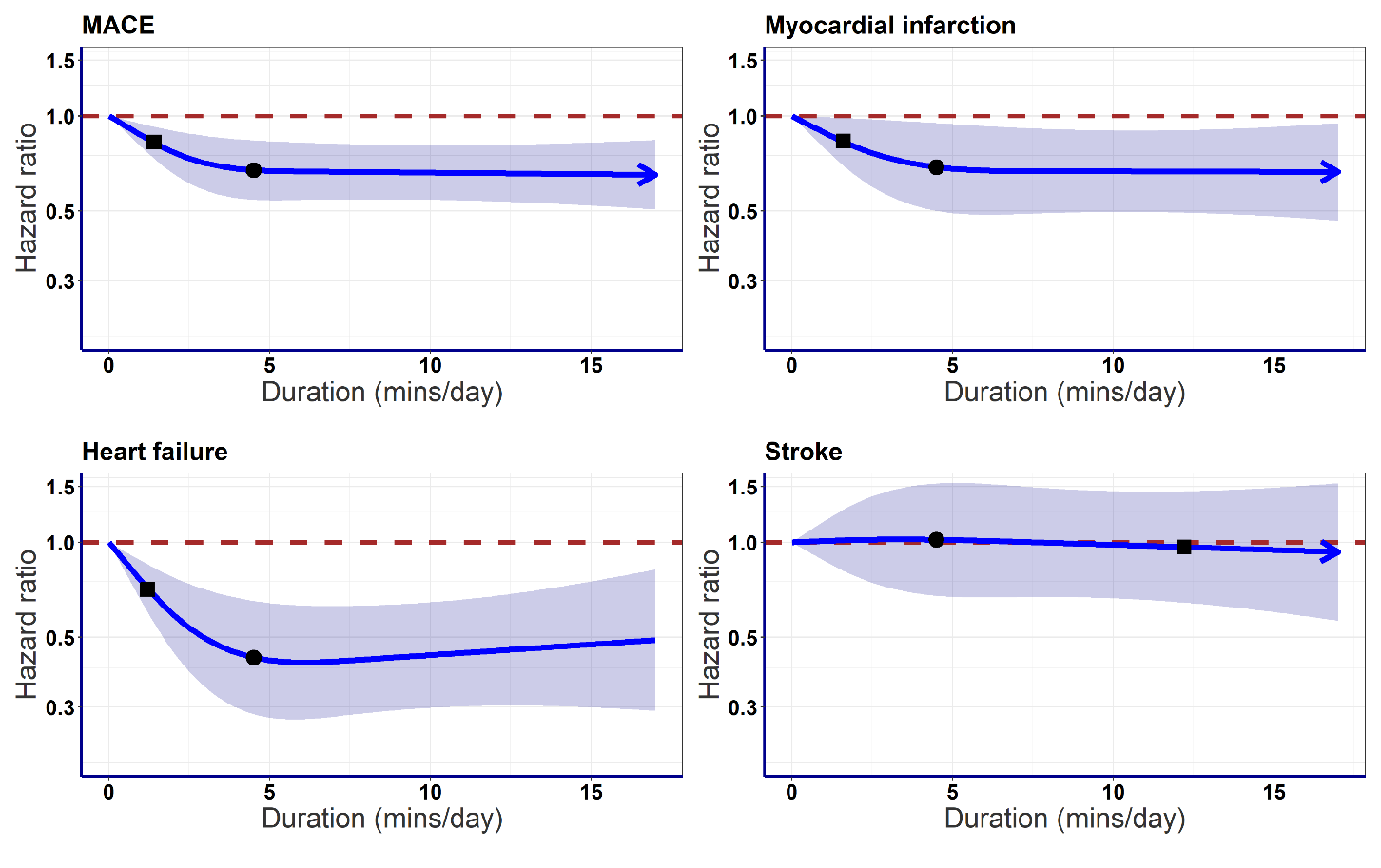

***Legend:*** Adjusted for sex, age, light intensity, moderate intensity, VILPA bouts over 1-minute, smoking history, alcohol consumption, accelerometry-derived sleep duration, diet, education, ethnicity, self-reported parental history of CVD, previous incidence of cancer, and self-reported medication use (cholesterol, blood pressure, and diabetes). **Panel A:** all MACE: n = 22,368; events: 819, **Panel B:** myocardial infarction: n = 21,928; events = 379. **Panel C:** heart failure: n = 21,764; events = 215. **Panel D:** stroke: n = 21,774; events = 225. Diamond, minimal dose, as indicated by the ED_50_ statistic which estimates the daily duration of VILPA associated with 50% of optimal risk reduction. Circle, HR associated with the median VILPA value (see eTable 3 for the list of values).

**eFigure 9:** Adjusted sex-specific dose response curves of daily VILPA duration for MACE and its sub-types, bouts lasting up to 2 minutes (minutes/day).

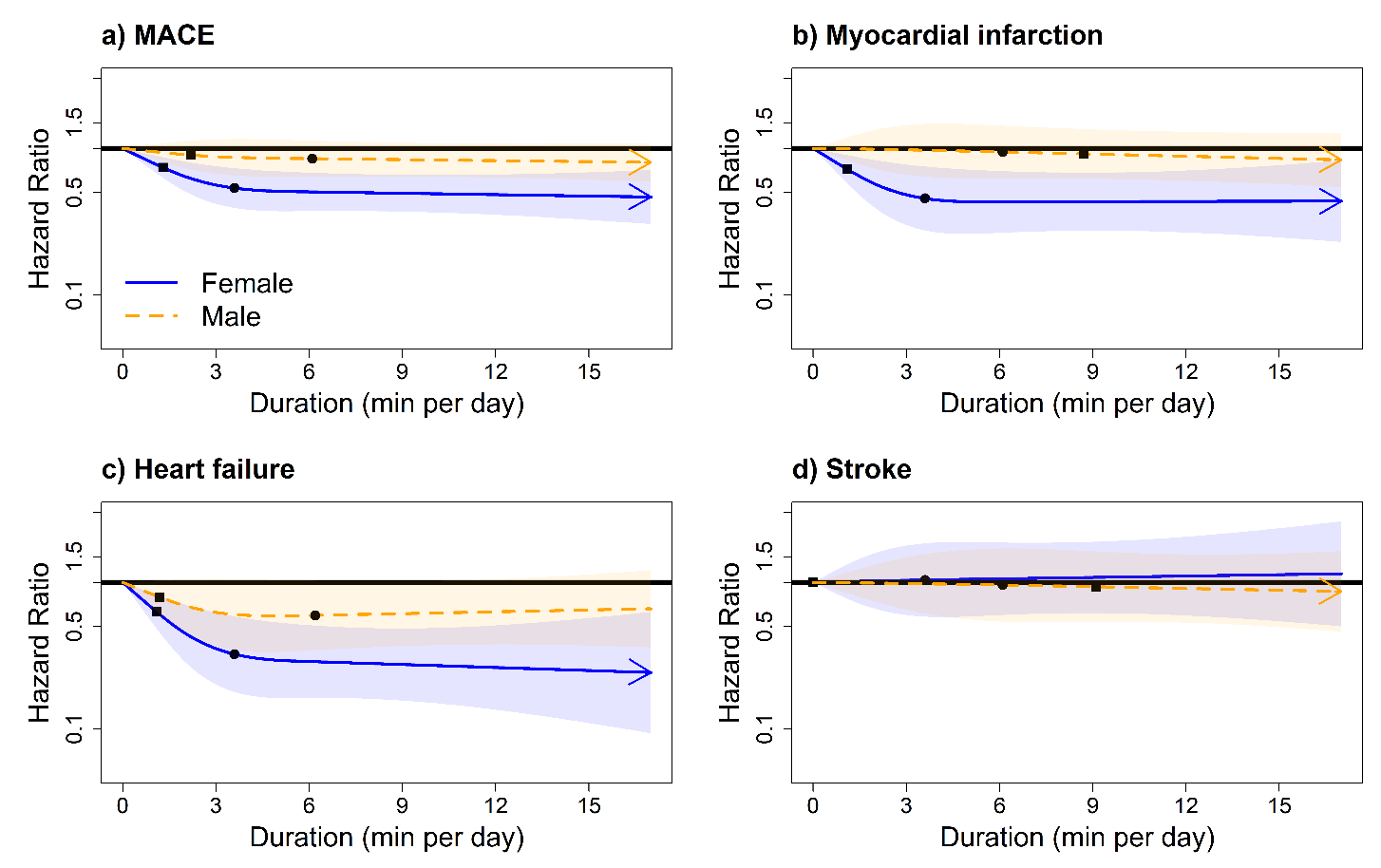

***Legend:*** Adjusted for age, light intensity, moderate intensity, VILPA bouts over 1-minute, smoking history, alcohol consumption, accelerometry-derived sleep duration, diet, education, ethnicity, self-reported parental history of CVD, previous incidence of cancer, and self-reported medication use (cholesterol, blood pressure, and diabetes). **Panel A:** all MACE: n = 22,368; events: 819 (female/male = 331/488), **Panel B:** myocardial infarction: n = 21,928; events = 379 (female/male = 129/250). **Panel C:** heart failure: n = 21,764; events = 215 (female/male = 96/119). **Panel D:** stroke: n = 21,774; events = 225 (female/male = 106/119). Diamond, minimal dose, as indicated by the ED_50_ statistic which estimates the daily duration of VILPA associated with 50% of optimal risk reduction. Circle, HR associated with the median VILPA value (see eTable 4 for the list of values).

**eFigure 10:** Adjusted dose response curves of vigorous physical activity (VPA) in exercisers for MACE and its subtypes for bouts lasting up to 1 minute (minutes/day).

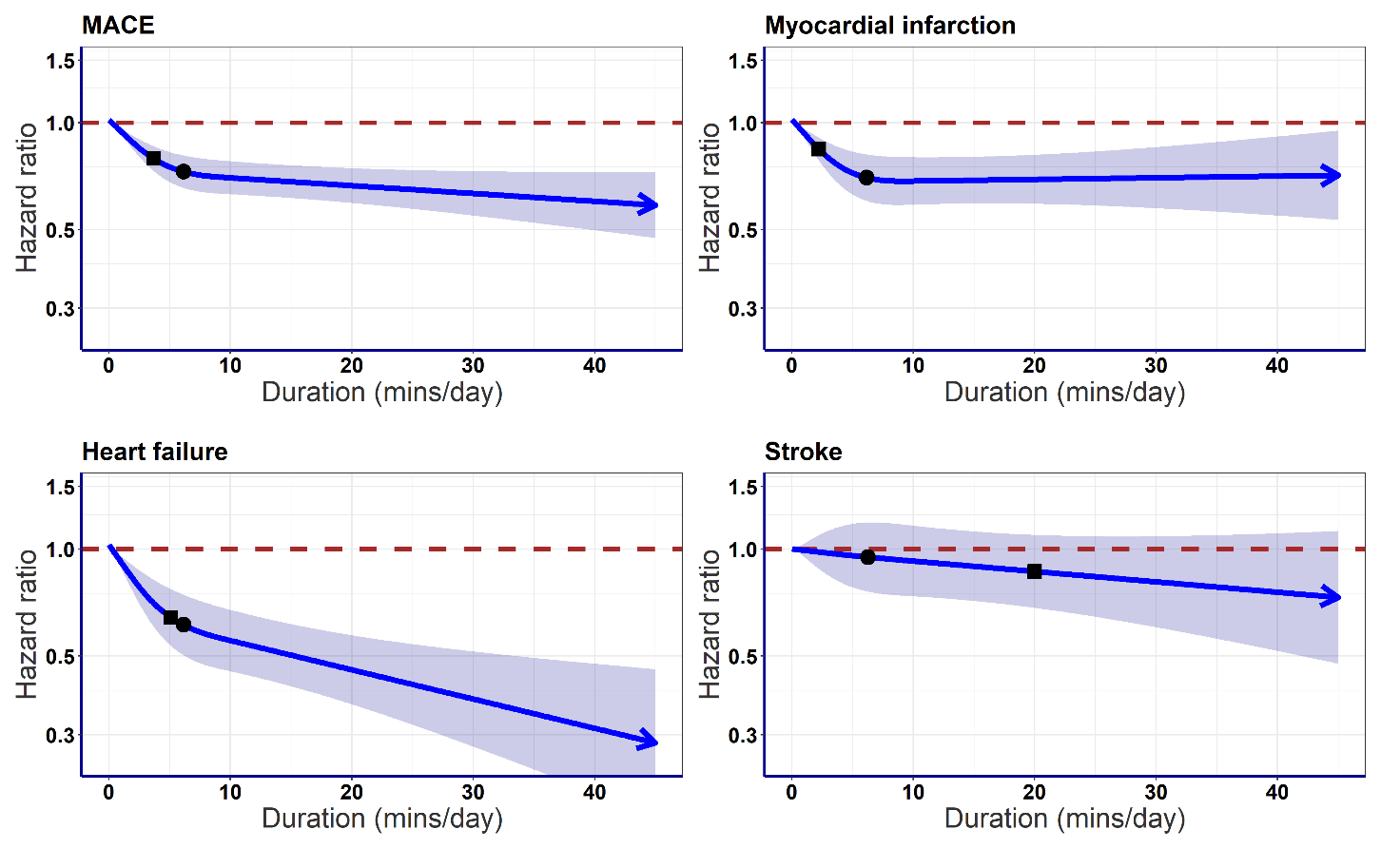

***Legend:*** Adjusted for sex, age, light intensity, moderate intensity, VILPA bouts over 1-minute, smoking history, alcohol consumption, accelerometry-derived sleep duration, diet, education, ethnicity, self-reported parental history of CVD, previous incidence of cancer, and self-reported medication use (cholesterol, blood pressure, and diabetes). **Panel A:** all MACE: n = 58,668; events: 1854, **Panel B:** myocardial infarction: n = 57,622; events = 828. **Panel C:** heart failure: n = 57,289; events = 495. **Panel D:** stroke: n = 57,325; events = 531. Diamond, minimal dose, as indicated by the ED_50_ statistic which estimates the daily duration of VILPA associated with 50% of optimal risk reduction. Circle, HR associated with the median VILPA value (see eTable 5 for the list of values).

**eFigure 11:** Sex-specific adjusted dose response curves of **length-standardised frequency of** vigorous physical activity (VPA) in exercisers for MACE and its subtypes for bouts lasting up to 1 minute (minutes/day).

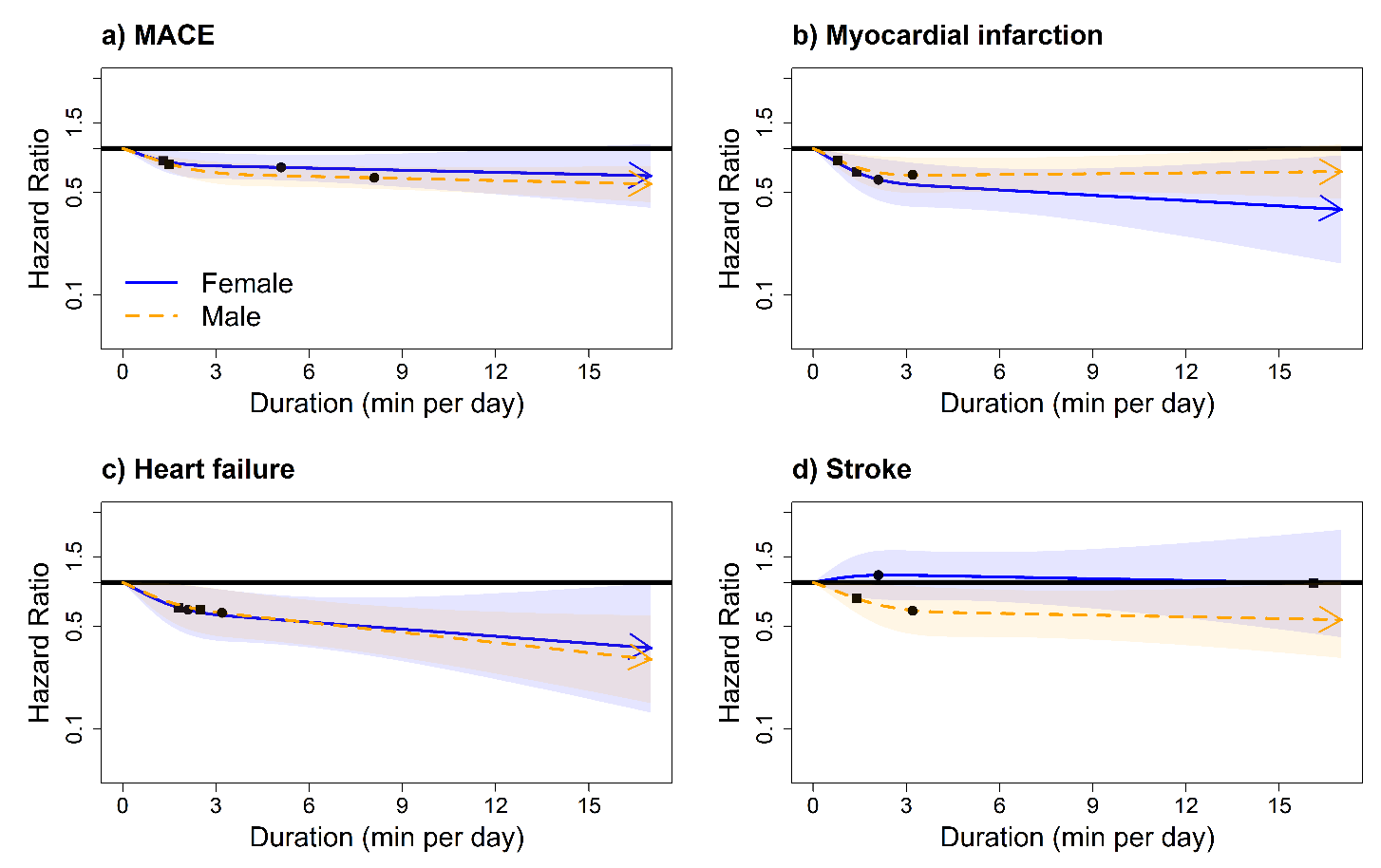

***Legend:*** Adjusted for sex, age, light intensity, moderate intensity, VILPA bouts over 1-minute, smoking history, alcohol consumption, accelerometry-derived sleep duration, diet, education, ethnicity, self-reported parental history of CVD, previous incidence of cancer, and self-reported medication use (cholesterol, blood pressure, and diabetes). The range was capped at the 97.5 percentile to minimize the influence of sparse data. **Panel A:** MACE: n = 58,648; events: 1854 (female/male = 749/1105), **Panel B:** myocardial infarction: n = 57,622; events = 828 (female/male = 287/541). **Panel C:** heart failure: n = 57,289; events = 495 (female/male = 210/285). **Panel D:** stroke: n = 57,325; events = 531 (female/male = 252/279). Diamond, minimal dose, as indicated by the ED_50_ statistic which estimates the daily duration of VILPA associated with 50% of optimal risk reduction. Circle, HR associated with the median VILPA value (see eTable 6 for the list of values).

**eFigure 12:** Sex-specific adjusted dose response curves of **raw frequency of** vigorous physical activity (VPA) in exercisers for MACE and its subtypes for bouts lasting up to 1 minute (minutes/day).

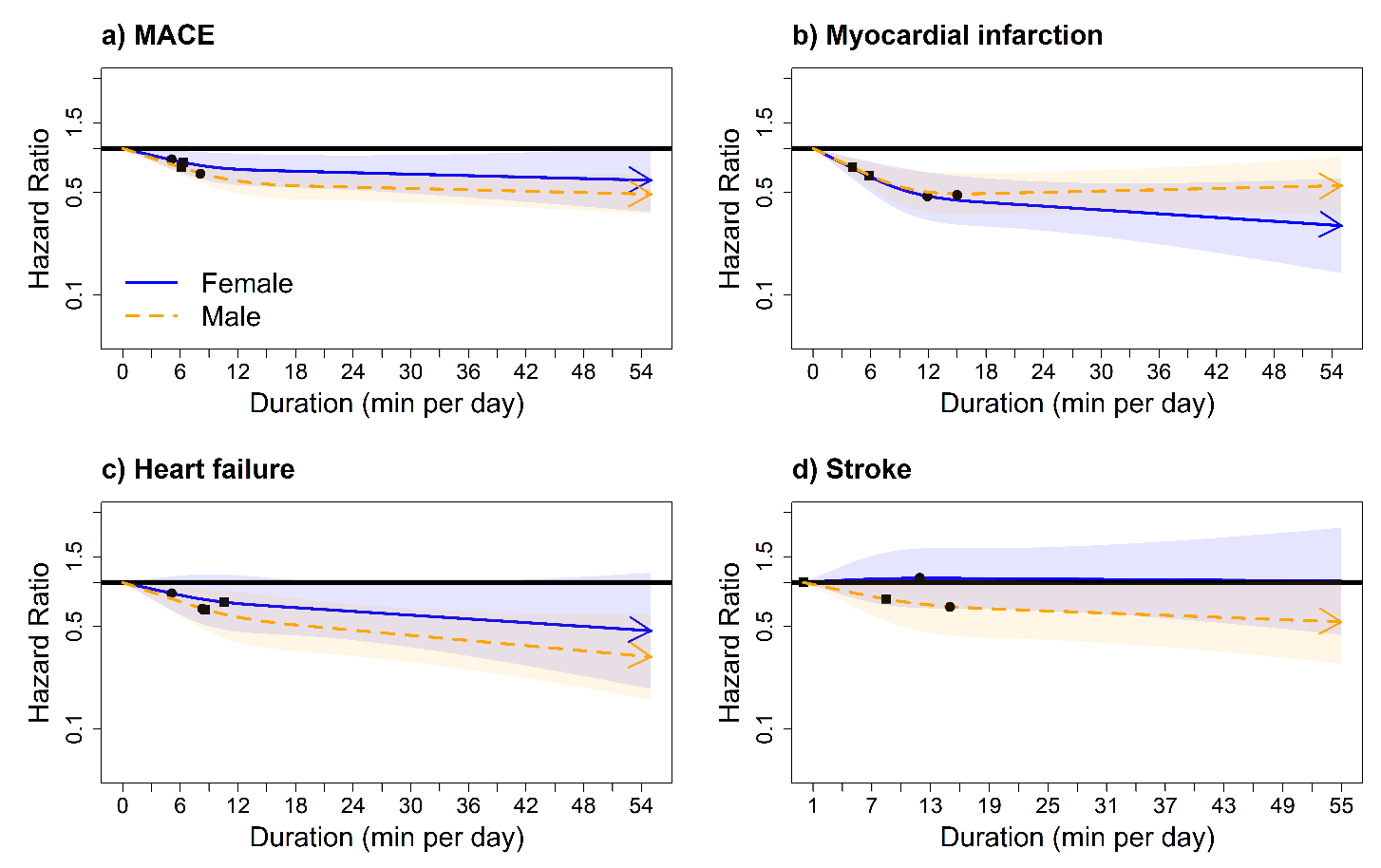

***Legend:*** Adjusted for sex, age, light intensity, moderate intensity, VILPA bouts over 1-minute, smoking history, alcohol consumption, accelerometry-derived sleep duration, diet, education, ethnicity, self-reported parental history of CVD, previous incidence of cancer, self-reported medication use (cholesterol, blood pressure, and diabetes) and residual of VILPA duration of 1-minute bouts. **Panel A:** MACE: n = 58,648; events: 1854 (female/male = 749/1105), **Panel B:** myocardial infarction: n = 57,622; events = 828 (female/male = 287/541). **Panel C:** heart failure: n = 57,289; events = 495 (female/male = 210/285). **Panel D:** stroke: n = 57,325; events = 531 (female/male = 252/279). Diamond, minimal dose, as indicated by the ED_50_ statistic which estimates the daily duration of VILPA associated with 50% of optimal risk reduction. Circle, HR associated with the median VILPA value (see eTable 6 for the list of values).

**eFigure 13:** Adjusted Sex-specific dose response curves of daily VILPA duration for MACE and its subtypes, with additional adjustment for glycated haemoglobin (HbA1c), low-density lipoprotein (LDL), high-density lipoprotein (HDL), triglycerides, systolic blood pressure, diastolic blood pressure, and body mass index; bouts lasting up to 1 minute (minutes/day).

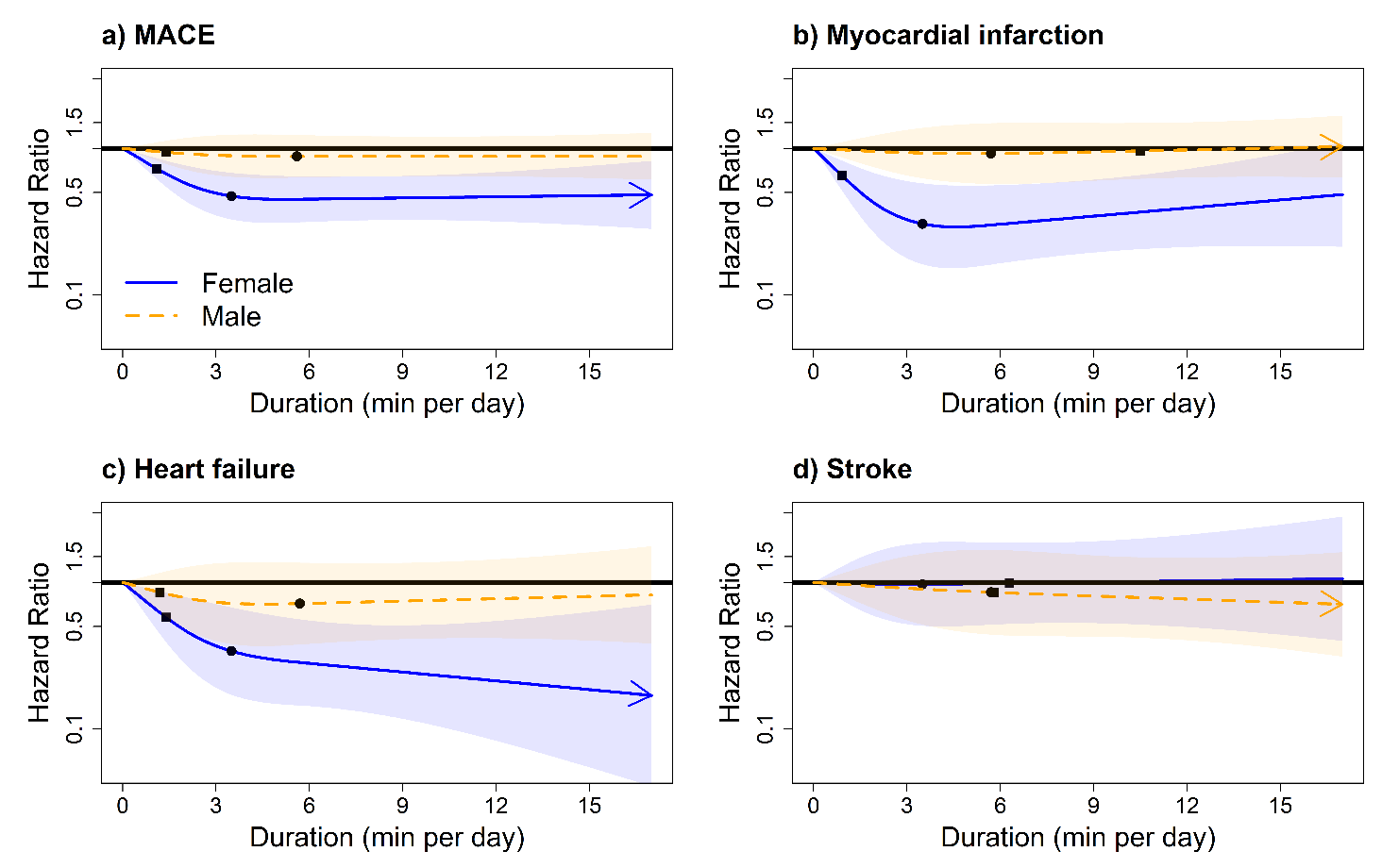

***Legend:*** Adjusted for age, light intensity, moderate intensity, VILPA bouts over 1-minute, smoking history, alcohol consumption, accelerometry-derived sleep duration, diet, education, self-reported parental history of CVD, previous incidence of cancer, self-reported medication use (cholesterol, blood pressure, and diabetes), glycated haemoglobin (HbA1c), low-density lipoprotein (LDL), high-density lipoprotein (HDL), triglycerides, systolic blood pressure, diastolic blood pressure, and body mass index. **Panel A:** all MACE: n = 22,289; events: 817 (female/male = 329/488), **Panel B:** myocardial infarction: n = 21,850; events = 378 (female/male = 128/250). **Panel C:** heart failure: n = 21,687; events = 215 (female/male = 96/119). **Panel D:** stroke: n = 20,895; events = 224 (female/male = 105/119). Diamond, minimal dose, as indicated by the ED_50_ statistic which estimates the daily duration of VILPA associated with 50% of optimal risk reduction. Circle, HR associated with the median VILPA value.

**eFigure 14:** Adjusted Sex-specific dose response curves of daily VILPA duration for MACE and its subtypes, after excluding participants with poor health or a BMI below 18.5 kg/m^2^ or current smokers.

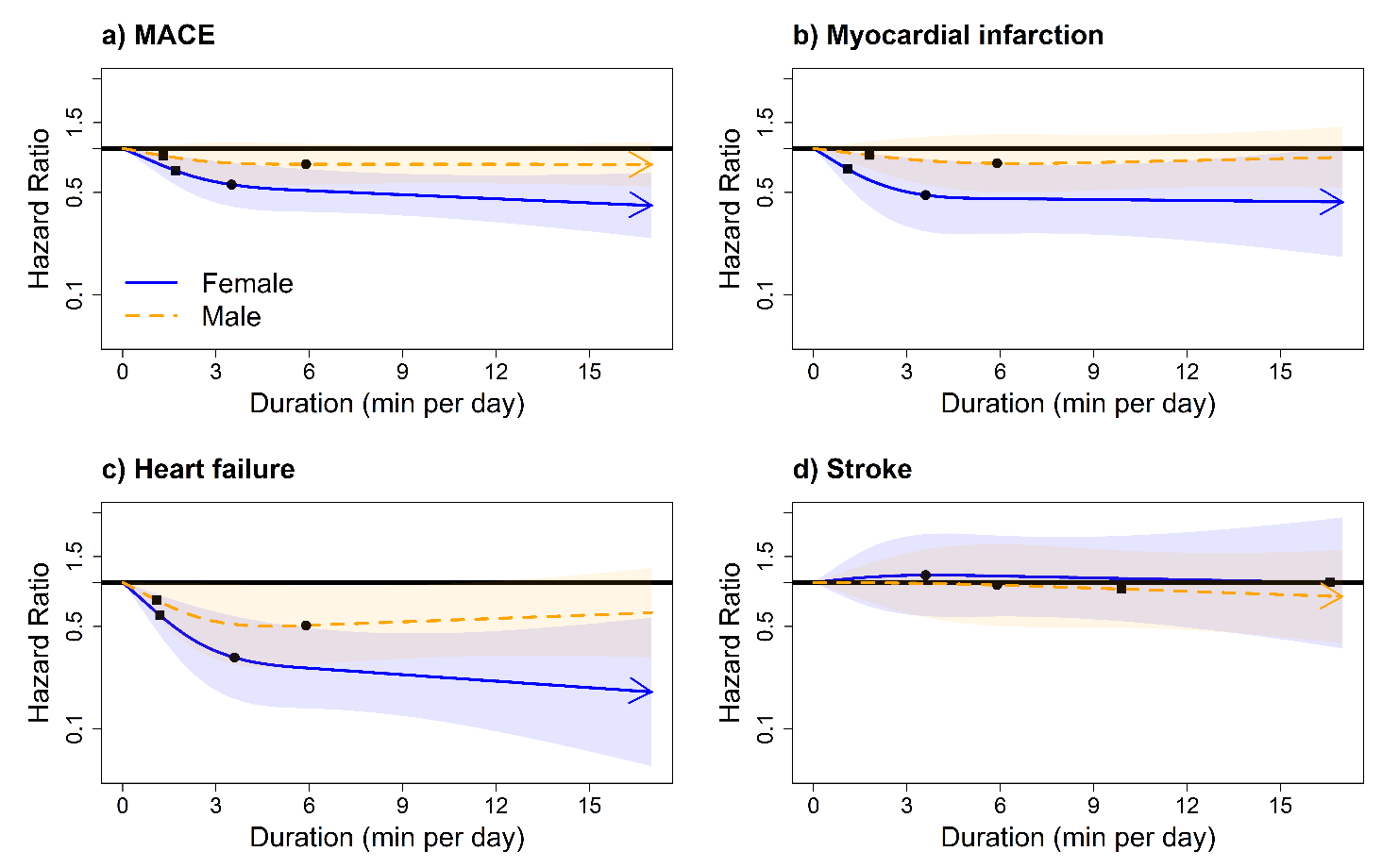

***Legend:*** Adjusted for age, light intensity, moderate intensity, VILPA bouts over 1-minute, smoking history, alcohol consumption, accelerometry-derived sleep duration, diet, education, self-reported parental history of CVD, previous incidence of cancer, and self-reported medication use (cholesterol, blood pressure, and diabetes). **Panel A:** all MACE: n = 19,471; events: 661 (female/male = 271/390), **Panel B:** myocardial infarction: n = 19,116; events = 306 (female/male = 105/201). **Panel C:** heart failure: n = 18,983; events = 173 (female/male = 80/93). **Panel D:** stroke: n = 18,348; events = 182 (female/male = 86/96). Diamond, minimal dose, as indicated by the ED_50_ statistic which estimates the daily duration of VILPA associated with 50% of optimal risk reduction. Circle, HR associated with the median VILPA value.

**eFigure 15:** Adjusted sex-specific dose response curves of VILPA duration for MACE and its sub-types, VILPA bouts lasting up to 1 minutes (bouts/day), with reference as 15^th^ percentile for VILPA duration (0.625 mins/day).

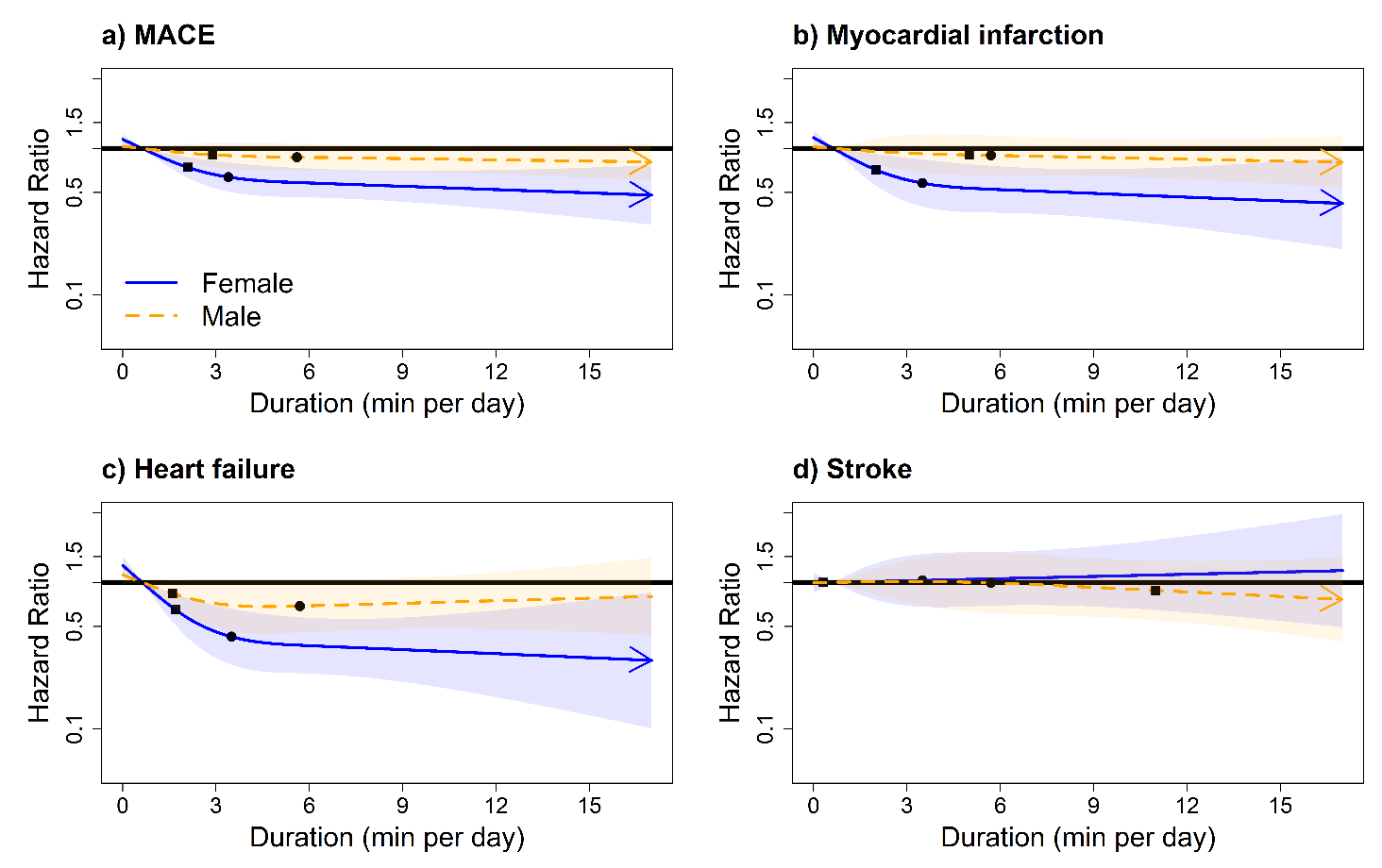

***Legend:*** Adjusted for age, light intensity, moderate intensity, VILPA bouts over 1-minute, smoking history, alcohol consumption, accelerometry-derived sleep duration, diet, education, self-reported parental history of CVD, previous incidence of cancer, and self-reported medication use (cholesterol, blood pressure, and diabetes). **Panel A:** all MACE: n = 19,471; events: 661 (female/male = 271/390), **Panel B:** myocardial infarction: n = 19,116; events = 306 (female/male = 105/201). **Panel C:** heart failure: n = 18,983; events = 173 (female/male = 80/93). **Panel D:** stroke: n = 18,348; events = 182 (female/male = 86/96). Diamond, minimal dose, as indicated by the ED_50_ statistic which estimates the daily duration of VILPA associated with 50% of optimal risk reduction. Circle, HR associated with the median VILPA value.

**eFigure 16:** Adjusted sex-specific dose response curves of VILPA duration for MACE and its sub-types, VILPA bouts lasting up to 1 minutes (bouts/day), excluding frail participants (n = 1,924).

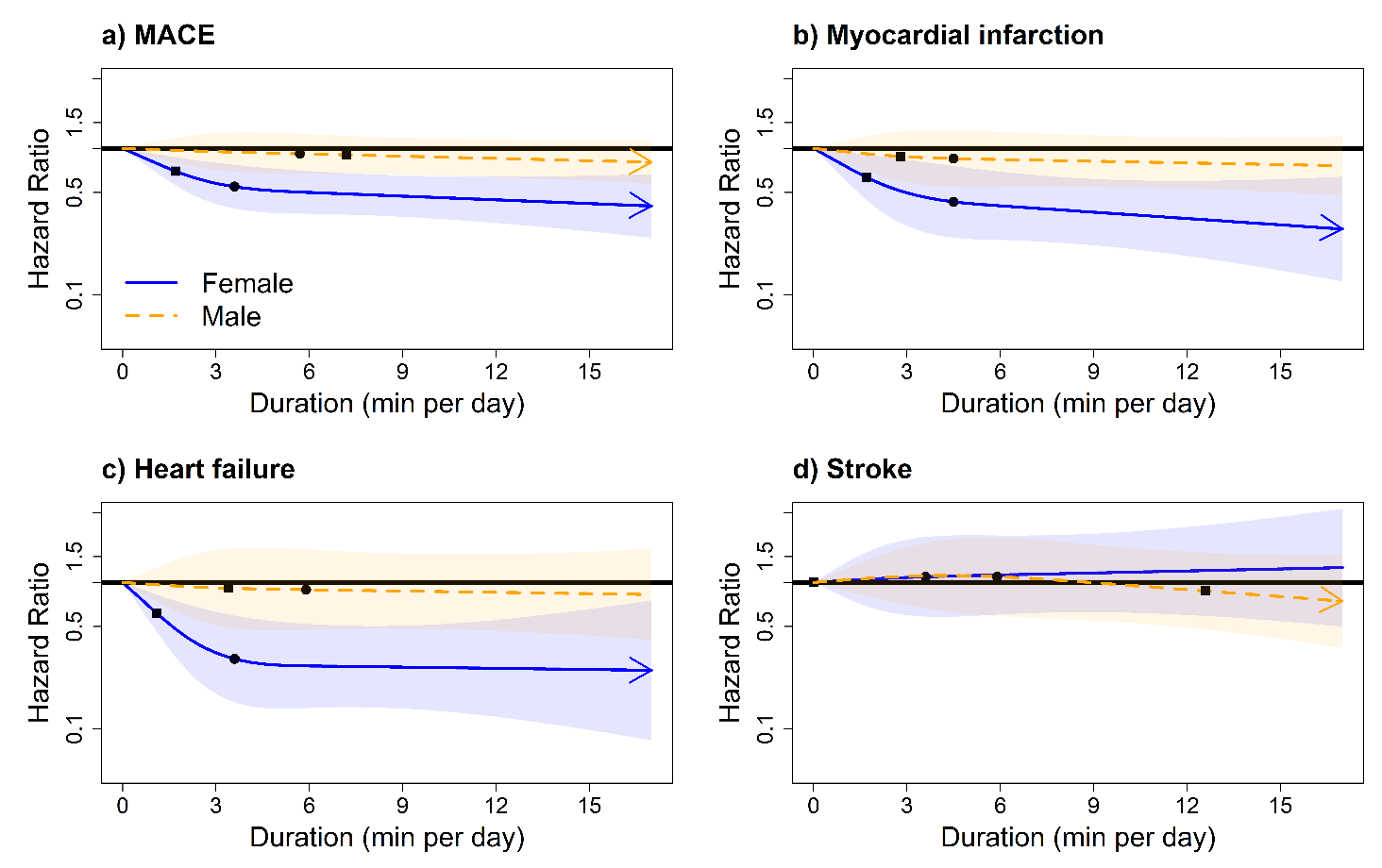

***Legend:*** Adjusted for age, light intensity, moderate intensity, VILPA bouts over 1-minute, smoking history, alcohol consumption, accelerometry-derived sleep duration, diet, education, self-reported parental history of CVD, previous incidence of cancer, and self-reported medication use (cholesterol, blood pressure, and diabetes). **Panel A:** all MACE: n = 19,465 events: 697 (female/male = 271/426), **Panel B:** myocardial infarction: n = 19,096; events = 328 (female/male = 105/223). **Panel C:** heart failure: n = 18,940; events = 172 (female/male = 73/99). **Panel D:** stroke: n = 18,965; events = 197 (female/male = 93/104). Diamond, minimal dose, as indicated by the ED_50_ statistic which estimates the daily duration of VILPA associated with 50% of optimal risk reduction. Circle, HR associated with the median VILPA value.

**eFigure 17:** Age-stratified and sex-specific dose response curves of daily VILPA duration for MACE and its subtypes for bouts lasting up to 1 minute (minutes/day).

**A) Age below median** (42.9 - 62.4 years)

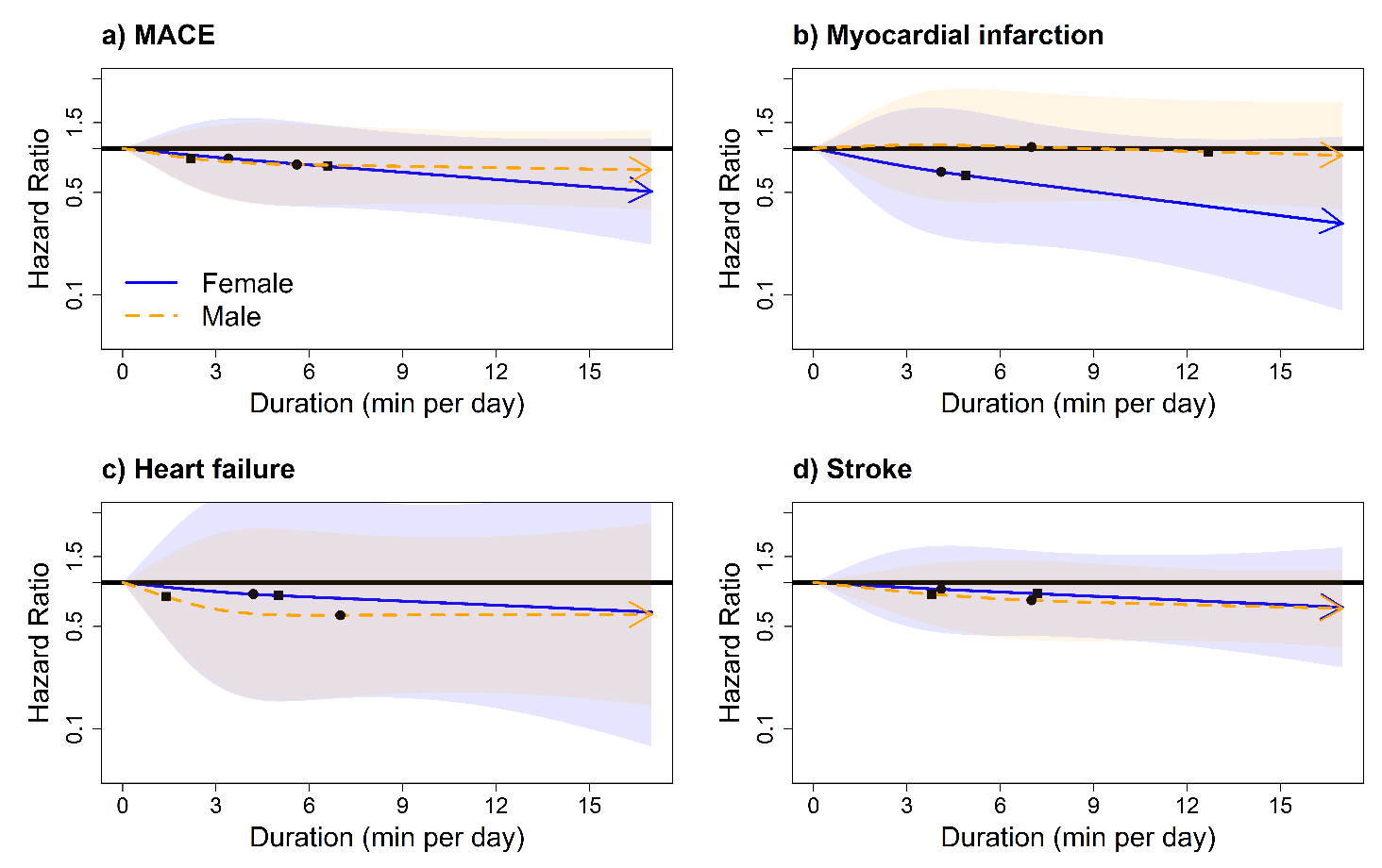

***Legend:*** Adjusted for age, light intensity, moderate intensity, VILPA bouts over 1-minute, smoking history, alcohol consumption, accelerometry-derived sleep duration, diet, education, self-reported parental history of CVD, previous incidence of cancer, and self-reported medication use (cholesterol, blood pressure, and diabetes). **Panel A:** all MACE: n = 10,695; events: 201 (female/male = 79/122), **Panel B:** myocardial infarction: n = 10,601; events = 107 (female/male = 33/74). **Panel C:** heart failure: n = 10,531; events = 37 (female/male = 13/24). **Panel D:** stroke: n = 10,551; events = 57 (female/male = 33/24). Diamond, minimal dose, as indicated by the ED_50_ statistic which estimates the daily duration of VILPA associated with 50% of optimal risk reduction. Circle, HR associated with the median VILPA value.

**B) Age above median** (62.4 -78.6 years**)**

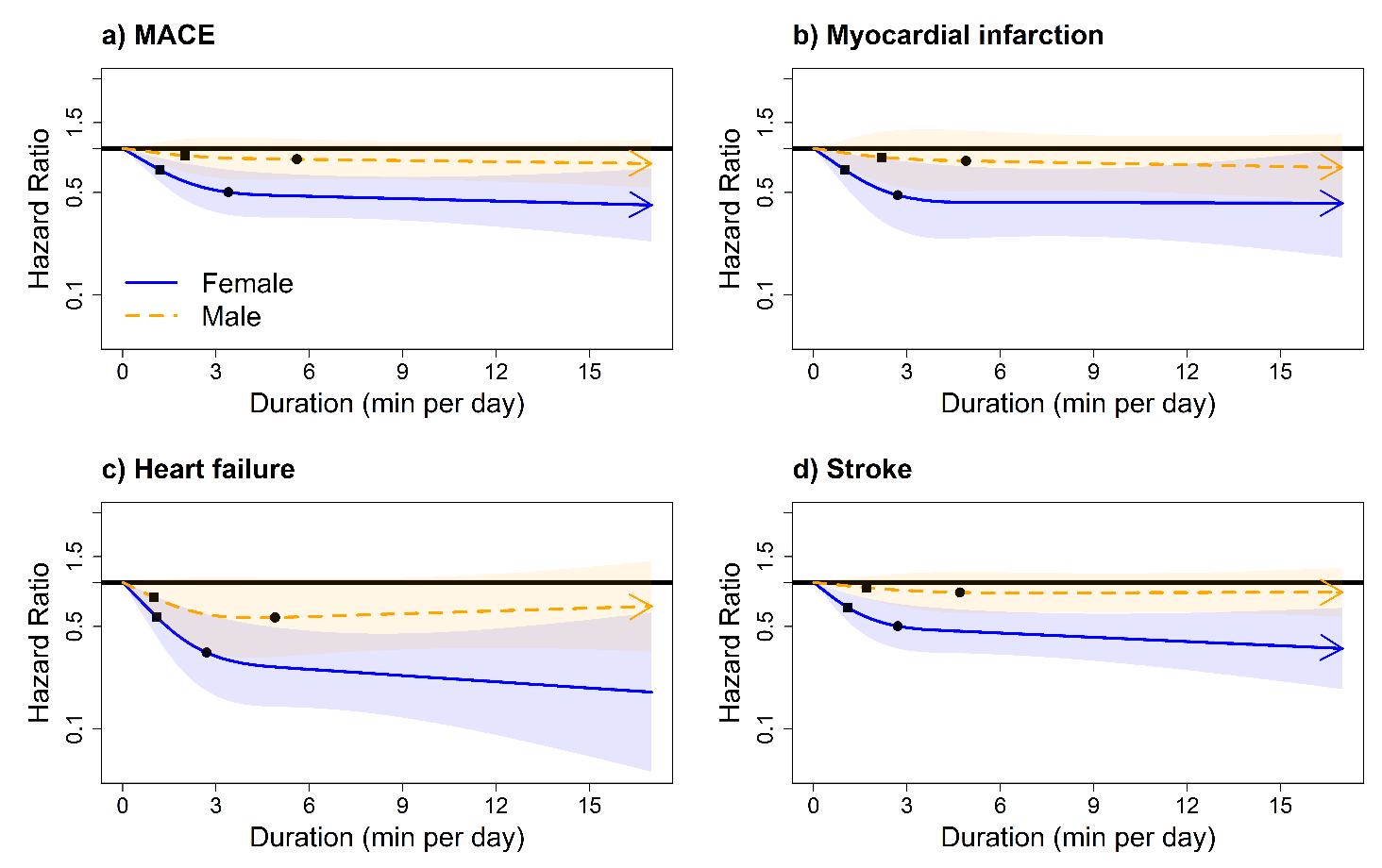

***Legend:*** Adjusted for age, light intensity, moderate intensity, VILPA bouts over 1-minute, smoking history, alcohol consumption, accelerometry-derived sleep duration, diet, education, self-reported parental history of CVD, previous incidence of cancer, and self-reported medication use (cholesterol, blood pressure, and diabetes). **Panel A:** all MACE: n = 11,673; events: 618 (female/male = 252/366), **Panel B:** myocardial infarction: n = 11,327; events = 272 (female/male = 96/176). **Panel C:** heart failure: n = 10,602; events = 178 (female/male = 83/95). **Panel D:** stroke: n = 11,223; events = 168 (female/male = 73/95). Diamond, minimal dose, as indicated by the ED_50_ statistic which estimates the daily duration of VILPA associated with 50% of optimal risk reduction. Circle, HR associated with the median VILPA value.

**eFigure 18:** Adjusted Sex-specific dose response curves of light intensity physical activity for MACE and its subtypes for non-exercisers.

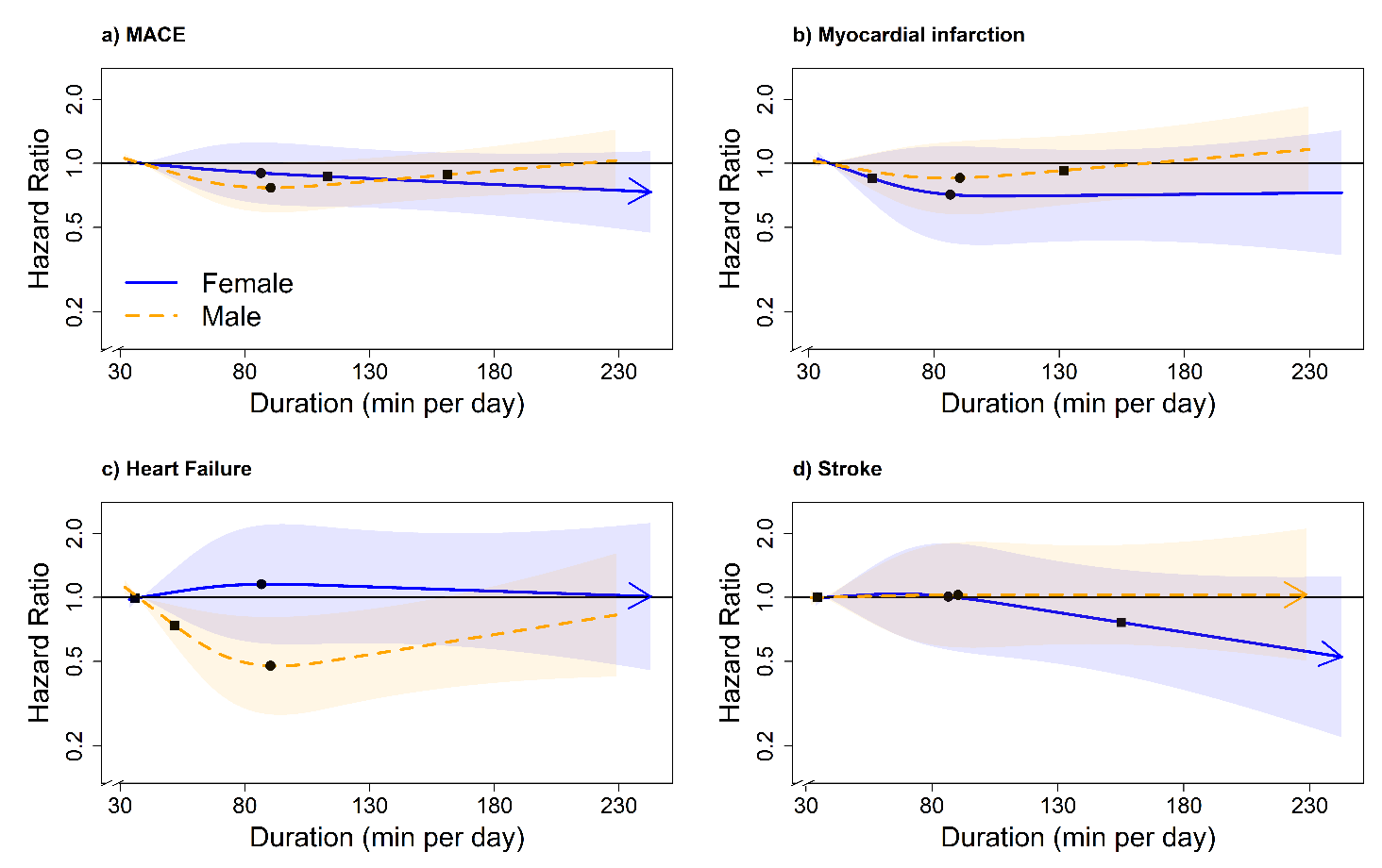

***Legend:*** Adjusted for age, moderate intensity, vigorous intensity, smoking history, alcohol consumption, accelerometry-derived sleep duration, diet, education, ethnicity, self-reported parental history of CVD, previous incidence of cancer, self-reported medication use (cholesterol, blood pressure, and diabetes) and residual of VILPA duration of 1-minute bouts. **Panel A:** all MACE: n = 22,368; events: 819 (female/male = 331/488), **Panel B:** myocardial infarction: n = 21,928; events = 379 (female/male = 129/250). **Panel C:** heart failure: n = 21,764; events = 215 (female/male = 96/119). **Panel D:** stroke: n = 21,774; events = 225 (female/male = 106/119). Diamond, minimal dose, as indicated by the ED_50_ statistic which estimates the daily duration of VILPA associated with 50% of optimal risk reduction. Circle, HR associated with the median VILPA value (see eTable 4 for the list of values).

**eFigure 19:** Adjusted Sex-specific dose response curves of moderate intensity physical activity for MACE and its subtypes for non-exercisers.

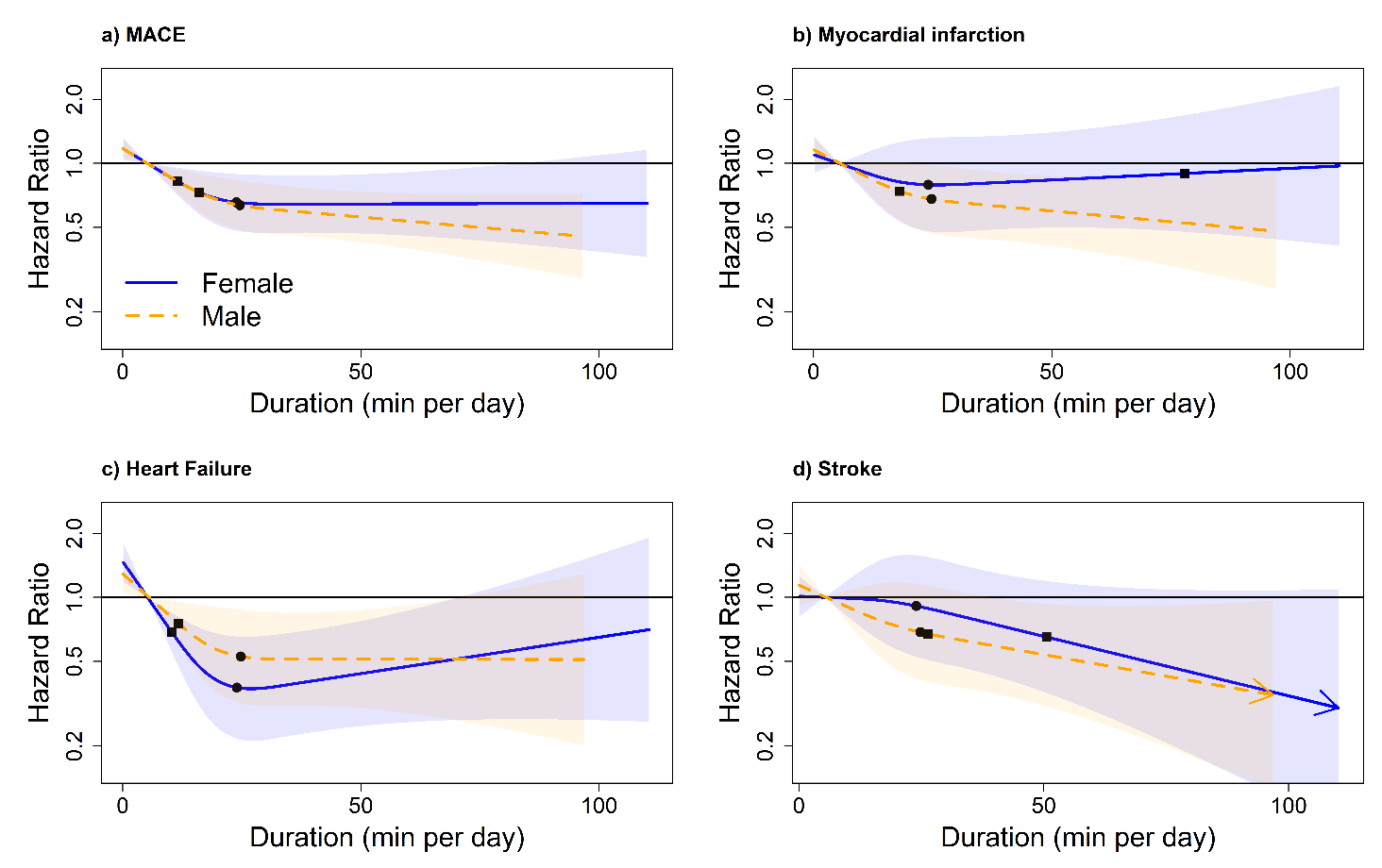

***Legend:*** Adjusted for age, light intensity, vigorous intensity, smoking history, alcohol consumption, accelerometry-derived sleep duration, diet, education, ethnicity, self-reported parental history of CVD, previous incidence of cancer, self-reported medication use (cholesterol, blood pressure, and diabetes) and residual of VILPA duration of 1-minute bouts. **Panel A:** all MACE: n = 22,368; events: 819 (female/male = 331/488), **Panel B:** myocardial infarction: n = 21,928; events = 379 (female/male = 129/250). **Panel C:** heart failure: n = 21,764; events = 215 (female/male = 96/119). **Panel D:** stroke: n = 21,774; events = 225 (female/male = 106/119). Diamond, minimal dose, as indicated by the ED_50_ statistic which estimates the daily duration of VILPA associated with 50% of optimal risk reduction. Circle, HR associated with the median VILPA value (see eTable 4 for the list of values).

**eFigure 20:** Adjusted Sex-specific dose response curves of light intensity physical activity for MACE and its subtypes for exercisers.

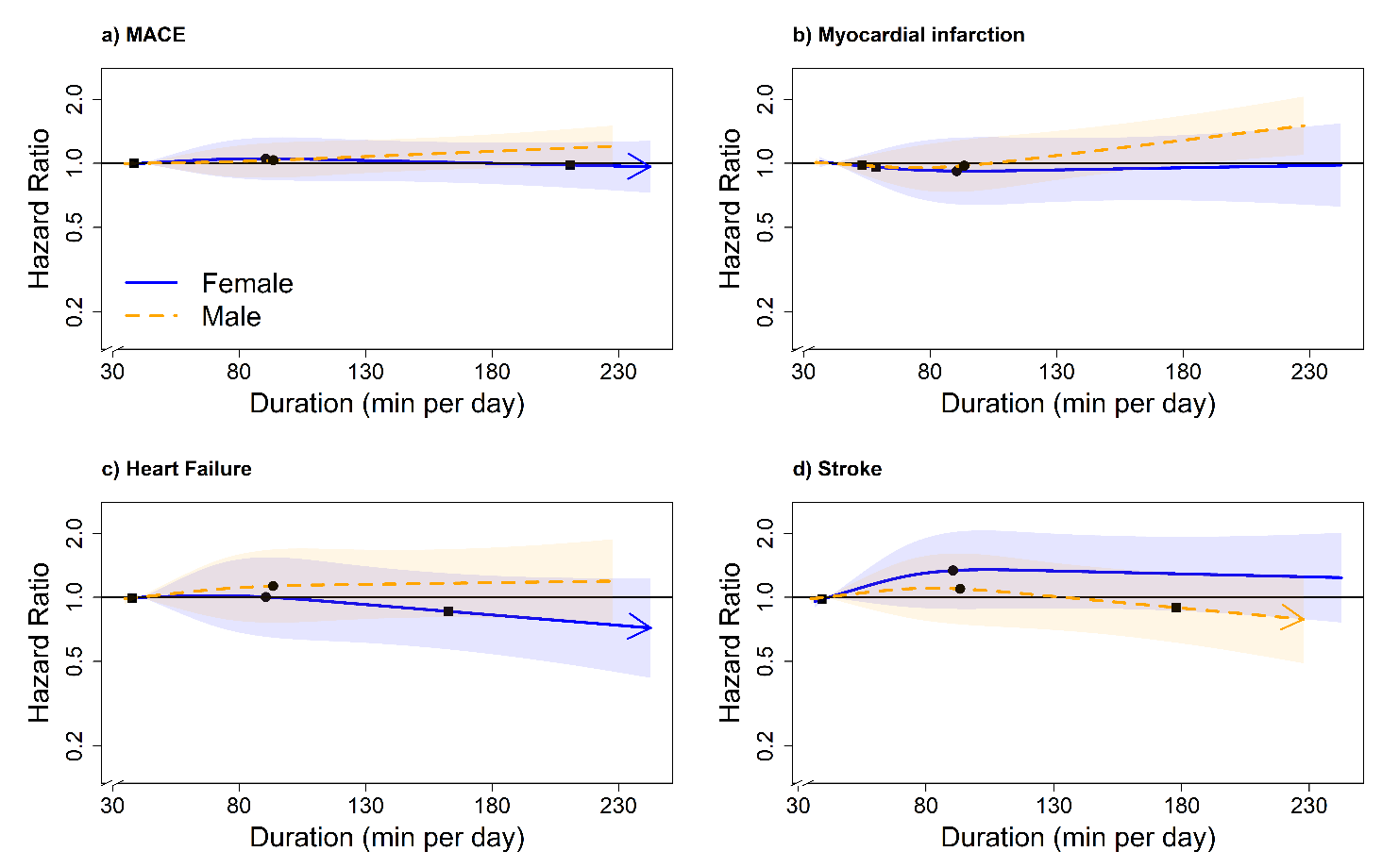

***Legend:*** Adjusted for sex, age, moderate intensity, vigorous intensity, smoking history, alcohol consumption, accelerometry-derived sleep duration, diet, education, ethnicity, self-reported parental history of CVD, previous incidence of cancer, and self-reported medication use (cholesterol, blood pressure, and diabetes). The range was capped at the 97.5 percentile to minimize the influence of sparse data. **Panel A:** MACE: n = 58,648; events: 1854 (female/male = 749/1105), **Panel B:** myocardial infarction: n = 57,622; events = 828 (female/male = 287/541). **Panel C:** heart failure: n = 57,289; events = 495 (female/male = 210/285). **Panel D:** stroke: n = 57,325; events = 531 (female/male = 252/279). Diamond, minimal dose, as indicated by the ED_50_ statistic which estimates the daily duration of VILPA associated with 50% of optimal risk reduction. Circle, HR associated with the median VILPA value.

**eFigure 21:** Adjusted Sex-specific dose response curves of moderate intensity physical activity for MACE and its subtypes for exercisers.

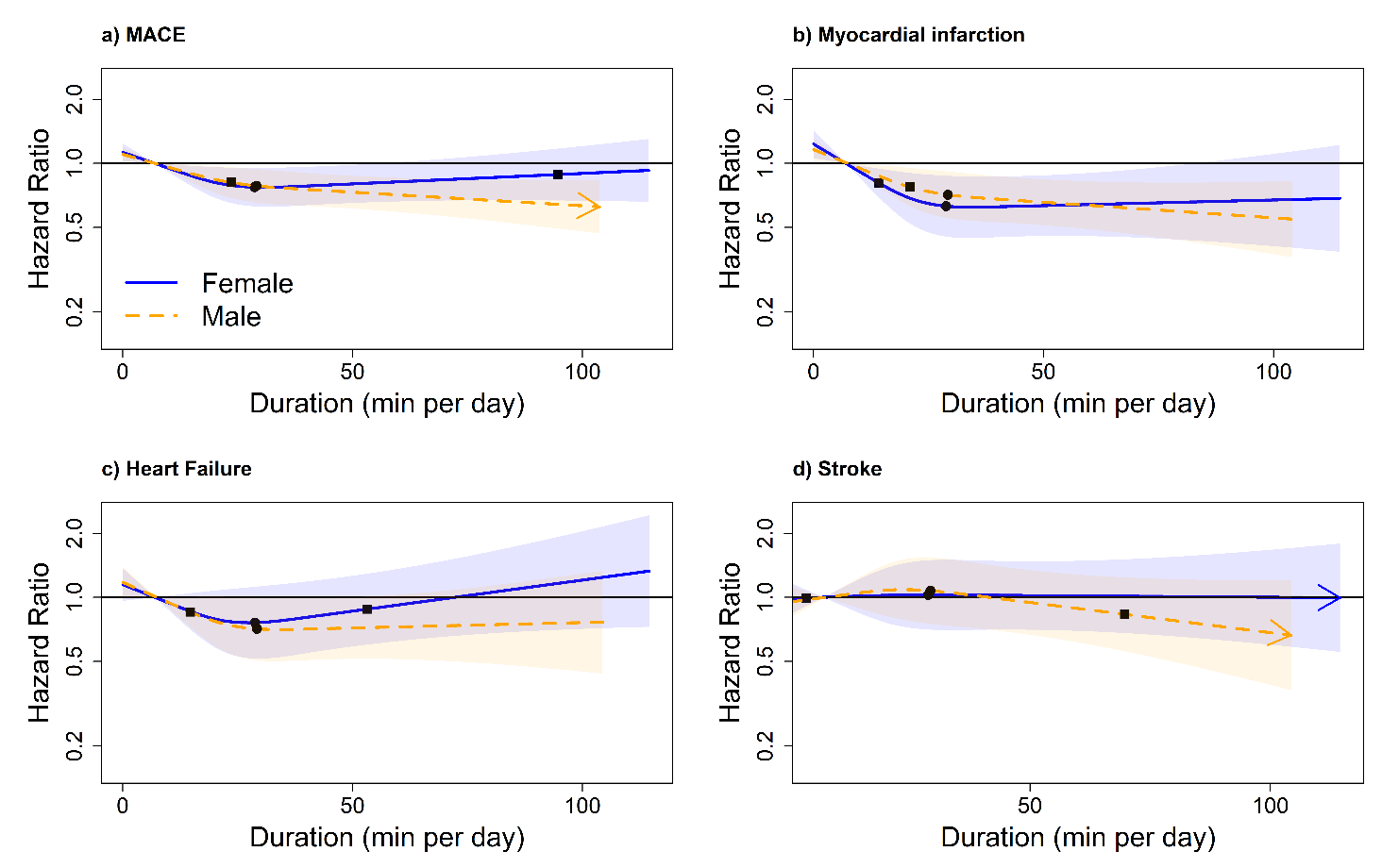

***Legend:*** Adjusted for sex, age, light intensity, moderate intensity, smoking history, alcohol consumption, accelerometry-derived sleep duration, diet, education, ethnicity, self-reported parental history of CVD, previous incidence of cancer, and self-reported medication use (cholesterol, blood pressure, and diabetes). The range was capped at the 97.5 percentile to minimize the influence of sparse data. **Panel A:** MACE: n = 58,648; events: 1854 (female/male = 749/1105), **Panel B:** myocardial infarction: n = 57,622; events = 828 (female/male = 287/541). **Panel C:** heart failure: n = 57,289; events = 495 (female/male = 210/285). **Panel D:** stroke: n = 57,325; events = 531 (female/male = 252/279). Diamond, minimal dose, as indicated by the ED_50_ statistic which estimates the daily duration of VILPA associated with 50% of optimal risk reduction. Circle, HR associated with the median VILPA value.

**eText 1:** Physical activity classification

Physical activity was classified using a previously validated wrist-worn accelerometery Random Forest (RF) activity classifier.^1^ RF is an ensemble of multiple decision trees. Each tree is learned on a bootstrap sample of training data and each node in the tree is split using the best among a randomly selected set of acceleration features. The decisions from each tree are aggregated and a final model prediction is based on majority vote. The RF model requires very little pre-processing of the data, as the features do not need to be normalized. Additionally, the model is resistant to over fitting the training data because each tree within the forest is independently grown to maximum depth using a randomly selected subset of features.

The classifier categorized physical activity in 10 second windows into 1 of 4 activity classes: sedentary, standing utilitarian movements (ironing a shirt, washing dishes), walking (gardening, active commuting, mopping floors), and running/high energetic activities (active playing with children). These activities were then assigned to 1 of 4 activity intensities: sedentary, light, moderate, and vigorous. Walking activities were classified as light (an acceleration value of <100mg), moderate (≥100mg), and vigorous (≥400mg). The figure below depicts the activity classification scheme. Differentiation from sleep^2^ and non-wear^3^ was identified using the change in tilt angle and acceleration standard deviation. Monitors were calibrated^4^ and corrected for orientation^5^ using previously published methods.

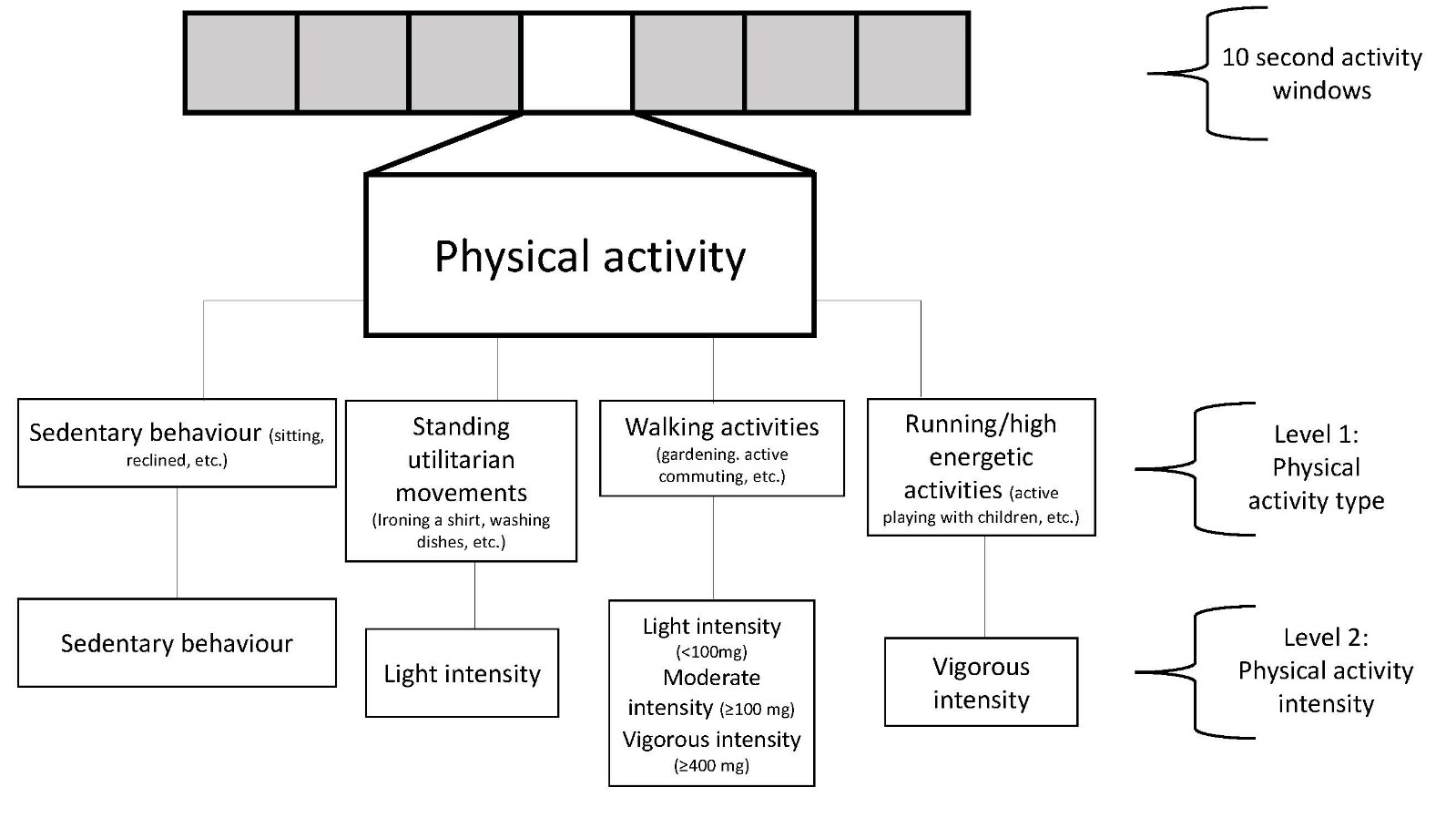
Activity classification scheme

Activities in an independent sample of 102 participants (Age = 55.8 ± 12.4; 55.8% female) from the US^6^ and Australia^7^ (includes published and unpublished data) providing 105,767 activity samples from structured and free-living activities (17,627 minutes) were used to assess robustness and generalizability of the classifier. For free-living activities participant-worn or researcher-held Go-Pro video-recordings were used to attain ground-truth physical activity. Video files were imported into the Noldus Observer XT software for continuous direct observation coding. A two-stage direct observation scheme was implemented in which the participant’s movement behaviour was coded for activity type and then activity intensity based on Compendium of Physical Activities^8^. The direct observation system generated a vector of date-time stamps corresponding to the start and finish of each movement event, which were used to assign the activity codes to the corresponding time segments of the accelerometer data. Interobserver reliability was assessed by dual coding. The intraclass correlation coefficient for coding activities was 0.912 (0.866-0.942). We present the classification and confusion matrix results below in **eText Tables 1 and 2**.

From a subset of the participants who had both ground-truth physical activity type-specific data ascertained through video recordings and the physical activity compendium, and physical activity intensity using indirect calorimetry (both gold-standard measures) we assessed intensity classification for individual activity types presented in the “expanded confusion matrix” below (**eText Table 3**; n=91; 245,945 seconds of activity data). Because of the >100 specific types of physical activity recorded we present the 3 most prevalent activity types for each intensity category.

We also provide a confusion matrix in **eText Table 4** below showing the performance among participants who self-reported as non-exercisers in the independent validity testing (n=88; 221,640 seconds of activity data).

Performance was further evaluated in a sample of 151 adults (age range 18-91 years, 65.6% female) recruited from the UK^9^. Participants in this dataset wore body cameras that provided pictures every 20 seconds to annotate ground-truth free-living activity labels. The picture-based activity coding scheme has been previously described^9^. Due to activities being coded once every 20 seconds, previously used methods in this dataset of consistent activity codes for at least 10 minutes were used to extract time segments. A total of 172,360 activity samples (28,727 minutes) were provided by participants. We present these results below in **eText Figure 1**.

Classifier performance in the three datasets is provided below:

**eText Table 1**: Intensity classification performance

|  | Sensitivity | Specificity | Precision | F-score | Overall Accuracy | Weighted Kappa | Overall F-score |
| --- | --- | --- | --- | --- | --- | --- | --- |
| Sedentary | 86.5 | 93.7 | 90.5 | 88.5 |  |  |  |
| Light | 71.2 | 89.4 | 55.8 | 62.6 |  |  |  |
| Moderate | 85.4 | 96.6 | 92.7 | 88.9 |  |  |  |
| Vigorous | 95.4 | 99.4 | 94.6 | 95.0 |  |  |  |
|  |  |  |  |  | 84.6 | 0.78 | 83.8 |

**eText Table 2**: Confusion matrix

|  | Sedentary | Light | Moderate | Vigorous |
| --- | --- | --- | --- | --- |
| Sedentary | **36,904** | 5,232 | 508 | 2 |
| Light | 3,120 | **11,712** | 1,612 | 17 |
| Moderate | 502 | 4,016 | **29,528** | 526 |
| Vigorous | 226 | 17 | 214 | **9,470** |
| Rows= ground truth; columns=predictions; bold=correct labels; numbers represent each 10-second window; Derived from the US and Australian datasets | | | | |

**eText Table 3:** Expanded confusion matrix of most prominent activity types within each intensity band in the independent validity testing (n=91; 245,945 seconds of activity data)

|  |  | **Sedentary** | **Light** | **Moderate** | **Vigorous** |
| --- | --- | --- | --- | --- | --- |
| **Sedentary** |  |  |  |  |  |
|  | Desk/computer work | **90.2%** | 9.8% | - | - |
|  | Sitting/lying | **93.9%** | 6.1% | - | - |
|  | Sitting using phone/appliance | **89.4%** | 10.6% | - | - |
| **Light** |  |  |  |  |  |
|  | Washing dishes/kitchen activities | 17.3% | **80.8%** | 2.9% | - |
|  | Household chores standing | 12.5% | **83.6%** | 3.9% | - |
|  | slow walking/grocery shopping | 15.2% | **77.8%** | 7.0% | - |
| **Moderate** |  |  |  |  |  |
|  | Walking briskly/fast (e.g. for transportation) | - | 11.4% | **86.2%** | 2.4% |
|  | Household chores cleaning rooms/ambulation | - | 12.2% | **87.6%** | 0.2% |
|  | Occupation brisk walking carrying light objects | - | 11.8% | **87.5%** | 0.7% |
| **Vigorous** |  |  |  |  |  |
|  | Manual work | - | 3.0% | 1.3% | **95.7%** |
|  | Very fast walking/burst of running (e.g for transportation) | - | 0.8% | 2.3% | **96.9%** |
|  | Heavy household outdoor chores | - | 2.4 | 4.2% | **93.4%** |

Rows= ground truth; columns=predictions; bold=correct classification

**eText Table 4a:** Confusion matrix among female participants who identified as non-exercisers in the independent validity testing (n=44; 115,142 seconds of activity data)

|  | Sedentary | Light | Moderate | Vigorous |
| --- | --- | --- | --- | --- |
| Sedentary | **92.6** | 6.9 | 0.5 | - |
| Light | 15.4 | **78.5** | 5.8 | 0.3 |
| Moderate | 0.6 | 13.4 | **85.4** | 0.6 |
| Vigorous | - | 1.2% | 1.9% | **96.9%** |
| Rows= ground truth; columns=predictions; bold=correct classification; **eText Table 4b:** Confusion matrix among male participants who identified as non-exercisers in the independent validity testing (n=44; 106,498 seconds of activity data) \| Sedentary \| Light \| Moderate \| Vigorous; Sedentary \| **89.9** \| 9.7 \| 0.4 \| -; Light \| 14.2 \| **79.2** \| 6.1 \| 0.5; Moderate \| 0.6 \| 11.1 \| **87.4** \| 0.9; Vigorous \| - \| 2.5 \| 2.9 \| **94.6**; Rows= ground truth; columns=predictions; bold=correct classification; \| \| \| \| | | | | |

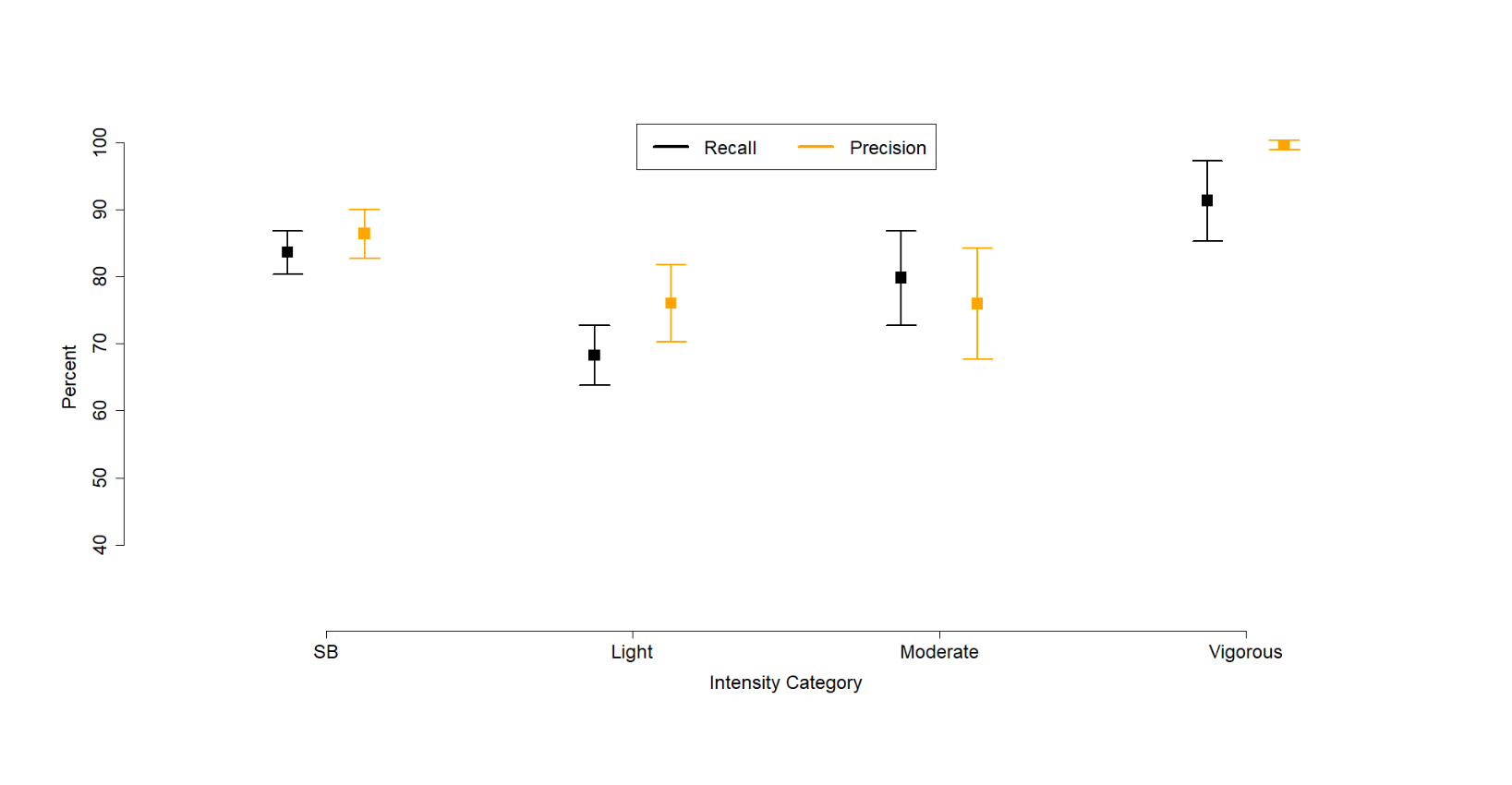
**eText Figure 1:** Participant-level specific sensitivity and precision in the UK sample

**eText 1** References:

**eText 2:** Bout length standardisation

As the length of raw bouts within the VILPA frequency exposure was highly variable, we length-standardized analytic bouts to one minute (for raw bouts lasting up to 1 minute) using a rolling sum on the time-series data until 1 or 2 minutes, respectively, was reached or exceeded. For example, a participant with five consecutive raw bouts lasting up to 1 minute each (20, 30, 20, 40, and 10 seconds long), would be assigned 1.83 analytic bouts: the first three raw bouts would count as one and the rolling sum would be reset; then the last two raw counts would count as 0.83 length-standardised bouts (50 seconds divided by 60). This bout handling has interpretational advantages as it permits a more concrete behavioural interpretation of the VILPA frequency findings than raw bouts, as each length-standardised bout can be specifically interpreted as lasting 1 minute.

**eText 3:** Physical activity energy expenditure, cardiorespiratory fitness, and relative physical activity intensity

In a sample of 2,043 female and 1,588 male non-exercisers with valid accelerometry data, we calculated physical activity energy expenditure during VILPA bouts using average acceleration (VO_2_ = 0.0320 x average acceleration + 7.28)^1^. Cardiorespiratory fitness was measured using a 6-minute incremental ramp cycle ergometer test with workload calculated to age, height, weight, resting heart rate, and sex for each participant. Heart rate was measured using a four-lead ECG. Following established procedures^2^ work rate at maximal heart rate was estimated by extrapolating pre-testing heart rate (work rate= 0 watts) and peak heart rate (peak work rate in watts) to age predicted maximal heart rate (max heart rate = 208 – 0.7 x age)^3^. Maximal oxygen consumption (VO_2_max; ml/kg/min) was estimated for the relationship between work rate and oxygen uptake using the equation^4^: VO_2_max = 7 + (10.8 x max work rate/ body mass (kg)).

Relative physical activity intensity (e.g., %VO2max) during VILPA bouts was calculated as the percentage of VO_2_ expended relative to each participant. %VO2max higher than 64% is equivalent to vigorous intensity^5^. The results of these analyses are summarised in **eTable 11**.

**eText 3** References

**eText 4:** Unabridged sections of summarised results in main manuscript

*Differences in the referent groups of sex-specific analyses*

Since very large differences in the characteristics of the referent group can lead to confounding by indication and spurious sex differences in dose-response, we compared health-related characteristics between females and males in the referent category (no VILPA) but we noted no systematic pattern of differences in favour of one sex group. For example, a higher proportion of males were smokers (19.4% vs. 11.6%) or were drinking above guidelines (43.4% vs. 21.6%) than females, while a lower proportion of females completed college education (34.7% vs 42.9%) or had a high fruit and vegetable consumption (26.3% vs 31.3%)

*Exercisers sample*

**eFigure 2** and **eTable 1** describe the sample derivation process and characteristics of the exercisers sample: over a mean follow-up of 7.9 (0.96) years (465,782 person-years) 58,648 exercisers were included in the all-MACE analyses (1,854 events; 749 female /1,105 male), *n*= 57,622 were included in the myocardial infarction analyses (828 events: 287 female / 541 male), *n* = 57,289 in the heart failure analyses (495 events: 210 female /285 male), and *n*=57,325 in the stroke analyses (531 events: 252 female /279 male).

*Bout length of VILPA (non-exercisers) and VPA (exercisers)*

In the core MACE analyses sample of 22,368 participants, 89.1% of VILPA bouts lasted up to 1 minute and 92.9% lasted up to 2 minutes. The median VILPA daily duration was 4.3 minutes (3.4/5.6 minutes for females/males) per day. The median length-standardised frequency was 1.7 (females: 1.4; males: 2.2) bouts per day. The median daily raw frequency was 10.1 (females: 9.3; males: 11.4 bouts per day. Among the 58,648 exercisers entered in the comparative all-MACE analyses, the large majority of (context agnostic) VPA was accrued in bouts lasting up to one (85.3% of all VPA bouts) or two (92.6%) minutes. Median VPA daily duration was 6.1 (females: 5.1; males: 8.1).

*Sensitivity analyses*

**A**ll sensitivity analyses produced results consistent with the main findings (eFigures 12-15): adjusting for clinical factors that could be considered potential mediators of the association between VILPA and MACE (**eFigure 12**), excluding participants who self-reported poor health or were underweight or were current smokers (**eFigure 13**), using the 15^th^ percentile of the VILPA distribution (0.63 minutes per day) as the referent data point (**eFigure 14**) or excluding frail participants (**eFigure 15**) produced very similar findings to the results of the main analyses presented in **Figure 3**.
